# The Transdiagnostic Connectome Project: a richly phenotyped open dataset for advancing the study of brain-behavior relationships in psychiatry

**DOI:** 10.1101/2024.06.18.24309054

**Authors:** Sidhant Chopra, Carrisa V. Cocuzza, Connor Lawhead, Jocelyn A. Ricard, Loïc Labache, Lauren M. Patrick, Poornima Kumar, Arielle Rubenstein, Julia Moses, Lia Chen, Crystal Blankenbaker, Bryce Gillis, Laura T. Germine, Ilan Harpaz-Rote, BT Thomas Yeo, Justin T. Baker, Avram J. Holmes

**Affiliations:** Department of Psychology, Yale University, New Haven, CT, USA; Department of Psychiatry, Brain Health Institute, Rutgers University, Piscataway, NJ, USA; Orygen, Center for Youth Mental Health, University of Melbourne, Melbourne, Australia; Department of Psychology, Stony Brook University, Stony Brook, NY, USA; Stanford Neurosciences Interdepartmental Program, Stanford University School of Medicine, Stanford, CA, USA; Department of Psychology, University of Pennsylvania, Philadelphia, PA, USA; Department of Psychiatry, Perelman School of Medicine, University of Pennsylvania, Philadelphia, PA, USA; Department of Psychiatry, Harvard Medical School, Boston, USA; Centre for Depression, Anxiety and Stress Research, McLean Hospital, Boston, USA; Department of Psychology, Cornell University, Ithaca, NY, USA; Institute for Technology in Psychiatry, McLean Hospital, Boston, USA; Department of Psychiatry, Yale University, New Haven, USA; Wu Tsai Institute, Yale University, New Haven, USA; Centre for Sleep and Cognition & Centre for Translational Magnetic Resonance Research, Yong Loo Lin School of Medicine, National University of Singapore, Singapore, Singapore; Department of Electrical and Computer Engineering, National University of Singapore, Singapore, Singapore; N.1 Institute for Health National University of Singapore, Singapore, Singapore; Department of Medicine, Human Potential Translational Research Programme & Institute for Digital Medicine (WisDM), Yong Loo Lin School of Medicine, National University of Singapore, Singapore; Integrative Sciences and Engineering Programme (ISEP), National University of Singapore, Singapore, Singapore; Martinos Center for Biomedical Imaging, Massachusetts General Hospital, Charlestown, USA

**Author notes:** **Correspondence:** Avram J. Holmes, Rutgers University, Department of Psychiatry, Brain Health Institute, 119 Staged Research Building, 661 Hoes Lane West, Piscataway, NJ 08854,. These authors contributed equally to this work (Sidhant Chopra, Carrisa V. Cocuzza, Connor Lawhead).

**Keywords:** Transdiagnostic Connectomes Project (TCP), Open data, Psychiatric Neuroimaging, functional MRI, brain-behavior, cognitive neuroscience

## Abstract

An important aim in psychiatry is the establishment of valid and reliable associations linking profiles of brain functioning to clinically relevant symptoms and behaviors across patient populations. To advance progress in this area, we introduce an open dataset containing behavioral and neuroimaging data from 241 individuals aged 18 to 70, comprising 148 individuals meeting diagnostic criteria for a broad range of psychiatric illnesses and a healthy comparison group of 93 individuals. These data include high-resolution anatomical scans, multiple resting-state, and task-based functional MRI runs. Additionally, participants completed over 50 psychological and cognitive assessments. Here, we detail available behavioral data as well as raw and processed MRI derivatives. Associations between data processing and quality metrics, such as head motion, are reported. Processed data exhibit classic task activation effects and canonical functional network organization. Overall, we provide a comprehensive and analysis-ready transdiagnostic dataset, which we hope will accelerate the identification of illness-relevant features of brain functioning, enabling future discoveries in basic and clinical neuroscience.

## Background & Summary

In recent years, there has been growing interest in establishing how alterations in brain anatomy and function may, at least in part, underpin the onset and maintenance of common psychiatric illnesses. However, progress in understanding these brain-behavior relationships in psychiatry has faced challenges partly due to a lack of open-access clinical cohorts and the restricted sampling of brain function and behavior across patient populations. While developments have been made in both brain-based explanatory and predictive models of clinically relevant behaviors, most of what we currently know about *in vivo* brain functioning comes from studying healthy populations. As such, the increased availability of clinically focused open-access data will facilitate the identification of network function characteristic of symptom-relevant behavioral and cognitive domains.

To date, research on the neurobiological origins of psychiatric illness has primarily focused on discrete illness categories studied in isolation. Although researchers have historically treated patient populations as discrete entities, murky boundaries often exist between nominally distinct diagnostic categories^1–3^. Transdiagnostic data collection efforts provide the unique opportunity to identify symptom and disorder general impairments that may transcend conventional diagnostic boundaries^4,5^. While existing large-scale population neuroscience datasets like the UK Biobank and Human Connectome Project^6^ have proven indispensable to foundational research questions in neuroscience, they predominantly consist of individuals without psychiatric illness. This narrow scope restricts the range of measurable behaviors, limiting our capacity to connect the full continuum of functioning to biological and environmental factors, given the incidence of psychiatric diagnoses (approximately 23% of all adults in the United States as of 2021 and a lifetime prevalence of approximately 50% starting in adolescence^7^). We present openly available data from the Transdiagnostic Connectomes Project (TCP) to work towards addressing these shortcomings. The TCP compiles richly phenotyped behavioral and MRI data from individuals with and without psychiatric diagnoses, covering a broad spectrum of human behavior. This resource provides the opportunity to uncover the neural substrates of illness-relevant behaviors across traditional diagnostic boundaries.

## Methods

In this section, we begin by describing recruitment strategies, screening procedures, and overall demographics of TCP participants. We then describe the clinician-administered measures, self-report questionnaires, and cognitive tests all participants completed. Finally, we describe the MRI data, detailing the acquisition parameters for each scan.

### Participants

Between November 2019 and March 2023, 241 participants completed the TCP study at one of two sites within the United States of America: 1) Yale University, Department of Psychology, FAS Brain Imaging Center, located in New Haven, Connecticut and 2) McLean Hospital Brain Imaging Center, located in Belmont, Massachusetts. Participants were recruited from the community via flyers, online advertisements, and patient referrals from participating clinicians. Participants provided written informed consent following guidelines established by the Yale University and McLean Hospital (Partners Healthcare) Institutional Review Boards (See Supplementary Appendix A for representative study consent forms from each site). Participants were eligible for the study if they were 1) 18-65 years old, 2) had no MRI contraindications, 3) were not colorblind, and 4) had no diagnosed neurological abnormalities. All participants underwent a Structured Clinical Interview for DSM-5 (SCID-V-RV) to assess the presence of current or past psychiatric illness. Interviews were conducted by clinical psychologists or trained research assistants who were supervised by qualified clinical psychologists. Research assistants and their clinical supervisors met weekly to discuss interview findings. The final study population included both healthy individuals without a history of illness or treatment and individuals with a diverse set of clinical presentations, including affective and psychotic psychopathology. A diagnostic and demographic breakdown of study participants is provided in Fig. 1.

**Figure 1.**
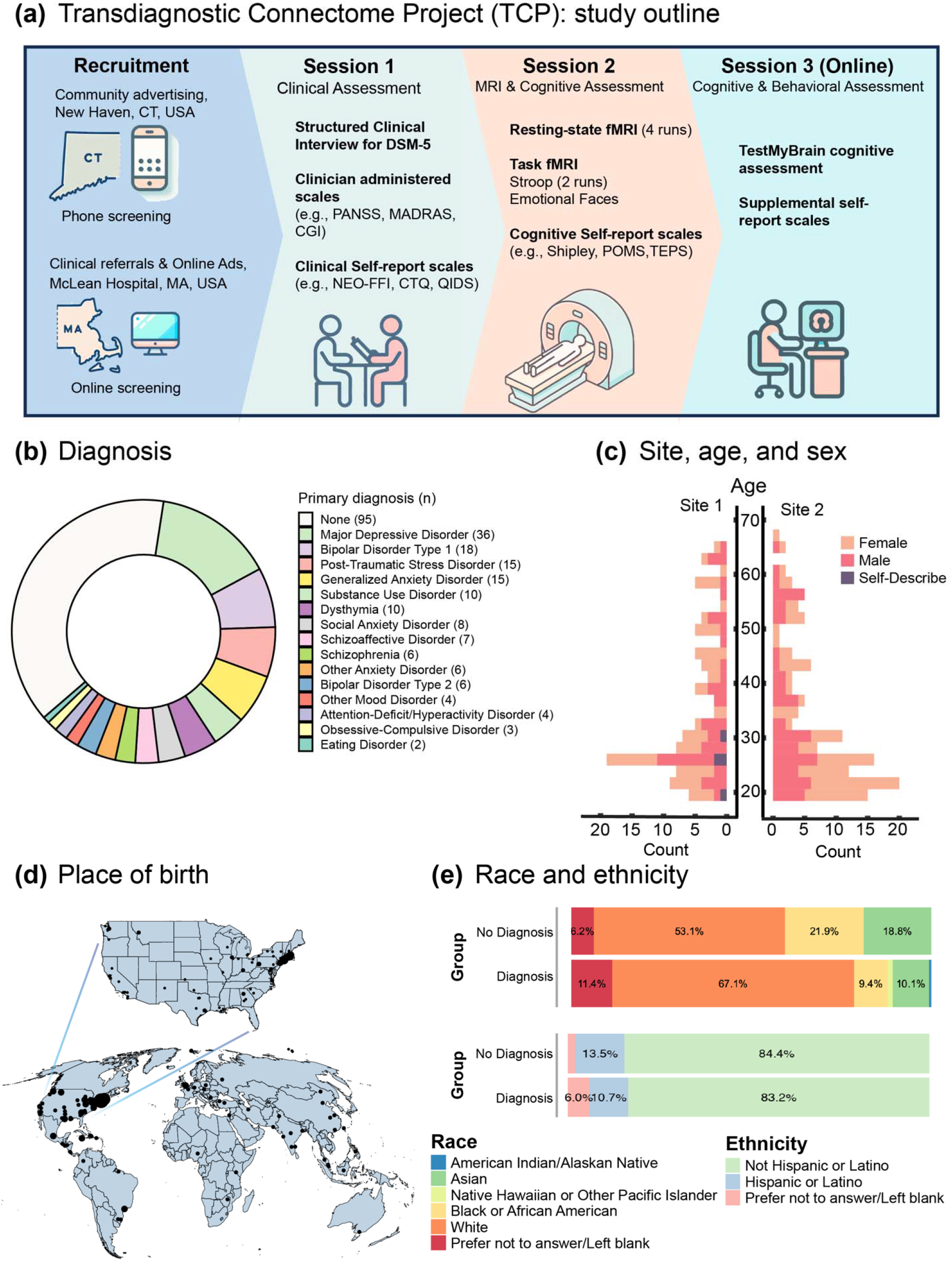
Overview of the Transdiagnostic Connectome Project (TCP). **(a)** Schematic representation of the Transdiagnostic Connectomes project, illustrating the recruitment of participants across two sites. Participants underwent three assessment sessions, including in-person clinician evaluations, self-report measures, neuroimaging, and online cognitive and self-report assessments. **(b)** Breakdown of primary mental health diagnoses for each individual based on the Structured Clinical Interview for DSM-5 (SCID-V-RV) criteria. **(c)** Bar chart displaying age distributions across the two sites, with bars colored based on the proportion of self-reported sex within each age bracket. **(d)** Geographic distribution of TCP participants’ place of birth. The dot size represents the number of participants from each location, with a world map at the bottom and an enlarged map of the United States of America on top. **(e)** Distribution of race and ethnicity according to National Institute of Health criteria, categorized for individuals with and without a mental health diagnosis.

After online or phone-based screening, participation in the TCP consisted of three study sessions (Fig. 1A). Participants completed 1) an in-person clinical, demographic, and health assessment, including a diagnostic interview, clinician-administered scales, and self-report scales (*see Behavioral measurements* for a complete list); 2) an MRI session, including anatomical, resting-state functional and task-based functional neuroimaging (see *MRI data acquisition*), as well as an additional battery of self-report cognitive and behavioral measures; 3) an online cognitive and behavioral assessment, including the TestMyBrain^8^ cognitive assessment and a supplemental set of self-report assessments (*see Behavioral measurements* for a complete list).

### MRI data acquisition

MRI data were acquired at both sites using harmonized Siemens Magnetom 3T Prisma MRI scanners and a 64-channel head coil. T1-weighted (T1-w) anatomical images were acquired using a multi-echo MPRAGE sequence following parameters: acquisition duration of 132 seconds, with a repetition time (TR) of 2.2 seconds, echo times (TE) of 1.5, 3.4, 5.2, and 7.0 milliseconds, a flip angle of 7°, an inversion time (TI) of 1.1 seconds, a sagittal orientation and anterior (A) to posterior (P) phase encoding. The slice thickness was 1.2 millimeters, and 144 slices were acquired. The image resolution was 1.2 mm^3^. A root mean square of the four images corresponding to each echo was computed to derive a single image. T2-weighted (T2-w) anatomical images with the following parameters: TR of 2800 milliseconds, TE of 326 milliseconds, a sagittal orientation, and AP phase encoding direction. The slice thickness was 1.2 millimeters, and 144 slices were acquired.

All seven functional MRI runs were acquired with the same parameters matching the HCP protocol^6,9^, varying only the conditions (rest/task) and separately acquired phase encoding directions (AP/PA). For the resting-state, Stroop task, and Emotional Faces task, a total of 488, 510, and 493 volumes were acquired, respectively, all using the following MRI sequence parameters: TR = 800 milliseconds, TE = 37 milliseconds, flip angle = 52°, and voxel size =2mm^3^. A multi-band acceleration factor of 8 was applied. An auto-align pulse sequence protocol was used to align the acquisition slices of the functional scans parallel to the anterior commissure-posterior commissure (AC-PC) plane of the MPRAGE and centered on the brain. To enable the correction of the distortions in the EPI images, B0-field maps were acquired in both AP and PA directions with a standard Spin Echo sequence. Detailed MRI acquisition protocols for both sites are available in Appendix B. In total, four resting-state (2_X_AP, 2_X_PA), 2 Stroop task acquisitions (1_X_AP, 1_X_PA), and 1 Emotional Faces task acquisition^10^ (1_X_AP) acquisitions were collected. Select participants out of the total sample did not complete each functional neuroimaging run; thus the sample sizes for each run were as follows: resting-state AP run 1, *n* = 241; resting-state PA run 1, *n* = 241; resting-state AP run 2, *n* = 237; resting-state AP run 2, *n* = 235; Stroop task AP, *n* = 226; Stroop task PA, *n* = 224; and Emotional Faces task AP, *n* = 226.

### Task fMRI paradigms

We collected functional MRI data while participants engaged in two tasks: 1) the Stroop task and 2) the Emotional Faces task. Fig. 2 shows the two task paradigms, including timing specifications for fixation periods, trial durations, inter-stimulus intervals, and response collection methods. In all task fMRI runs, stimuli were presented using the PsychoPy presentation software^11^ and projected onto a screen viewed through a mirror mounted atop the MRI scanner’s head coil.

**Figure 2.**
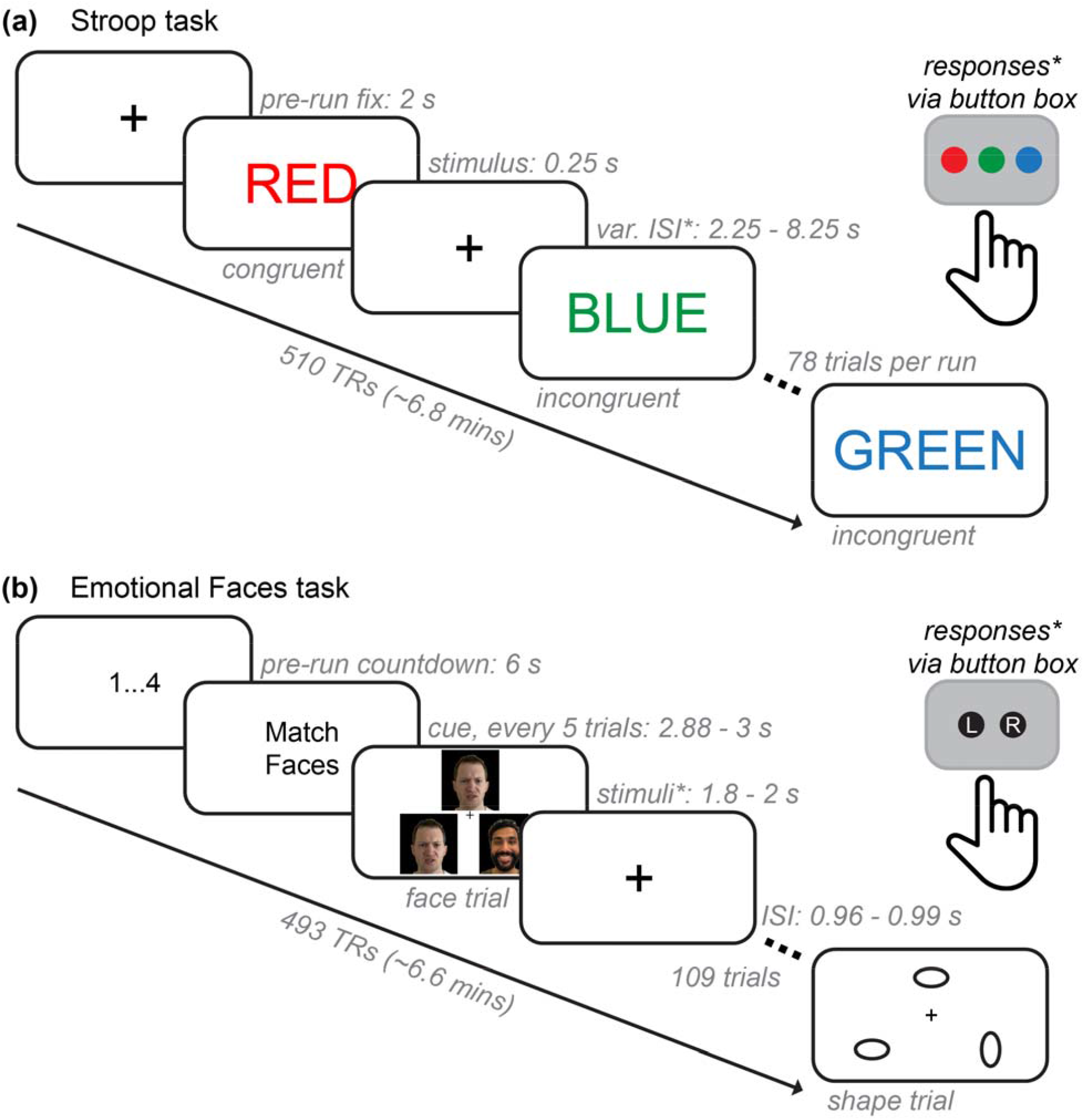
Functional neuroimaging task paradigms. **(a)** The Stroop task paradigm. For each functional run (2 total: AP and PA), an initial fixation (2 s) was followed by the presentation of color name words (red, green, or blue) in either congruent or incongruent ink color. Participants were given a button box with three buttons that were pre-mapped to record responses as either red, green, or blue and were told to identify the color of the ink on each trial. Stimuli were presented for 250 ms, and responses were allowed within the variable interstimulus interval (var. ISI; response periods denoted by asterisks), which included a range of 2.25 through 8.25 s. There were three congruent trial types, shown for about 70% of trials (18 each), and six incongruent trial types, shown for about 30% of trials (4 each). There were 78 trials for each functional run, each approximately 6.8 minutes (or 510 TRs at 0.8 s each) in total. **(b)** The Emotional Faces task paradigm. There was an initial countdown of four screens showing 1 to 4 in consecutive order (6s total). Then, every six trials showed a cue that either said “Match Faces” or “Match Shapes”. Participants were given a button box with two pre-mapped buttons that recorded responses as either “left” or “right” and were told to choose which of the two (left or right) of the bottom images matched the top image in the triangle. Stimuli were presented for 1.8 through 2 s, and there was an ISI fixation between each trial. Feedback plus ISI was approximately 1 s per trial. Each of the faces and shapes trial types were shown in equal amounts (50% of trials each). There were 109 trials altogether, which lasted a total of 6.6 minutes (493 TRs). Note that some components of these figures have been slightly modified from their original presentation form for ease of visualization and to protect the privacy of the original models; the face stimuli shown are photographs of authors of this manuscript meant to be representative, but not exact matches, of the original images used in the experiment. We confirm that all authors consented to the inclusion of their photographs in the manuscript for public dissemination.

The Stroop task is a classical experimental manipulation of cognitive control–specifically, the ability to inhibit automatic responses when presented with conflicting information to accomplish a given task goal and/or context^12–17^. During the task, participants were presented with various color name words (e.g., red, blue, and green) that were shown in various “ink” colors (i.e., font colors) and asked to identify the color of the ink (Fig. 2A). If the ink and the written word matched (e.g., “red” shown in red font), this was a congruent trial and is relatively easy and fast to identify. However, if the ink and the written word did not match (e.g., “red” shown in blue font), this was an incongruent trial and is more difficult and slower to identify. This is known as the “Stroop effect” (or “Stroop interference”) and is quantified via reduced accuracy and slower reaction time on incongruent versus congruent trials. The Stroop effect is generally more pronounced (i.e., larger accuracy and reaction time differences between conditions) when a participant has lower inhibitory cognitive control or difficulty recruiting the neurocognitive resources needed to process conflicting information accurately^18^. Resolving interference in an experimentally manipulated context is thought to capture the extent to which one can deploy cognitive flexibility and/or selective attention in everyday life^19–21^. The Stroop effect has been investigated using various task adaptions in a variety of clinical research programs, including studies of psychotic^22–25^, attentional^26–29^, mood^30–32^, and substance use disorders^33–35^. Additionally, cognitive control is negatively impacted across a large number of psychiatric disorders^36–39^. Therefore, behavioral performance and neurocognitive processes exhibited while performing the Stroop task are well-suited to the transdiagnostic research questions addressable with the TCP dataset.

The Emotional Faces task^10^ is also a widely implemented task paradigm in neuroimaging. Participants were presented with images of human faces or geometric shapes and asked to categorize stimuli as either faces or shapes (Fig. 2B). Trials that showed pictures of human faces included people with neutral, positive (e.g., happy), and negative emotional expressions (e.g., anger) taken from the NimStim database of face stimuli^40^; trials showing shapes included ovals with different orientations. Three images were arranged in a triangle, and participants were instructed to indicate which of the two bottom shapes or faces matched the shape or face at the top of the screen. This task is relatively easy; therefore, behavioral performance is not typically of central interest. Instead, this task paradigm reliably elicits a response within regions of the amygdala during emotional face relative to shape trials and can provide neuroimaging studies with 1) essential benchmarking using a well-established task activation and 2) an entry point for neuroscience research questions on affective processing. The latter is an important consideration for transdiagnostic research, given that a variety of psychiatric conditions are known to involve dysregulated processing of emotions and deficits in social cognition. These topics have been examined using the Emotional Faces task in studies of autism spectrum disorder^41,42^, anxiety disorders^43,44^, post-traumatic stress disorder^45^, psychotic disorders^46^, psychopathy^47^, and those exhibiting social phobia^48^.

### Behavioral measurements

Table 1 lists the entire battery of assessments across the three testing sessions. The selected measures index a broad range of functional, lifestyle, emotional, mental health, cognitive, environmental, personality, and social factors. These assessments include multiple commonly used clinical tools such as the Montgomery-Åsberg Depression Rating Scale (MADRS^49^), Depression Anxiety Stress Scale (DASS^50^), Positive and Negative Syndrome Scale (PANSS^51^), and Young Mania Rating Scale (YMRS^52^), as well as scales that capture distinctive aspects of experiences such as Temperament and Character Inventory^53^, Temporal Experience of Pleasure Scale^54^, Experience in Close Relationship Scale^55^, and the Positive Urgency Measure^56^. The dataset also includes common measures of cognition, including the Shipley Vocabulary Test^57^ and the TestMyBrain suite^8^, including matrix reasoning, as well as self-report measures such as Cognitive Emotion Regulation Questionnaire^58^, Cognitive Failures Questionnaire^59^ and Cognitive Reflection Test^60^. Item-level participant responses are available for download (see *Data Records*). The associated distributions, missingness, means, medians, and standard deviation for the total sample for all scales and subscales are provided in Supplement Table 1.

**Table 1.**
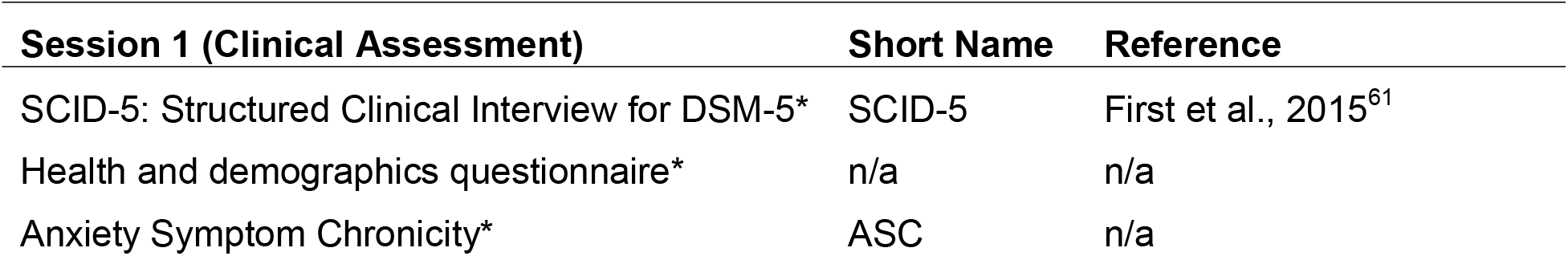
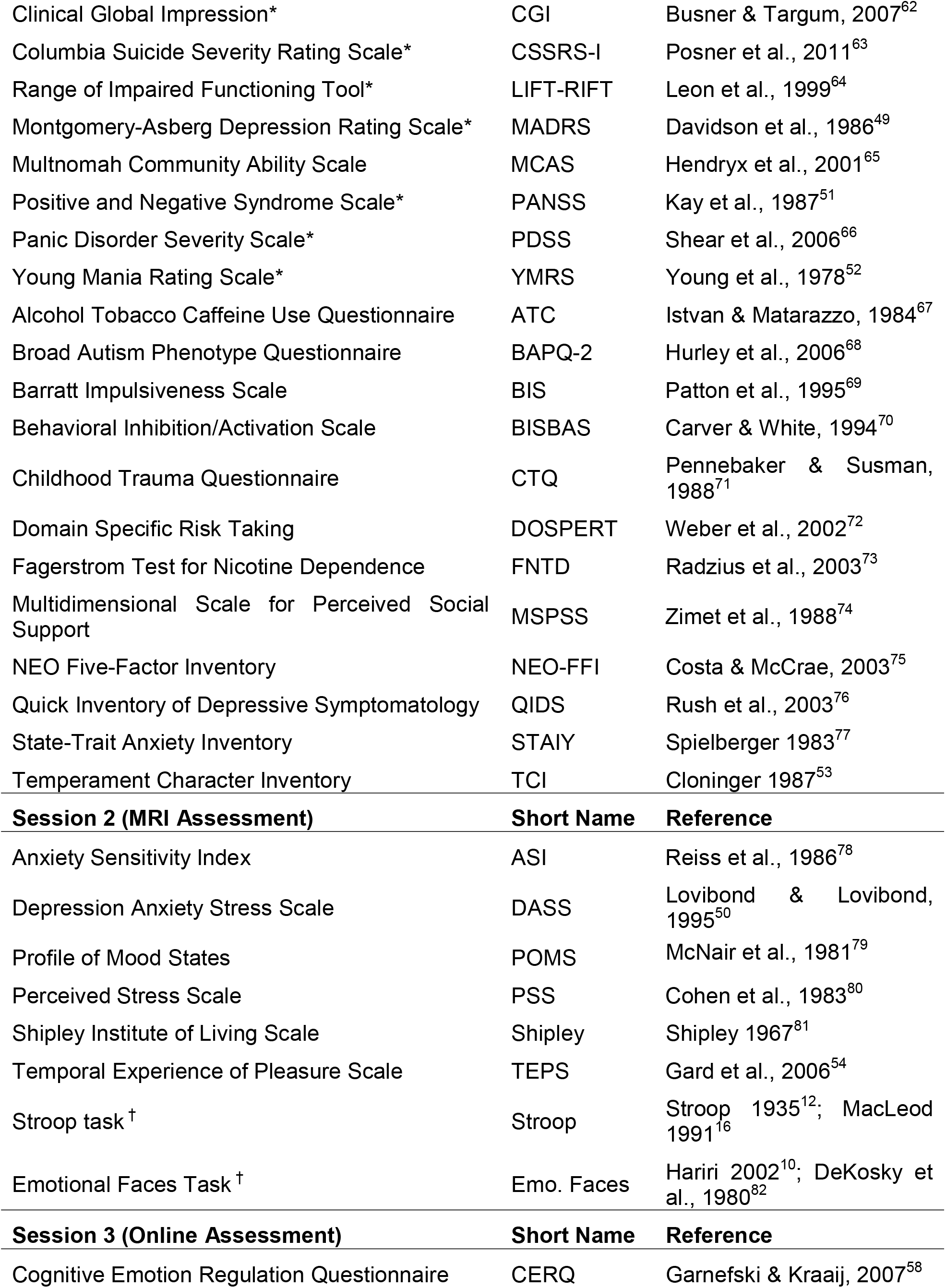
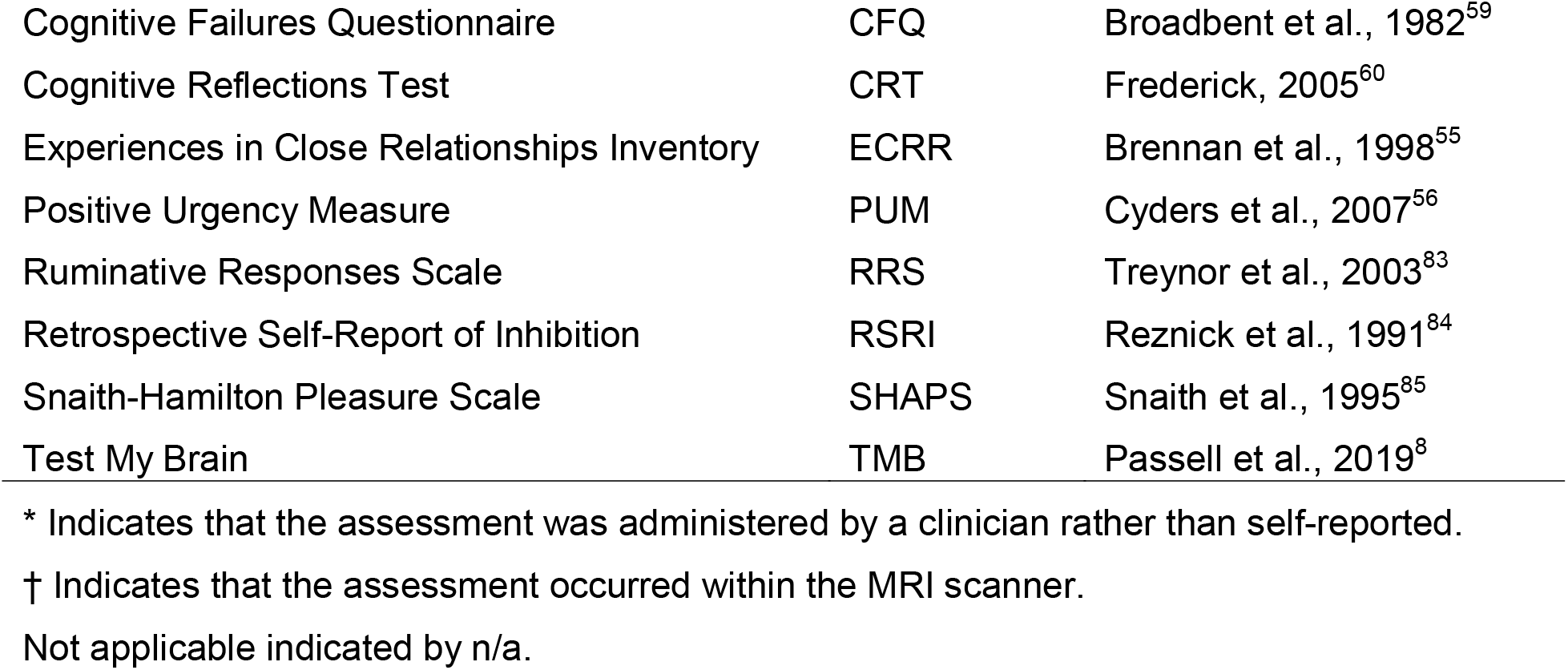
List of all behavioral assessments.

## Data Records

Raw neuroimaging and behavioral data can be accessed via OpenNeuro: https://openneuro.org/datasets/ds005237. Additionally, raw and processed data can be accessed via the National Institute of Mental Health Data Archive (NDA) data repository (https://nda.nih.gov/edit_collection.html?id=3552) upon publication. The dataset is released as part of *Open Access Permission*, has been consented for broad research use, and can be accessed by users who are not affiliated with an NIH-recognized research institution. Note that 5 participants did not grant permission to share their data via openly online-hosted repositories and are excluded from this release.

Raw MRI data have undergone DICOM to NifTI conversion using *dcm2niix*^86^ and are provided in BIDS-compliant^87^ format. BIDS organizes data into a standardized file, folder and naming structure. Imaging data for each participant is stored in folders named according to the assigned Global Unique Identifier (GUID), an alphanumeric code created by the NDA GUID Tool. For BIDS data uploaded to OpenNeuro, a ‘sub-’ prefix was added to subject folders and filenames. Each participant folder contains an ‘*anat*’ subfolder containing T1-w and T2-w images, a ‘*func*’ subfolder containing all runs of functional data as well as event files for task runs, and a ‘*fmap*’ subfolder containing the field maps. Each image and event file has a corresponding *.json* file that contains relevant meta-data. All anatomical images were defaced prior to uploading on the NDA using the *face_removal_mask* function from the R library *fslr*^88^ (R package version: 2.52.2, R version: 4.3.0), which is a wrapper library of FSL^89^ (version 6.0.5.1). The *face_removal_mask* function is an R implementation of the Python package *pydeface*^90^. The demographic and behavioral data are available as *.csv* files, with a separate file for each scale and a corresponding NDA-conformant dictionary delineating available data fields.

To facilitate the use of the TCP data, in addition to raw BIDS formatted data, we provide both raw data and processed derivatives according to the HCP processing pipelines (version 4.7.0, see *Human Connectome Project minimal processing* below for details) on the NDA, including anatomical surfaces, as well as minimally processed and denoised functional timeseries in both densely sampled surface space and standard volumetric space. A full list of HCP-related derivatives can be found at: https://www.humanconnectome.org/study/hcp-young-adult/document/1200-subjects-data-release. We additionally provide analysis-ready functional connectivity matrices using cortical, subcortical, and cerebellar regions. The processing and denoising pipeline, and quality control procedures are described below in *Technical Validation*.

## Technical Validation

In this section, we first describe the processing and denoising pipeline used for the MRI data. Next, we benchmark this process by examining 1) the residual relationship between in-scanner movement and functional connectivity before and after denoising, 2) the effect of region-to-region distance on this residual relationship, and 3) the number of statistically significant associations between head motion and functional connectivity. Subsequently, we explore the presence of group-level canonical functional network structures and assess the similarity of these structures across different fMRI runs. Additionally, we report commonly computed network diagnostics^91^ given by graph theory, canonical task-based fMRI activation contrasts, and behavioral outcomes from in-scanner tasks. Finally, we examine correlation patterns across behavioral scales and subscales for diagnosed and non-diagnosed individuals and investigate latent behavioral structures using dimensionality reduction. All processing and analyses reported below, except differences in behavioral measures reported in Fig. 7 and Fig. 8, are conducted on the entire sample and do not differentiate between those with and without a diagnosis.

### Human Connectome Project minimal processing

MRI data were minimally processed and denoised via the Human Connectome Project (HCP) pipelines^92^, version 4.7.0 (https://github.com/Washington-University/HCPpipelines). Broadly, this includes: 1) FreeSurfer structural MRI processing, 2) functional MRI volume processing, 3) functional MRI surface processing, 4) denoising via ICA-FIX, 5) “multimodal surface matching” registration (MSMAll^93^), and 6) de-drifting, resampling, and applying the MSMAll registration. Tools implemented by HCP pipelines are mainly adapted from the FMRIB Software Library (FSL) and FreeSurfer^94^, to improve the spatiotemporal accuracy of MRI data, particularly with acquisition advancements such as multiband acceleration^92,95,96^.

In brief, anatomical T1w/T2w images were used to create MNI-aligned structural volumes in each participant’s native space. These images were corrected for gradient nonlinearities and magnetic field inhomogeneities and reconstructed into segmented brain structures. Folding-based surface registration to the standard FreeSurfer atlas (i.e., *fsaverage*) and to the high-resolution Conte69 atlas^97^ was followed by downsampling to various spatial resolutions (2 mm used herein). fMRI volume processing steps further corrected for spatial distortions and implemented motion correction, bias field correction, and normalization. Motion correction was implemented via FLIRT registration of individual frames to a single-band reference image. In HCP-style, we provide motion parameters in separate files to characterize x/y/z translation, rotation, and their derivatives, as well as demeaned and detrended versions of each (24 parameters total), which may be used for nuisance regression (see *fMRI denoising*). Echo planar distortions were corrected by FSL’s “topup”^98,99^ using spin echo field maps that were acquired in the opposite phase encoding directions of each scan.

fMRI surface processing steps transformed 4D volumetric timeseries into 2D surface-based timeseries that were registered to a standard set of grayordinates across all participants. This involved HCP’s “partial volume weighted ribbon-constrained volume-to-surface mapping algorithm”^92^, which uses non-resampled images in each participant’s native space to align surfaces along tissue contours. Surfaces were smoothed using the “geodesic Gaussian surface smoothing algorithm”^92^ and additional 2 mm FWHM smoothing that enhances subcortical regularization. Following denoising (see *fMRI denoising* for full details), surface-based functional timeseries were aligned with MSMAll^93^. MSMAll improves inter-participant alignment by using areal features from multiple sources in the reference pipeline developed by Glasser et al.^93^, including cortical folding, sulcal depth, T1w/T2w myelin maps, resting-state network maps, and visuotopic maps. The HCP minimal processing pipelines are optimized for surface-based processing of high-resolution anatomical and functional MRI images. Thus, we used the resulting grayordinate timeseries from each of the seven functional runs to perform the quality assurance and preliminary analyses reported herein. This included cortical, subcortical, and cerebellar vertices (i.e., 91,282 “whole-brain” gray ordinates) that were regionally mapped according to their corresponding atlas schemes (see *Parcellating timeseries into brain regions and functional connectivity*).

### fMRI denoising

To remove sources of noise such as movement, scanner drift, and physiological artifacts, we implemented the data-driven MELODIC independent component analysis-FMRIB ICA-based Xnoiseifier (ICA-FIX; ^100,101^). ICA-FIX is a denoising approach performed on each functional timeseries for each participant. Given that we closely followed the HCP’s acquisition protocols and minimal processing pipelines, we applied FIX classifiers pre-trained on HCP data (using the HCP pipeline default, “HCP_hp2000”). To provide flexibility for the varied research questions addressable with the TCP dataset, we separately performed both “single-run” and “multi-run” ICA-FIX. Single-run ICA-FIX, which was used for benchmarking and quality control herein (see Figure S1 for example quality control scenes provided by the HCP pipelines), was performed on each functional run independently, and multi-run ICA-FIX was performed on: 1) concatenated the four resting-state runs, and 2) three task runs functional timeseries. Consistent with the HCP processing pipeline, we used a high-pass temporal filter of 2000s FWHM during single-run ICA-FIX. While spatial ICA likely provides components with better signal-to-noise separation via multi-run FIX (given longer timeseries data), single-run FIX is likely optimal for research questions (or benchmarking) requiring statistical independence across functional runs.

We performed global signal regression (GSR) to further control for noise sources in fMRI timeseries data^102–105^. GSR has been shown to remove global sources of noise^106–108^ and improve behavioral prediction models^109^. However, it has also been shown that the global signal can carry behaviorally-relevant information^109–111^. Given this ongoing debate surrounding the use of GSR^104,105^, we provide denoised derivative timeseries with and without GSR and evaluate the impact of GSR on fMRI data in the *Technical Validation* sections below. We performed GSR for each participant and each functional run by regressing the mean timeseries across all vertices from each vertex^112^.

### Parcellating timeseries into brain regions and functional connectivity

We parcellated the dense (i.e., 91,282 vertices) CIFTI timeseries into 434 brain regions that covered the cortex, subcortex, and cerebellum, by averaging the functional timeseries of the vertices belonging to a given region together. We used a previously validated surface-based functional atlas to parcellate the cortex into 400 regions^113^. This “homotopic” cortical atlas is a recent update to the widely-used Schaefer atlas^114^ that improves upon hemispheric lateralization in brain systems known to be asymmetric, such as language processing regions. This atlas is openly available via website links in Schaefer et al.^114^. Subcortical vertices were parcellated into 16 bilateral brain regions (32 total) that are part of the medial temporal lobe, the thalamus, and the striatum (including the pallidum)^115^. These 32 regions were yielded by the “scale II” resolution provided by Tian and colleagues, which we implemented based on the finding that anatomical boundaries are well-captured at this resolution while also providing functional subdivisions. This atlas is openly available via website links in Tian et al. ^115^. Lastly, we parcellated the cerebellum into one region per hemisphere (2 total) using the atlas provided by Buckner et al.^116^. Following the best practices provided by Buckner and colleagues, we regressed neighboring (6 mm) cortical signals from cerebellar vertices before parcellation to account for potential spatial autocorrelation between these brain segments.

Functional connectivity matrices were derived for each participant and each functional scan at three different stages of the pipeline: after minimal processing (see *Human Connectome Project minimal processing pipeline*), after ICA-FIX denoising, and after GSR was applied to denoised data (see *Functional MRI denoising*). The pairwise product-moment correlation between regional timeseries was computed, resulting in a 434 by 434 functional connectivity matrix for each participant and fMRI run.

### Functional connectivity quality control and benchmarking

Estimates of brain function derived from fMRI, such as functional connectivity, are sensitive to artifacts from multiple sources, including in-scanner head movement, respiratory motion, and scanner effects^117^. To assess the success of denoising procedures, residual relationships between functional connectivity and in-scanner head motion can be examined and compared at different stages of the processing pipeline^107,118,119^. Head motion during each fMRI scan was estimated using framewise displacement (FD), a summary measure of head movement from one volume to the next^107^. For each scan, FD was calculated according to the method described by Jenkinson et al.^120^ and the resulting FD trace was band-stop filtered and down-sampled to account for the high sampling rate of the multiband fMRI acquisition^121^. Distributions of mean FD for each participant and each fMRI run are provided in Figure S2.

### FD-FC correlations

For each rs-fMRI run, we computed the cross-participant Spearman correlation between FD and functional connectivity at each pair of regions after denoising (Fig. 3A-B). Similar to previous work^107,119^, before denoising (i.e., after minimal processing), we find widespread positive associations between functional connectivity and FD with a moderate effect size across most connections in all four runs (Fig. 3A, upper triangles), indicating a strong and wide-spread effect of in-scanner head motion on functional connectivity estimates. The mean correlation over the brain ranges across each run from ρ = .12 - .16 (Fig. 3B). ICA-based denoising consistently reduced these positive associations (ρ = .04 - .08; Fig. 3B) and adding GSR brings the mean correlation to ρ = .00 - .02 (Fig. 3A, lower triangles; Fig. 3B). However, in some cases (Rest 1 AP and Rest 1 PA), both ICA-based denoising and GSR induced negative associations between FD and functional connectivity. Overall, denoising procedures substantially reduced FD-FC associations across the brain. A broadly similar pattern of results was evident across the three task-based fMRI runs (Fig. 4A-B), with mean correlations across each run ranging from ρ = .07 - .14 before denoising, from ρ = .00 - .04 after denoising and ρ = .00 - .02 after GSR, albeit with GSR having a less pronounced effect on reducing FD-FC correlations.

**Figure 3.**
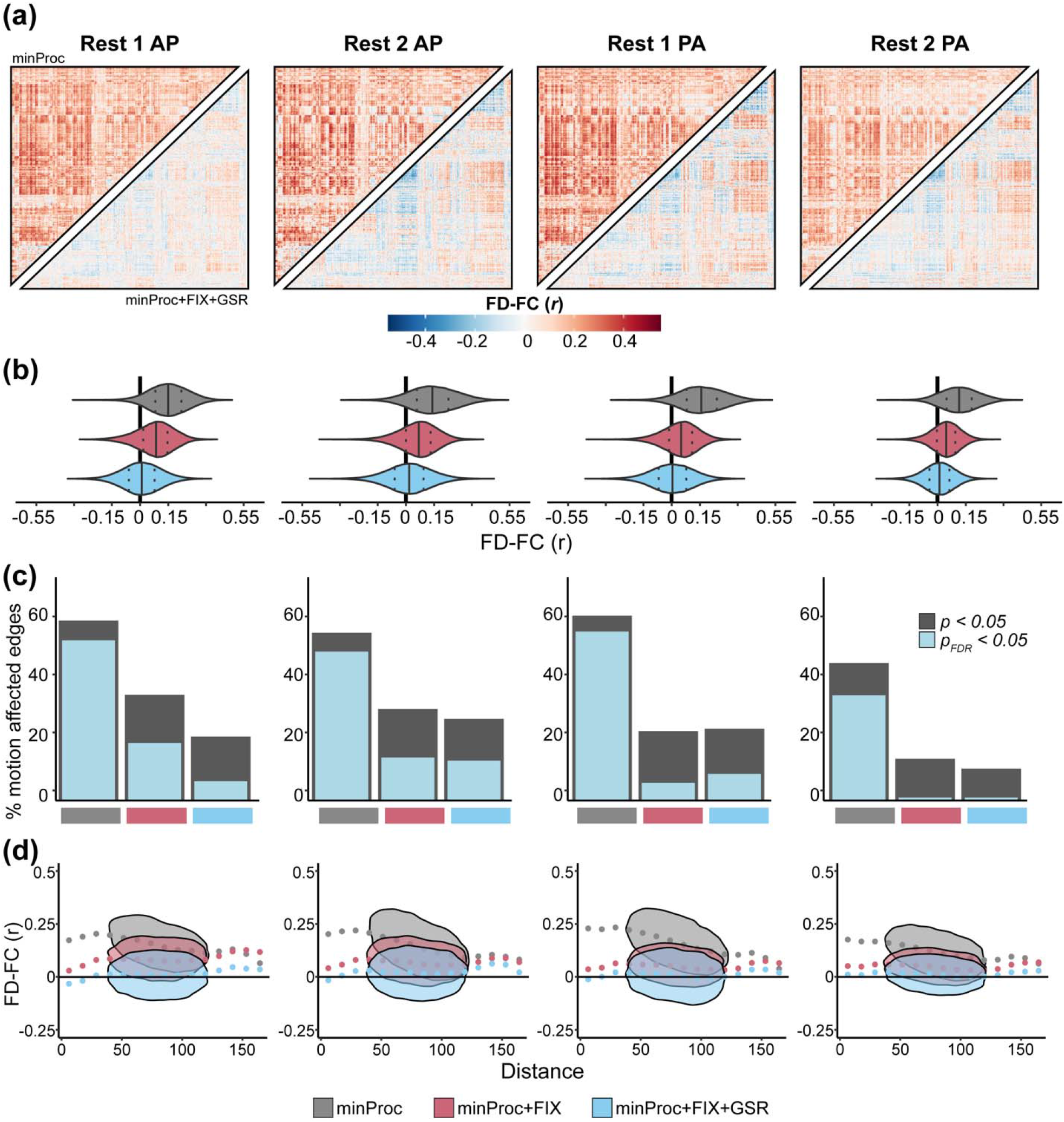
Functional connectivity and head motion association across processing stages for resting-state fMRI runs. **(a)** Inter-participant correlation between functional connectivity (FC) and mean framewise displacement (FD) at each of 93,961 connections for each resting-state fMRI run before denoising, i.e., after minimal processing (minProc; upper triangles) and after ICA-based denoising (FIX) and Global Signal Regression (GSR; lower triangles). **(b)** Distributions of FD-FC correlations for each run resting-state fMRI at three different processing stages: minProc, FIX, and GSR. **(c)** Percentage of connections significantly (*p*<0.05, gray; *p*<0.05_FDR_, light blue) correlated with FD for each resting-state fMRI at each of the three processing stages. **(d)** Associations and density between FD-FC values and pairwise Euclidean distance between 432 regions for each resting-state fMRI run and processing stage.

**Figure 4.**
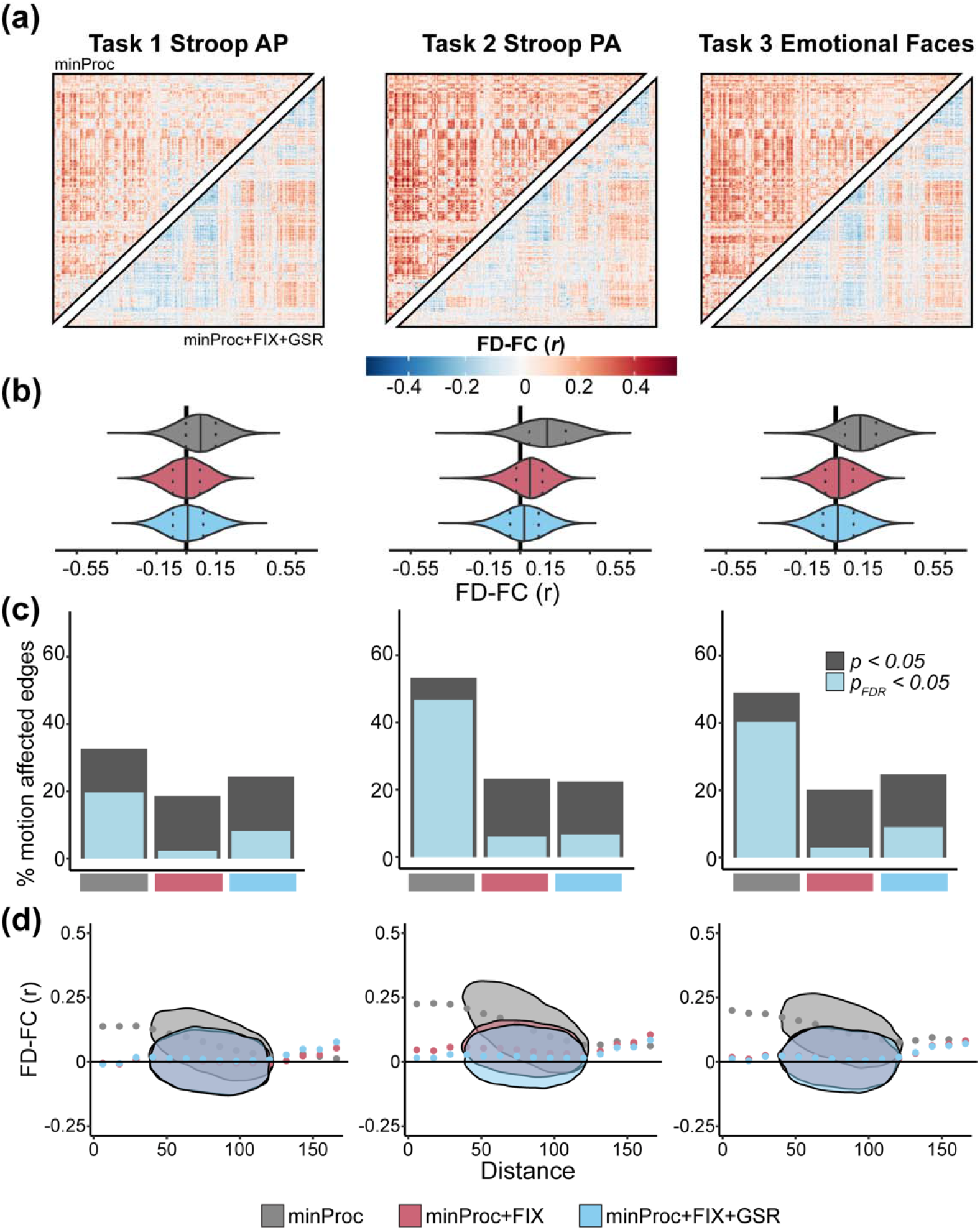
Functional connectivity and head motion association across processing stages for task-based fMRI runs. **(a)** Inter-participant correlation between functional connectivity (FC) and mean framewise displacement (FD) at each of 93,961 connections for each task-based fMRI run before denoising, i.e., after minimal processing (minProc; upper triangles) and after ICA-based denoising (FIX) and Global Signal Regression (GSR; lower triangles). **(b)** Distributions of FD-FC correlations for each run task-based fMRI at three different processing stages: minProc, FIX, and GSR. **(c)** The percentage of connections significantly (*p*<0.05, gray; *p*<0.05_FDR_, light blue) correlated with FD for each task-based fMRI at the three processing stages. **(d)** Associations and density between FD-FC values and pairwise Euclidean distance between 432 regions for each task-based fMRI run and processing stage.

When examining the proportion of connections significantly correlated with FD, we find that before denoising, across rs-fMRI runs, 35% to 56% of connections met the threshold for significance (*p*_FDR_<.05; Fig. 3D). After ICA-based denoising, there was a substantial drop in motion-affected connections, reflecting 1% to 19% of connections, depending on the fMRI run (Fig. 3D). In all runs except Rest 1 PA, GSR further reduces the number of motion-affected connections. A similar pattern of results was seen for the task-fMRI run, except that GSR consistently increased the number of motion-affected connections across all three runs compared to ICA-based denoising alone (Fig. 4D).

### FD-FC distance dependance

For each rs-fMRI run, we examined how the FD-FC relationship varies as a function of the pairwise Euclidean distance between the centroid of regions, as head motion generally has a more pronounced effect on the FD-FC correlation for short-range connectivity^117,118,122^. Specifically, the presence of head motion can artifactually increase correlations between regions that are closer and decrease correlations between areas that are further apart. This pattern of distance-dependence motion contamination was present across all rs-fMRI runs before denoising and was substantially reduced after ICA-based denoising (Fig. 3C). The addition of GSR did not have a notable impact. Similar results were found for task-fMRI runs (Fig. 4C).

### Functional network structure

The regions in the cortical and subcortical atlases used are each provided with corresponding network assignments. We used the 17-network solution for cortical regions^123^ and a 3-network anatomical solution for subcortical regions comprising the medial temporal lobe, striatum, and thalamus^115^. Lastly, we considered the two cerebellar regions as part of their own network, altogether resulting in 21 functional networks (Fig. 5A-B).

**Figure 5.**
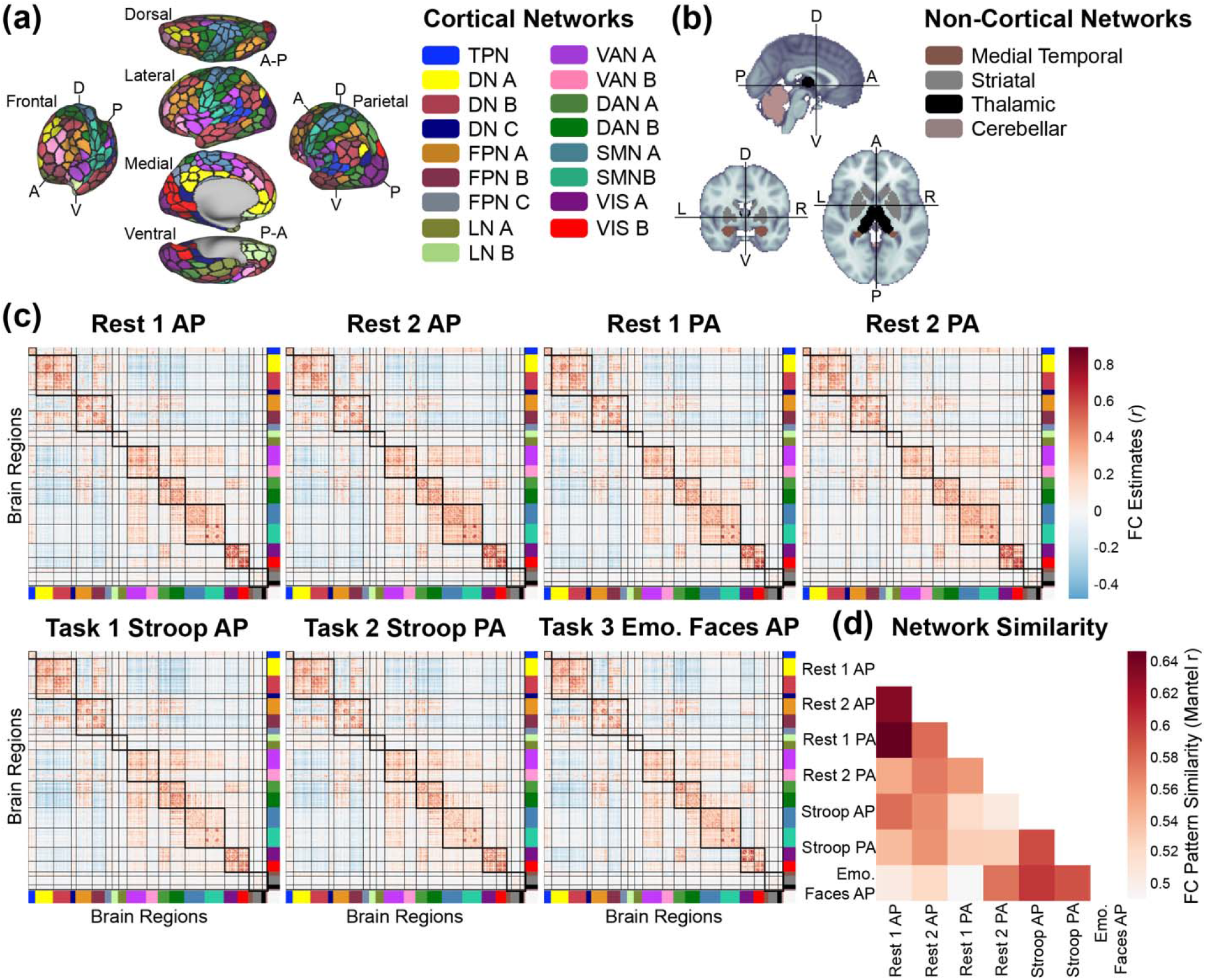
Average functional network structure and similarity across each fMRI run. **(a)** Whole-brain regional parcellation (black borders, 432 regions^113,115,116^) and network partition (filled in colors, 21 networks) across the cortex (a) and non-cortex **(b)** (see *Parcellating timeseries into* brain *regions and functional* connectivity *for full details*). TPN: temporal parietal network, DN: default network, FPN: frontoparietal network (sometimes referred to as cognitive control network), LN: limbic network, VAN: salience/ventral attention network, DAN: dorsal attention network, SMN: somatomotor network, VIS: visual network. The medial temporal network contains hippocampal and amygdalar regions; the striatal network contains caudate, nucleus accumbens, putamen, and globus pallidus regions. The thalamic and cerebellar networks contain only thalamus and cerebellum, respectively. **(c)** Functional connectivity (FC) estimates for 6 neurocognitive states (across-participant averages). Thick and thin black borders delineate within- and between-network borders, respectively. **(d)** Similarity of functional connectivity patterns for each pair of neurocognitive states using the nonparametric Mantel test.

Functional connectivity estimated from rest and task fMRI typically exhibits a reliable and robust set of brain networks that encompass spatially discontinuous regions with temporally correlated activity. The topology and connectivity strengths of these networks are heritable^124^ and linked with behavioral outcomes in both health^125^ and illness^126^. To examine whether the expected functional network structure was present across each of the fMRI runs, functional connectivity matrices were Fisher z-transformed, averaged across participants, and then the group-average matrix was transformed back into product-moment correlations (Fig. 5C, visualizing FC estimated from data with denoising and GSR). The columns and rows of each group average connectivity matrix were reordered according to the previously established 21-network scheme (Fig. 5A-B), revealing the expected pattern of pronounced within-network connectivity (Fig. 5C, diagonal blocks), compared to reduced between-network connectivity (Fig. 5C, off-diagonal blocks). We report group average functional connectivity matrices with and without ICA-based denoising and with and without GSR in Figure S3.

Across the network neuroscience literature, there is strong evidence that functional network organization is highly similar across neurocognitive states, particularly across rest- and task-state connectivity estimates^127–130^. Rest-to-task changes in network connectivity patterns are typically limited in magnitude, small in extent (i.e., only a subset of regional pairs change), and tend to be decreases in connectivity^131^. It is worth noting, however, that prior work suggests these relatively subtle task-evoked changes likely carry important information for behavioral and/or cognitive functioning^130,132^. In the present work, we report the similarity of connectivity patterns for each pair of functional runs (Fig. 5D), estimated with the nonparametric Mantel test^133^. As expected, stronger similarity patterns were generally observed across similar states, i.e., resting states (runs 1 AP/PA and 2 AP/PA), Stroop (AP/PA), and emotional faces task states.

### Benchmarking functional network properties with graph-theoretic metrics

A common approach in network neuroscience is to leverage graph theoretic tools to characterize the properties underlying brain connectivity patterns via neurobiologically relevant and computationally tractable metrics^91,134–140^, sometimes referred to as network diagnostics^141^. Given that the neuroimaging acquisition and processing protocols of the TCP dataset were optimized for functional network analysis, we aimed to demonstrate that well-established network properties are discoverable in TCP connectomes. To this end, we implemented the following network analyses: 1) clustering coefficient, 2) degree strength, and 3) betweenness centrality (Fig. 6). We used the Brain Connectivity Toolbox^91^ (http://www.brain-connectivity-toolbox.net) adapted to Python (i.e., bctpy), which is openly available here: https://pypi.org/project/bctpy/. Each metric was applied to both pairwise regional functional connectivity estimates (i.e., “region level”) and subsequently averaged based on network assignment (i.e., “network level”). Given that we used product-moment correlation to estimate functional connectivity (see *Parcellating timeseries into brain regions and functional connectivit*y and *Functional connectivity quality control and benchmarking*), we used the “weighted and undirected” variant^91,142^ of these metrics. We used min-max normalization to average network metric scores across participants and scaled all results between 0 and 1. Min-max normalization was used to maintain the relative distribution of scores across participants while scaling possible values to a fixed, comparable range. However, we encourage future investigations to consider normalization techniques that account for potential extremes if appropriate for the research question and network metric.

**Figure 6.**
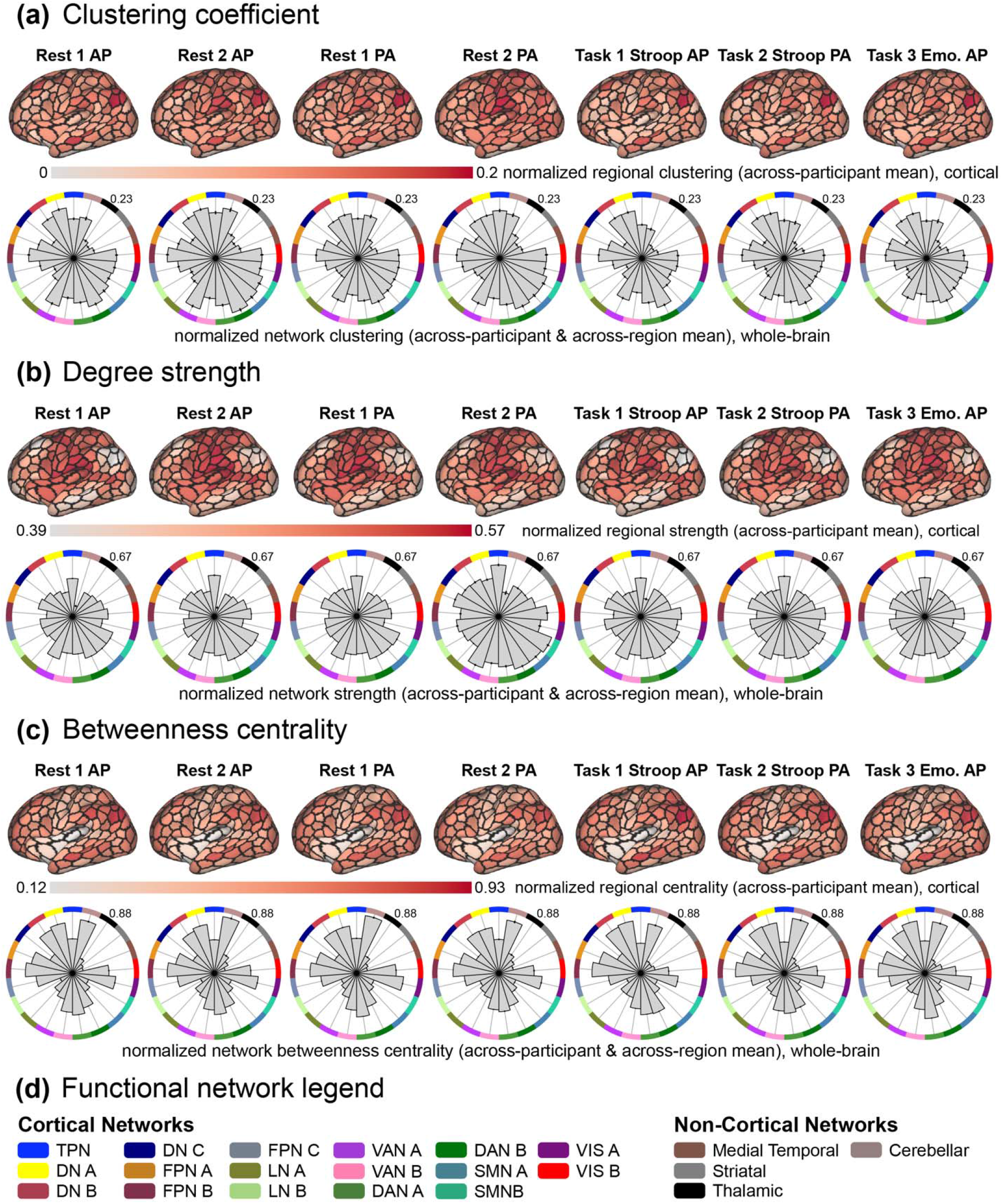
Network properties for each fMRI run. **(a)** The across-participant average (i.e., group-level) clustering coefficient scores at the region level (*top*: projected onto surface-based cortical brain schematics with black outlines delineating cortical regions given by Yan et al.^113^; showing the lateral view of the right hemisphere only for ease of visualization) and at the network-level (*bottom*: polar plots organized by network assignment, which follows label colors in panel d, and the maximum value is listed on the top right of each plot). Before averaging, scores were normalized across the entire dataset using min-max feature scaling (between the values of 0 and 1). **(b)** The same as panel a, but for the metric of degree strength. **(c)** The same as panels a and b, but for the betweenness centrality metric. **(d)** Legend of colors used to indicate functional network assignment in polar plots, corresponding to Fig. 5.

The clustering coefficient is a metric for quantifying the extent that local connectivity patterns are segregated and is based on the average “intensity” or “abundance” of triangles that are present around a given region^142–144^. Triangles refer to connectivity estimates adjacent to the given region, and their intensity is given by the relative extent (i.e., fraction) that those regions are also connected. Clustering can be thought of as the relative magnitude that a neighborhood of connections is “established” or “complete”. Therefore, a relatively large region-level score indicates clustered connectivity surrounding that region, and a large network-level score indicates that regions in that functional system tend to cluster together. A common inference for such clustering is that it supports local efficiency of information processing and community structure^145^. Across participants, we observed non-random clustering patterns across brain regions and functional networks (Fig. 6A), which is broadly consistent with prior work demonstrating that functional brain networks exhibit clustering and expected properties such as small worldness^145^. In resting-state connectivity matrices, high clustering was observed in all dorsal attention, somatomotor, temporoparietal, cerebellar, and visual network regions, as well as ventral attention A, frontoparietal B, and default A and B network regions. This resting-state pattern was slightly modified during Stroop and Emotional Faces task states. Stroop connectivity patterns exhibited reduced clustering within somatomotor, visual, cerebellar, and temporoparietal network regions. Emotional Faces connectivity patterns exhibited an overall similar pattern to resting-state, just reduced in magnitude.

The degree strength of a given brain region is a straightforward and commonly applied network metric that quantifies the relative magnitude of connectivity estimates for a given region^91^. This is similar to the degree – the number of connections for a given region – but more appropriate for fully connected networks. Regions with relatively large degree strength are thought to be more important in a given network, and the network-level degree strength is often interpreted as the wiring cost or density of that brain system^91,146^. This is an important consideration for research questions that are sensitive to heterogeneous metabolic demands across different brain systems, different participant groups, or both, given the association between the efficiency of energy consumption and degree of connectivity patterns^147^. Here (Fig. 6B), resting-state functional connectivity patterns exhibited relatively higher degree strength in temporoparietal and somatomotor network regions, as well as ventral attention A, dorsal attention B, and default C network regions. This pattern was consistent but with reduced overall magnitude in all task states, suggesting that the network-level degree strength is a relatively stable metric across neurocognitive states. An interesting exception was the relative reduction of degree strength exhibited by default A and B network regions in Stroop task states, which is broadly consistent with the traditional view of the default network being less prominent during task engagement^148^ (although see Spreng^149^).

Betweenness centrality is a metric that estimates the extent to which a given region interacts with other regions^150,151^. Such central areas are sometimes called “hubs” and are integral for integrated information flow across brain systems^91,152,153^. Multiple network metrics quantify different hub properties; however, betweenness centrality quantifies the fraction of shortest connectivity paths that contain the given region^91^. Therefore, a high betweenness centrality score indicates that the region participates in a relatively large number of short paths in the brain network and can be conceptualized as bridges in the system. Across all resting-state connectomes, regions in the frontoparietal, dorsal attention, visual, and cerebellar networks exhibited high betweenness centrality, as well as regions in the default A/B, and ventral attention B networks (Fig. 6C). This pattern was similar across Stroop task-state connectivity patterns, but with a relatively more pronounced centrality in frontoparietal B network regions, and relatively reduced centrality in visual and cerebellar network regions, which is broadly consistent with the theory that frontoparietal network interactions support cognitive control processes^154,155^. During the Emotional Faces task, connectivity patterns exhibited relatively more centrality in visual A and dorsal attention A network regions, which is expected given that this task requires participants to make judgements about the context of visual stimuli.

### Inter-scale correlations and latent structure across measures of behavior

To assess associations between 110 scales and subscale scores across participants (see Table S1 for distributions and descriptive metrics), after z-scoring each scale, we computed product-moment correlations between each pair of measures using pairwise complete observations. This was done separately for individuals with (Fig. 7A, lower triangle) and without a diagnosis (Fig. 7A, upper triangle), with both groups showing a highly similar inter-scale correlation structure (*r* = .74).

**Figure 7.**
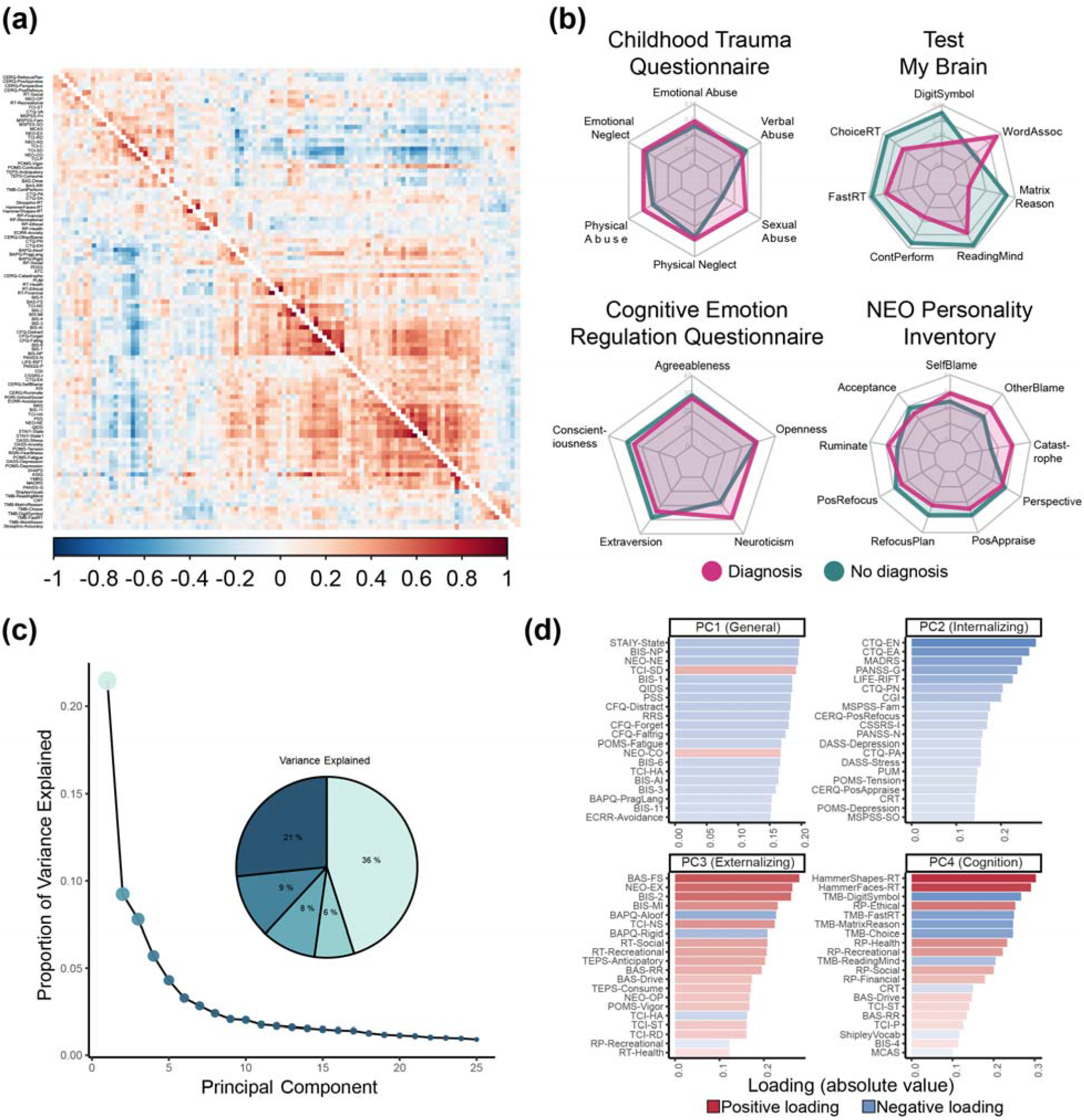
Behavioral data associations and latent structure. **(a)** Inter-participant correlation matrix between 110 scales and subscales for individuals with (top-triangle) and without (bottom-triangle) a diagnosis, with the rows and columns ordered using hierarchical clustering. **(b)** Radar plots showing scale scores differences between individuals with (pink) and without (green) a diagnosis across select measures **(c)** Screeplot and pie chart of the variance explained by the first 25 components from a PCA conducted on imputed and transformed behavioral data across all participants. **(d)** Top 30 absolute positive (red) and negative (blue) loadings on the first four principal components.

When examining select individual scales for consistency with previous findings (Fig. 7B), individuals with a mental health diagnosis, on average, had higher scores on measures of abuse on the Childhood Trauma Questionnaire^71^ and neuroticism on the NEO Five-Factor Inventory^75^. These individuals also showed higher scores on measures of catastrophizing, blame, and rumination, as indexed by the Cognitive Emotional Regulation questionnaire^58^, and worse performance across a range of computerized measures of cognitive performance^8^ from the Test My Brain battery.

In recent years, there has been a growing interest in exploring novel data-driven approaches to understanding psychopathological nosology by examining covariation among symptoms and maladaptive behaviors^156^. To examine latent dimensions of behavior that may be captured by the range of scales and subscale scores, we used Principal Component Analysis (PCA). Standard PCA cannot account for missing data and is biased by highly non-gaussian distributions, such as zero-inflated distributions often seen in clinical measures. Therefore, we imputed missing data using a simulation-based multi-algorithm comparison framework (*missCompare*^157^).

Following best practice, prior to imputation, the number of participants and measures was reduced to minimize missingness^157^. An optimal threshold for removing participants and scales was found by visualizing the proportion of missing data as a function of removing an increasing number of participants and measures and finding inflection points. This led to removing participants with >20% and variables with >25% missing data. This resulted in 191 participants and 104 measures being entered into the imputation process. Following the *missCompare* framework, 50 simulated datasets matching multivariate characteristics of the original data were generated, and then missingness patterns from the original data were added to the simulated data. Then, the missing data in each simulated dataset were imputed using a curated list of 16 algorithms^157^, and finally, the computation time and imputation accuracy of each algorithm were assessed by calculating Root Mean Square Error (RMSE), Mean Absolute Error (MAE) and Kolmogorov-Smirnov (KS) values between the imputed and the simulated data points (see Figure S4). These metrics were compared under three conditions: Missing Completely At Random (MCAR), Missing At Random (MAR), and Missing Not At Random or Non-Ignorable (MNAR). The *missForest* algorithm^158^, an iterative imputation method based on a random forest, consistently performed the best compared to all other algorithms across all metrics evaluated and was therefore used to impute the missing values in the observed data. Finally, post-imputation diagnostics, including visual examination of data distributions and stability of correlation coefficients between measures, before and after imputation, were examined, with both checks showing minimal impact of imputation. Non-gaussian measures were transformed using an optimal normalizing transformations framework (*bestNormalize*^159^), where normalization efficacy is evaluated and compared across a suite of possible transformations and evaluated for normality on goodness of fit statistics. Highly non-gaussian measures, such as those with zero inflation, were binarized.

The PCA analysis of the imputed and transformed behavioral data revealed that the first component, which accounted for 21% of the variance, represented a general functioning and well-being factor (Fig. 7C-D). This component exhibited high absolute loadings on a broad array of measures, and higher scores were associated with lower anxiety and depressive symptoms, neuroticism, impulsivity, stress, forgetfulness, fatigue, avoidance, and higher conscientiousness and self-directedness (Fig. 7D). The second, third, and fourth components were linked to internalizing, externalizing, and cognitive functioning, explaining 9%, 8%, and 6% of the variance, respectively (Fig. 7C). Higher scores on the second component were associated with lower childhood emotional and physical trauma, depressive traits and symptoms, general clinical functioning, perceived social support, neuroticism, suicidal ideation, stress, and tension (Fig. 7D). Elevated scores on the third component were related to higher extraversion, fun, novelty and pleasure-seeking, impulsiveness, social and recreational risk-taking, openness to experience and reward dependence, and lower harm avoidance, and autism traits, including rigidity and aloofness (Fig. 7D). Higher scores on the fourth component were associated with slower reaction times on multiple cognitive tasks, slower processing speed, worse abstract reasoning, memory and vocabulary, and higher concerns related to health, ethics, and recreation.

### Task-based fMRI activation and behavioral outcomes

Here, we report the performance data and brain activity estimates for the Stroop and Emotional Faces task fMRI paradigms to validate them against known effects in the literature. The Stroop task is a well-replicated assessment of inhibitory cognitive control^12^. In the Stroop task, participants must identify colors of printed words that are congruent or incongruent with the given text (see *Task fMRI paradigms* for further details). Thus, congruent trials are lower conflict and easier to identify, and incongruent trials are higher conflict and more difficult to identify. As expected, participants performed significantly better on congruent than incongruent trials both in terms of accuracy (*t*(225) = 7.59, *p* = 4.3 × 10^-13^) and reaction time (*t*(225) = -29.32, *p* = 4.3 × 10^-79^) (Fig. 8A-B). While these “Stroop effects” were statistically significant in both the diagnosis and no diagnosis groups, participants with psychiatric diagnoses exhibited less differentiated accuracy between congruent and incongruent trials (diagnoses: *t*(134) = 4.94, *p* = 1.2 × 10^-6^; no diagnoses: *t*(90) = 6.14, *p* = 1.1 × 10^-8^), but more differentiated reaction times between congruent and incongruent trials (diagnoses: *t*(134) = -23.5, *p* = 1.1 × 10^-49^; no diagnoses: *t*(90) = -17.79, *p* = 1.6 × 10^-31^). This suggests that participants with diagnoses required more time than those without diagnoses to exert inhibitory cognitive control during incongruent trials (with reference to performance on congruent trials).

**Figure 8.**
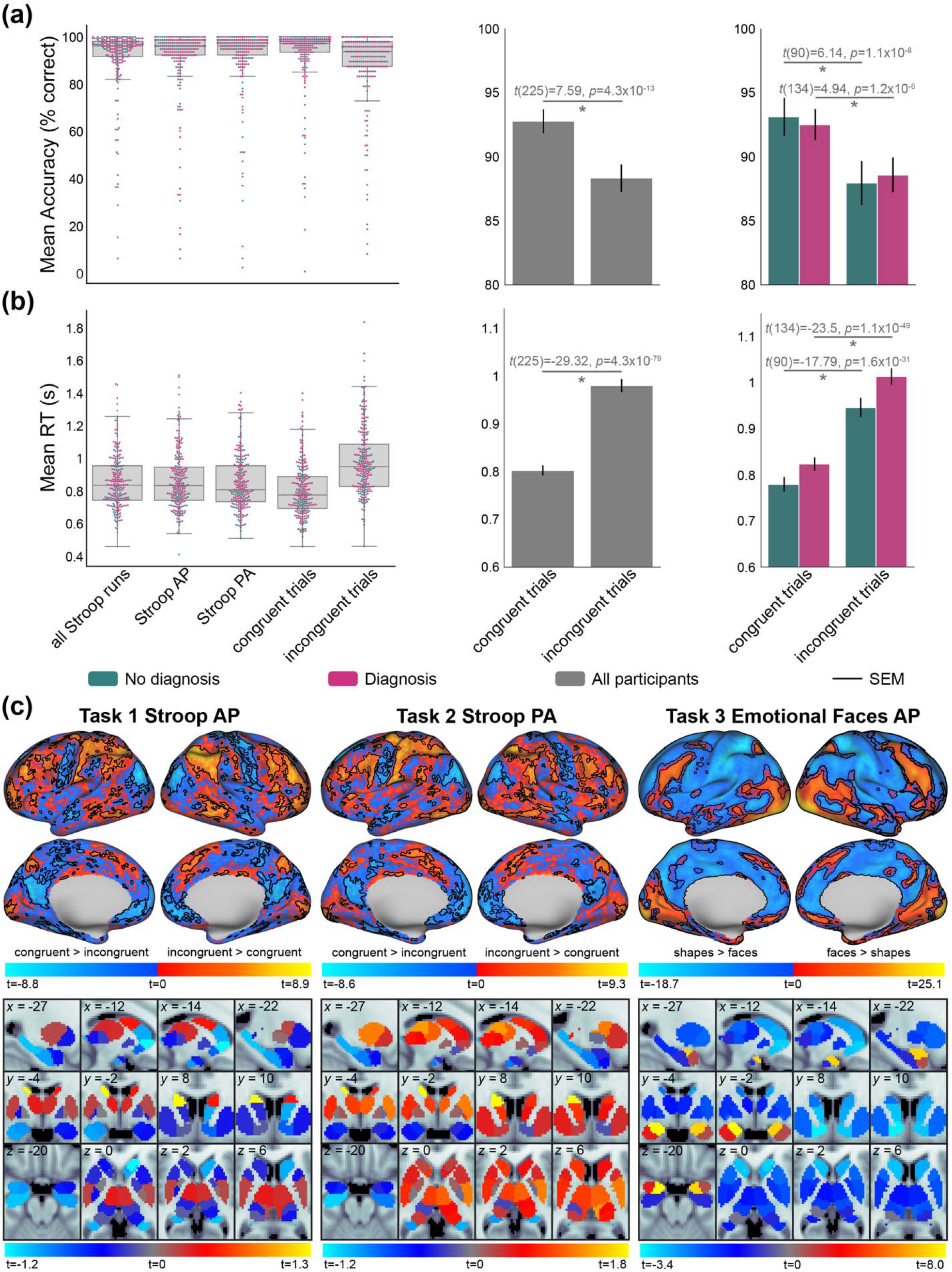
Functional task-activation and behavioral data from Stroop and Emotional Faces tasks. **(a)** Stroop task performance quantified via percentage of correct responses. *Left*: across-participant distribution of accuracy scores for all trials and runs, trials in each AP and PA functional run, and trials in each of the congruent and incongruent conditions. Lines inside boxplots indicate median performance, and dots indicate individual participant scores. For each figure, participants with diagnoses are coded with pink, and without diagnoses coded with turquoise, as applicable. *Middle*: Paired t-test of congruent versus incongruent accuracies across all participants. *Right*: The same as the middle panel, but for the diagnosis and no diagnosis groups. SEM: standard error of the mean. **(b)** Same as panel a, but for the performance metric of reaction time (seconds). **(c)** GLM-based planned contrasts for functional neuroimaging data for the Stroop (*left*: AP; *middle*: PA) and Emotional Faces (*right*) tasks. Cool and warm color scales show complementary contrasts, overlaid together on one brain schematic. For example, for the two Stroop runs, brain activity that was greater for congruent versus incongruent trials in cool color scale, and incongruent greater than congruent trials in warm color scale. Results for cortical vertices are projected onto a surface-based brain schematic (black borders surround vertices whose contrasts passed FDR correction). Subcortical results are projected onto a standard MNI (2 mm) volume-based brain schematic, showing average contrast statistics for each of the 32 regions provided by Tian et al.^115^. Three slices are shown following the visualizations in Tian et al.^115^, plus an additional slice to highlight the amygdala (x=-27, y=-4, z=-20). The group-level statistical activation maps are shown in all projections.

Next, we performed task GLMs on the functional neuroimaging data acquired during these task states (Fig. 8C). For the Stroop GLM, we used the congruent and incongruent conditions as regressors, and for the Emotional Faces GLM, we used face and shape conditions as regressors. For each participant, task events were time-locked to the onset of each trial and convolved with a canonical hemodynamic response function (HRF^160^), as well as a parameter for the first temporal derivative of the HRF. In addition, 8 parameters accounting for potential low-frequency signals (i.e., drift; with a cutoff of 0.01 Hz) that used a discrete cosine transform were added, as well as a constant parameter. We examined correct trials only and specifically contrasted the aforementioned conditions of interest (e.g., incongruent > congruent) for each vertex across the whole brain. The resulting individual-level statistical maps were compared using one-sample t-testing (versus 0) across participants to obtain group-level statistical maps (shown in Fig. 8). We used false discovery rate (FDR) correction for multiple comparisons^161^ with a family-wise error rate of alpha=0.05. Vertices that passed FDR correction are outlined in black borders in Fig. 8.

As shown in prior work, brain regions in the cingulate gyrus and lateral prefrontal cortex were significantly more responsive to incongruent versus congruent trials during the Stroop task^162,163^. Additionally, regions distributed across functional systems were more responsive to incongruent trials, including the frontoparietal, dorsal/ventral attention, somatomotor, and visual networks. In the Emotional Faces task, subcortical vertices in the amygdala exhibited a robust and selective responsiveness to emotional faces versus shapes. This is consistent with the literature which reports that the Emotional Faces task reliably activates the amygdala in response to affective images of human faces^10,164^. These results validate that the Stroop and Emotional Faces task fMRI paradigms activate brain regions and systems previously shown to be involved with cognitive control and emotional reactivity, respectively. We performed these analyses across all participants, but we encourage future research with the TCP dataset to examine the extent to which brain activity and network interactions may be differentiable with diagnostic status.

## Usage Notes

Transdiagnostic neuroimaging datasets with a broad range of behavioral measures are necessary to address complex questions regarding the relation between brain and behavior in psychiatry. The TCP release provided a curated collection of neuroimaging, behavioral, cognitive, and personality data from 241 individuals meeting diagnostic criteria for a broad range of disorders, as well as individuals who do not meet these diagnostic thresholds (i.e., healthy controls). The data collection provides both processed and analysis-ready neuroimaging data using HCP-validated processing pipelines, as well as raw and anonymized BIDS-formatted data to allow researchers to implement alternate processing. Raw neuroimaging data and all behavioral measures can be accessed via OpenNeuro (https://openneuro.org/datasets/ds005237). Raw and processed neuroimaging data, as well as all behavioral measures, can be openly accessed via the NDA (https://nda.nih.gov/edit_collection.html?id=3552) upon publication.

## Code Availability

The HCP processing pipelines are openly available here: https://github.com/Washington-University/HCPpipelines. All network metric code is available here: https://github.com/aestrivex/bctpy (Python) and here: https://sites.google.com/site/bctnet/ (MATLAB). All other code used for post-processing, FC estimation, and quality assurance analyses are available here: https://github.com/HolmesLab/TransdiagnosticConnectomeProject.

## Data Availability

Raw neuroimaging data and all behavioral measures can be accessed via OpenNeuro (https://openneuro.org/datasets/ds005237). Raw and processed neuroimaging data, as well as all behavioral measures, can be openly accessed via the NDA (https://nda.nih.gov/edit_collection.html?id=3552) upon publication.

## Acknowledgments

This work was supported by the National Institute of Mental Health (R01MH123245 to AJH and R01MH120080 to AJH and BTTY). BTTY is also supported by the NUS Yong Loo Lin School of Medicine (NUHSRO/2020/124/TMR/LOA), the Singapore National Medical Research Council (NMRC) LCG (OFLCG19May-0035), NMRC CTG-IIT (CTGIIT23jan-0001), NMRC STaR (STaR20nov-0003), Singapore Ministry of Health (MOH) Centre Grant (CG21APR1009), the Temasek Foundation (TF2223-IMH-01), and the United States National Institutes of Health (R01MH120080). Any opinions, findings and conclusions or recommendations expressed in this material are those of the authors and do not reflect the views of the grantors. SC is supported by an American Australian Association Graduate Education Fund Scholarship. JAR is supported by the Stanford University Knight-Hennessy Scholars Program, and the National Academies of Sciences, Engineering, and Medicine’s Ford Foundation Predoctoral Fellowship. LMP is supported by a T32 Fellowship from the National Institute of Mental Health (T32MH019112-32).

## Supplementary information

**Table S1.** Summary metrics for scales and subscales.

| Variable | Missing (%) | Median | Mean | Min | Max |
| --- | --- | --- | --- | --- | --- |
| ASC | 58.70 | 251.00 | 434.01 | 0.00 | 2,050.00 |
| ASI | 10.12 | 15.50 | 17.53 | 0.00 | 59.00 |
| ATC | 9.72 | 10.00 | 9.96 | 3.00 | 24.00 |
| BAPQ-Aloof | 14.57 | 2.75 | 2.83 | 1.17 | 5.92 |
| BAPQ-PragLang | 14.57 | 2.67 | 2.76 | 1.17 | 4.17 |
| BAPQ-Rigid | 14.57 | 3.00 | 3.02 | 1.42 | 4.75 |
| BAS-Drive | 9.31 | 11.00 | 10.83 | 4.00 | 16.00 |
| BAS-FS | 9.31 | 11.00 | 11.27 | 5.00 | 16.00 |
| BAS-RR | 9.31 | 17.00 | 16.93 | 8.00 | 20.00 |
| BIS-1 | 16.19 | 9.00 | 9.82 | 5.00 | 20.00 |
| BIS-11 | 9.31 | 21.00 | 20.74 | 7.00 | 28.00 |
| BIS-2 | 16.19 | 13.00 | 13.94 | 7.00 | 25.00 |
| BIS-3 | 16.19 | 11.00 | 11.61 | 6.00 | 21.00 |
| BIS-4 | 16.19 | 10.00 | 10.41 | 5.00 | 17.00 |
| BIS-5 | 16.19 | 7.00 | 7.37 | 4.00 | 14.00 |
| BIS-6 | 16.19 | 5.00 | 5.80 | 3.00 | 11.00 |
| BIS-AI | 16.19 | 22.00 | 22.01 | 12.00 | 35.00 |
| BIS-MI | 16.19 | 21.00 | 21.31 | 13.00 | 34.00 |
| BIS-NP | 16.19 | 15.00 | 15.62 | 8.00 | 30.00 |
| CERQ-Acceptance | 9.31 | 8.00 | 7.25 | 2.00 | 10.00 |
| CERQ-Catastrophe | 9.31 | 5.00 | 5.02 | 2.00 | 10.00 |
| CERQ-OtherBlame | 9.31 | 4.00 | 4.12 | 2.00 | 9.00 |
| CERQ-Perspective | 9.31 | 7.00 | 6.69 | 2.00 | 10.00 |
| CERQ-PosAppraise | 9.31 | 8.00 | 7.63 | 2.00 | 10.00 |
| CERQ-PosRefocus | 9.31 | 5.00 | 5.14 | 2.00 | 10.00 |
| CERQ-RefocusPlan | 9.31 | 8.00 | 7.65 | 2.00 | 10.00 |
| CERQ-Ruminate | 9.31 | 7.00 | 6.64 | 2.00 | 10.00 |
| CERQ-SelfBlame | 9.31 | 7.00 | 6.47 | 2.00 | 10.00 |
| CFQ-Distract | 8.10 | 13.00 | 12.86 | 0.00 | 30.00 |
| CFQ-Faltrig | 8.10 | 9.00 | 9.73 | 0.00 | 27.00 |
| CFQ-Forget | 8.10 | 13.00 | 14.21 | 0.00 | 31.00 |
| CGI | 7.29 | 1.00 | 1.13 | 0.50 | 3.50 |
| CRT | 5.26 | 1.00 | 1.26 | 0.00 | 3.00 |
| CSSRS-I | 4.05 | 0.00 | 5.98 | 0.00 | 24.00 |
| CTQ-EA | 20.65 | 8.88 | 9.95 | 5.00 | 25.00 |
| CTQ-EN | 20.65 | 10.00 | 10.70 | 5.00 | 25.00 |
| CTQ-PA | 20.65 | 5.00 | 7.07 | 5.00 | 23.00 |
| CTQ-PN | 20.65 | 6.00 | 7.43 | 5.00 | 20.00 |
| CTQ-SA | 20.65 | 5.00 | 7.04 | 5.00 | 25.00 |
| CTQ-VA | 20.65 | 0.00 | 0.40 | 0.00 | 3.00 |
| DASS-Anxiety | 6.48 | 3.00 | 5.36 | 0.00 | 32.00 |
| DASS-Depression | 6.48 | 3.00 | 7.97 | 0.00 | 40.00 |
| DASS-Stress | 6.48 | 6.00 | 8.75 | 0.00 | 39.00 |
| ECRR-Anxiety | 9.72 | 3.82 | 3.83 | 2.61 | 5.12 |
| ECRR-Avoidance | 9.72 | 3.83 | 3.87 | 1.67 | 5.78 |
| FTND | 6.07 | 0.00 | 0.25 | 0.00 | 7.00 |
| HammerFaces-RT | 9.72 | 828.27 | 837.08 | 520.35 | 1,619.70 |
| HammerShapes-RT | 9.72 | 733.32 | 748.47 | 458.36 | 1,468.65 |
| LIFE-RIFT | 14.98 | 6.00 | 6.99 | 4.00 | 19.00 |
| MADRS | 6.07 | 3.00 | 7.15 | 0.00 | 39.00 |
| MCAS | 7.69 | 84.00 | 81.96 | 59.00 | 89.00 |
| MSPSS-Fam | 12.15 | 5.50 | 5.22 | 1.00 | 7.00 |
| MSPSS-Fri | 12.15 | 6.00 | 5.61 | 1.00 | 7.00 |
| MSPSS-SO | 12.15 | 6.00 | 5.44 | 1.00 | 7.00 |
| NEO-AG | 17.00 | 35.00 | 34.18 | 17.00 | 48.00 |
| NEO-CO | 17.00 | 33.00 | 32.80 | 6.00 | 48.00 |
| NEO-EX | 17.00 | 27.00 | 27.40 | 3.00 | 45.00 |
| NEO-NE | 18.22 | 22.00 | 22.62 | 1.00 | 45.00 |
| NEO-OP | 17.00 | 32.00 | 30.87 | 12.00 | 45.00 |
| PANSS-G | 7.29 | 19.00 | 21.10 | 16.00 | 47.00 |
| PANSS-N | 6.88 | 7.00 | 9.22 | 7.00 | 28.00 |
| PANSS-P | 7.29 | 7.00 | 8.21 | 7.00 | 30.00 |
| PDSS | 12.15 | 0.00 | 0.23 | 0.00 | 6.00 |
| POMS-Confusion | 10.12 | 40.00 | 41.52 | 33.00 | 67.00 |
| POMS-Depression | 10.12 | 38.00 | 39.79 | 32.00 | 67.00 |
| POMS-Fatigue | 10.12 | 41.00 | 45.21 | 30.00 | 76.00 |
| POMS-Tension | 10.12 | 36.00 | 38.33 | 30.00 | 61.00 |
| POMS-Vigor | 10.12 | 52.00 | 53.17 | 36.00 | 80.00 |
| PSS | 5.67 | 15.00 | 15.86 | 0.00 | 37.00 |
| PUM | 10.53 | 19.00 | 22.14 | 14.00 | 56.00 |
| QIDS | 19.03 | 4.00 | 5.32 | 0.00 | 22.00 |
| RP-Ethical | 24.70 | 37.00 | 36.66 | 18.00 | 56.00 |
| RP-Financial | 24.70 | 39.00 | 38.58 | 8.00 | 56.00 |
| RP-Health | 24.70 | 39.00 | 38.98 | 13.00 | 56.00 |
| RP-Recreational | 24.70 | 35.50 | 35.01 | 8.00 | 53.00 |
| RP-Social | 24.70 | 23.00 | 23.68 | 8.00 | 53.00 |
| RRS | 9.72 | 46.00 | 47.45 | 22.00 | 86.00 |
| RSRI-FearIllness | 8.91 | 1.83 | 1.95 | 1.00 | 3.67 |
| RSRI-SchoolSocial | 8.91 | 2.50 | 2.44 | 1.08 | 4.42 |
| RT-Ethical | 24.70 | 18.00 | 18.58 | 8.00 | 37.00 |
| RT-Financial | 24.70 | 18.00 | 18.58 | 8.00 | 37.00 |
| RT-Health | 24.70 | 22.00 | 23.15 | 8.00 | 51.00 |
| RT-Recreational | 24.70 | 27.00 | 27.63 | 8.00 | 56.00 |
| RT-Social | 24.70 | 39.00 | 38.06 | 14.00 | 56.00 |
| SHAPS | 12.55 | 0.00 | 1.27 | 0.00 | 12.00 |
| STAIY-State | 7.69 | 38.00 | 39.68 | 20.00 | 73.00 |
| STAIY-State1 | 7.69 | 38.00 | 39.68 | 20.00 | 73.00 |
| ShipleyVocab | 4.45 | 57.00 | 55.97 | 30.00 | 71.00 |
| StroopInc-Accuracy | 25.91 | 46.00 | 44.10 | 18.00 | 48.00 |
| StroopInc-RT | 25.91 | 901.59 | 913.59 | 500.32 | 1,524.47 |
| TCI-C | 24.29 | 81.00 | 79.67 | 44.00 | 98.00 |
| TCI-HA | 24.29 | 57.00 | 57.51 | 24.00 | 95.00 |
| TCI-NS | 24.29 | 53.00 | 53.90 | 30.00 | 86.00 |
| TCI-P | 24.29 | 72.00 | 71.24 | 25.00 | 96.00 |
| TCI-RD | 24.29 | 69.00 | 69.14 | 41.00 | 96.00 |
| TCI-SD | 24.29 | 76.00 | 74.58 | 46.00 | 98.00 |
| TCI-ST | 24.29 | 39.00 | 39.96 | 17.00 | 75.00 |
| TEPS-Anticipatory | 6.88 | 44.00 | 43.29 | 11.00 | 59.00 |
| TEPS-Consume | 6.88 | 39.00 | 38.10 | 8.00 | 48.00 |
| TMB-Choice | 12.15 | 11.63 | 11.94 | 3.50 | 47.60 |
| TMB-ContPerform | 9.72 | 71.87 | 68.20 | 6.25 | 96.87 |
| TMB-DigitSymbol | 12.55 | 49.00 | 49.02 | 17.00 | 147.00 |
| TMB-FastRT | 12.15 | 33.16 | 33.00 | 18.45 | 49.45 |
| TMB-MatrixReason | 11.74 | 24.00 | 22.26 | 4.00 | 34.00 |
| TMB-ReadingMind | 12.96 | 25.00 | 24.07 | 8.00 | 34.00 |
| TMB-WordAssoc | 12.96 | 48.00 | 57.69 | 13.00 | 100.00 |
| YMRS | 7.29 | 0.00 | 1.12 | 0.00 | 34.00 |

**Figure S1.**
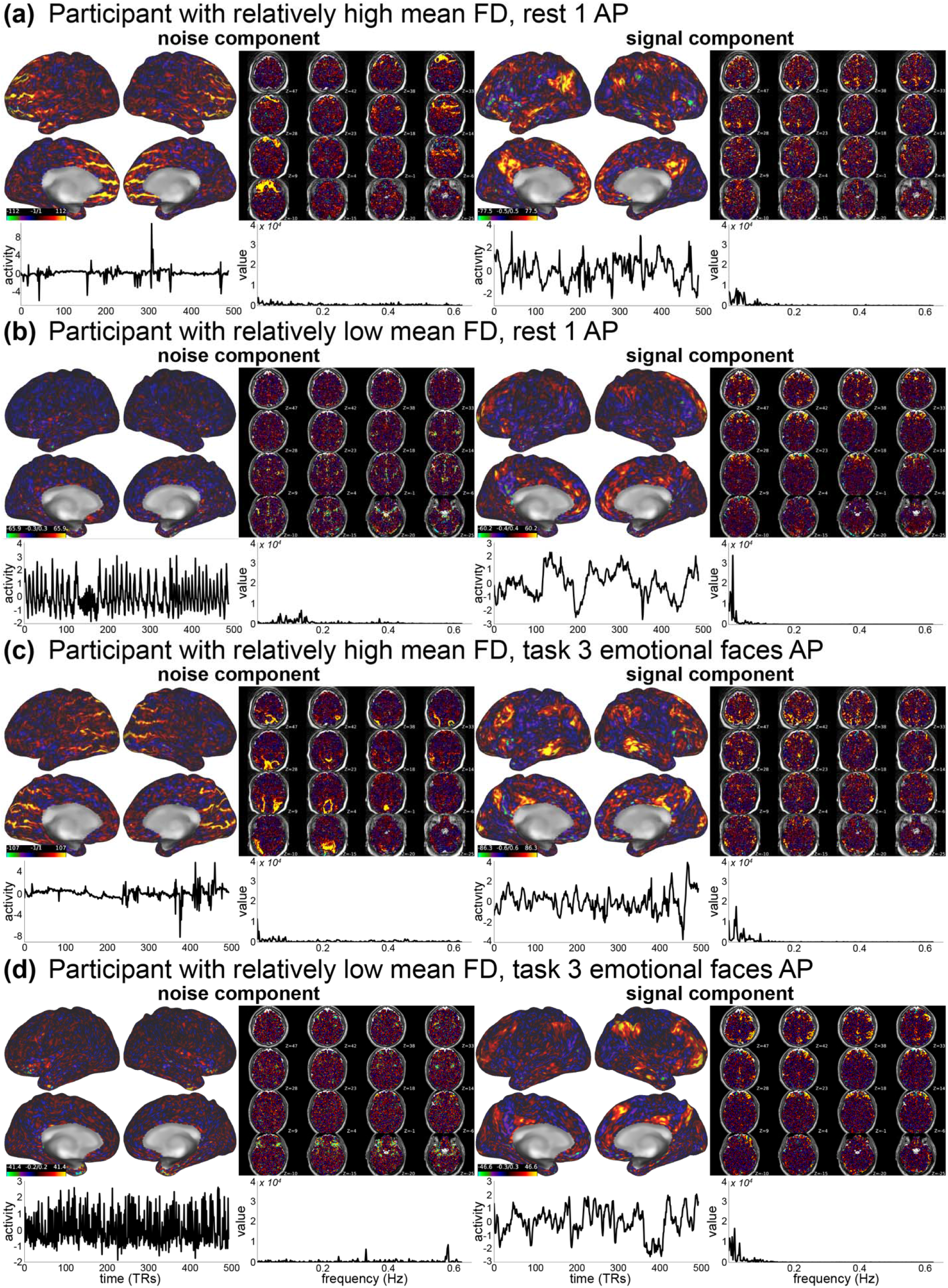
Example quality assurance of ICA-FIX. **(a)** Example connectome workbench scenes provided by the HCP processing step “PostFix”, which allows researchers to perform quality checks on the classification given by ICA-FIX denoising. This panel shows one example participant who exhibited relatively high FD across resting-state run 1 AP TRs. *Left panel*: example noise component; *right panel*: example signal component. In each of these panels the top left and right show montages of this component on the surface and across various axial volume slices, respectively. The bottom left is the component across the timeseries, and the bottom right is the corresponding power spectrum. **(b)** The same as panel a, but for a participant who exhibited relatively low FD across resting-state run 1 AP. **(c)** and **(d)** are the same as panels a and b, respectively, but for the Emotional Faces fMRI task. In each case, signal components show prototypical timeseries with power spectrum mainly exhibited in low frequencies, while noise components either contain “spikes’’ with respect to the timeseries (as in panel a, left), or exhibit waveforms with classical random “noisiness” (as in panel d, left). The noise components for panels a and c are also clearly evident on the surface via unusual banding patterns across select portions of cortex, whereas signal components appear to map onto functional systems more smoothly.

**Figure S2.**
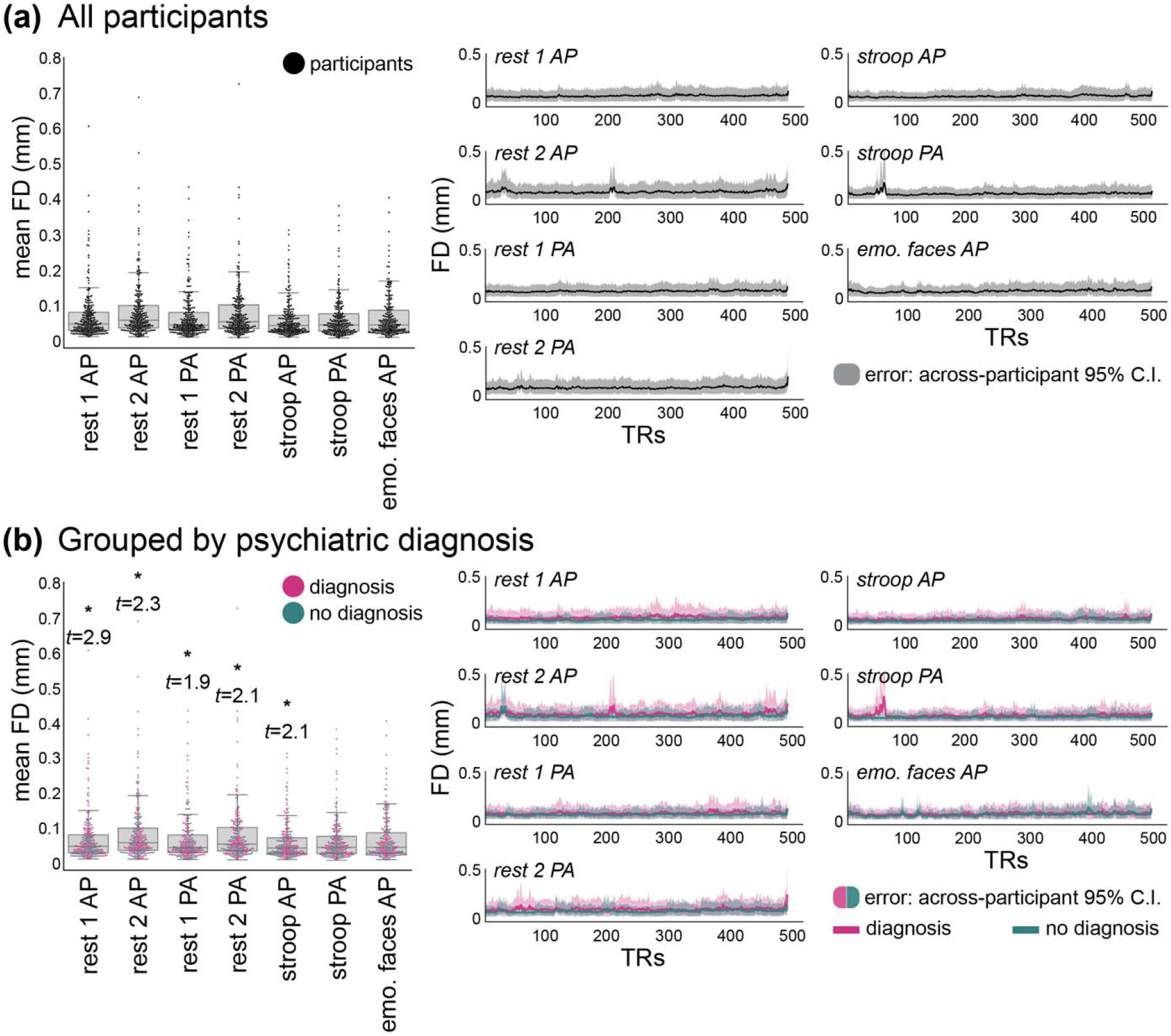
Mean-filtered and down-sampled framewise displacement (FD) for each fMRI run. **(a)** *Left*: boxplots of the FD (in mm) indicate across-region and across-run averages exhibited by each participant (black dots). *Right*: FD traces across the entire functional timeseries (in TRs) for each run. Black lines indicate across-participant averages; shaded gray portions indicate across-participant confidence intervals (95%). Panel a shows all TCP participants. **(b)** Same as panel a but grouping TCP participants by those with a psychiatric diagnosis (magenta) and those without a psychiatric diagnosis (turquoise). Asterisks on the left-most boxplots indicate that those with diagnoses exhibited significantly higher FD than those without diagnoses for that functional run, as given by Welch’s t-test for independent samples with unequal variances. The statistics for all comparisons were as follows: (1) rest AP, run 1: *t*(223.16) = 2.89, *p* = 0.002; (2) rest AP, run 2: *t*(227.74) = 2.32, *p* = 0.011; (3) rest PA, run 1: *t*(236.69) = 1.92, *p* = 0.028; (4) rest PA, run 2: t(230.6) = 2.05, p = 0.021; (5) task 1, Stroop AP: *t*(222.65) = 2.12, *p* = 0.018; (6) task 3, Stroop PA: *t*(143.09) = 1.47, *p* = 0.072; (7) task 3, Emotional Faces AP: *t*(217.59) = 1.2, *p* = 0.116. While the impact of motion was well-controlled for all participants, those exhibiting the largest FD were participants with diagnoses (refer to outlier dots on the left and t-test results, as well as relatively more magenta “spikes” on right FD traces), consistent with the literature. All panels show FD of timeseries data that was denoised with ICA-FIX and had GSR applied.

**Figure S3.**
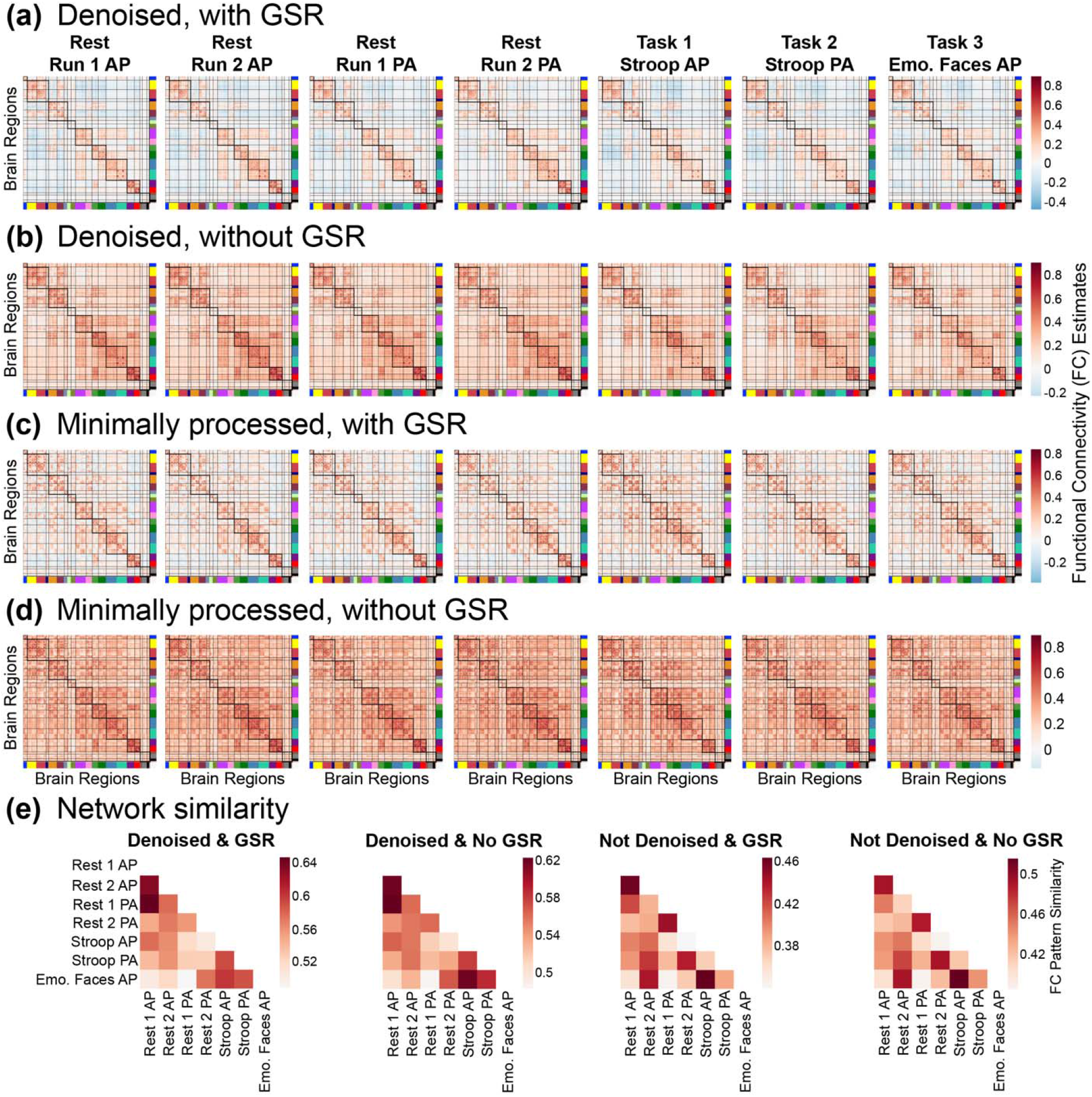
Group-average functional connectivity matrices at different stages of processing. **(a)** The same connectomes shown in Fig. 5; timeseries data was denoised with ICA-FIX and GSR was applied. **(b)** Timeseries data was denoised, but no GSR was applied. Note the global consistency in network structure to panel a, but a shifted range for connectivity estimates. As shown in prior work, GSR improved the separation of within-network versus between-network connectivity patterns. **(c)** Timeseries data were minimally processed with the HCP pipelines, but denoising steps were not performed. Here, GSR was applied. **(d)** Timeseries did not have denoising nor GSR applied. Note that while the gross network structure was still apparent, the absence of denoising (panels c and d) introduced mini-clusters of connectivity patterns dispersed throughout the network; these are likely spurious. **(e)** The same network similarity (given by Mantel r) results shown in Fig. 5, but for all four processing variants in panels a through d. The pattern of network similarity results was consistent for the approaches with denoising, with and without GSR. However, the absence of denoising (two right-most heatmaps) yielded some inconsistencies. Resting-state 2 AP exhibited increased similarity with the Emotional Faces task and Resting-state 2 PA exhibited increased similarity with the Stroop AP task; these increases are also likely spurious. Taken together, we recommend use of functional connectivity estimates with denoising, and with or without GSR depending on the research question (i.e., panel a or b).

**Figure S4.**
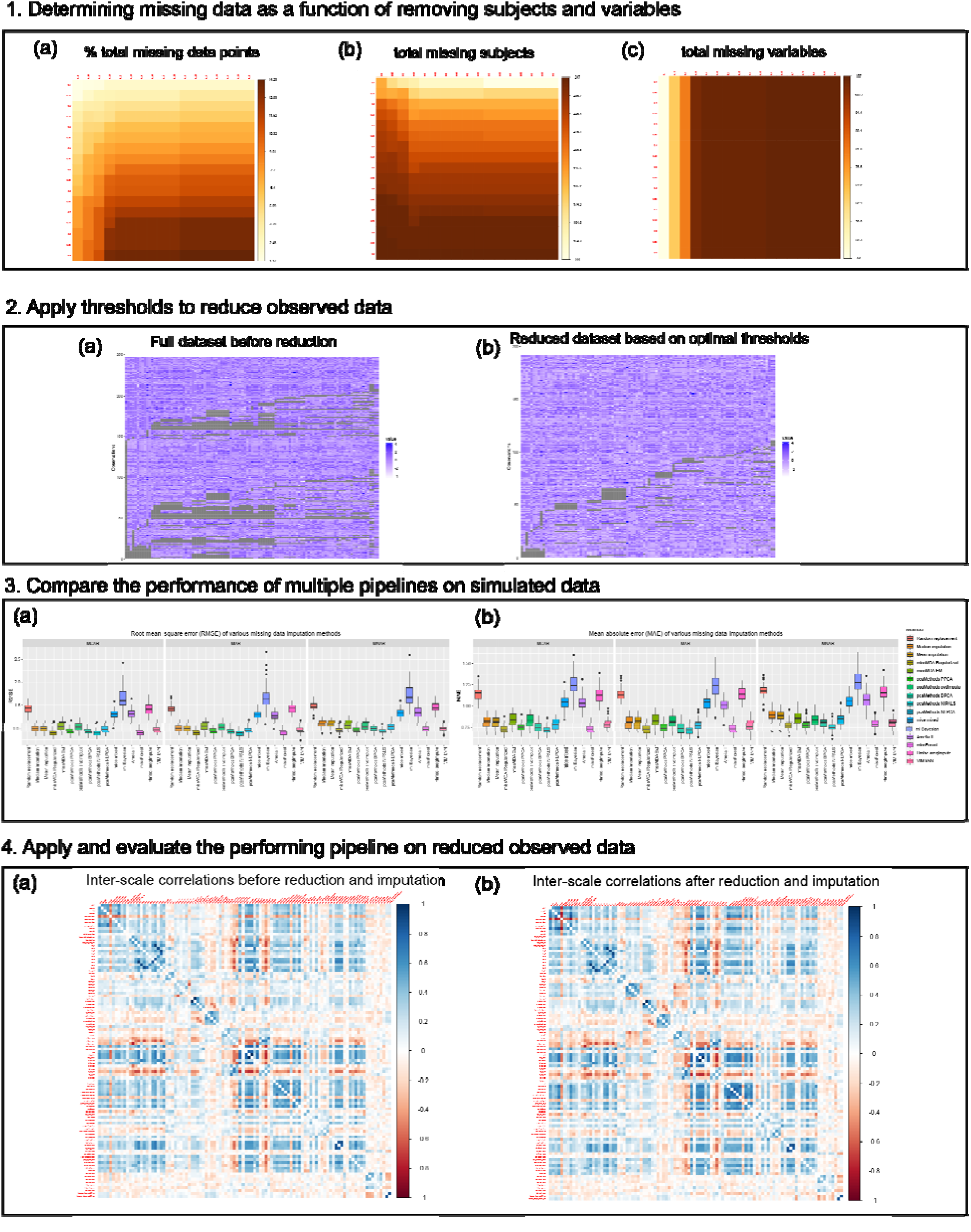
Imputation pipeline for behavioral data prior to dimensionality reduction. **1)** Optimal thresholds for removing participants and variables with excessive missing data prior to imputation were computed based on inflection points that retained the most data across total data points, participants, and variables, while reducing overall missingness. **2)** This resulted removing participants with >20% and variables with >25% missing data. This resulted in 191 participants and 104 measures entering the imputation process. **3)** Using the missCompare package, we compared the performance of 16 different imputation pipelines on 50 simulated version of the observed data under three conditions: Missing Completely At Random (MCAR), Missing At Random (MAR) and Missing Not At Random or Non-Ignorable (MNAR). **4)** The missForrest pipeline showed the best performance and was used to impute data in the reduced observed dataset. Distributions and correlations between scales were compared before and after imputation to ensure imputation quality.

## Notes

### Competing Interest Statement

The authors have declared no competing interest.

### Author Declarations

IRB of both Yale University and McLean Hospital (Partners Healthcare) gave ethical approval for this work. Representative study consent forms from each site are provided in Supplementary Appendix A.

