## Appendix for "The Transdiagnostic Connectome Project: a richly phenotyped open dataset for advancing the study of brain-behavior relationships in psychiatry": CMRR Holmes_Yale_McLean_Jan2022.pdf

**Table of contents**

\\USER

Holmes

PCY

CMRR Holmes_Yale_McLean_Jan2022

Localizer
AAHScout
Localizer_aligned
T1_vNav_setter
T1_MEMPRAGE_GSP_vNavTrk
T2_vNav_setter
T2_SPACE_GSP_vNavTrk
SpinEchoFieldMap_AP
SpinEchoFieldMap_PA
rfMRI_REST_AP
rfMRI_REST_PA
Stroop_Bl1_AP
Stroop_Bl2_PA
Hammer_Bl1_AP
rfMRI_REST_AP
rfMRI_REST_PA

**\\USER\Holmes\PCY\CMRR Holmes_Yale_McLean_Jan2022\Localizer**

TA: 9.2 s PM: REF Voxel size: 1.2×1.2×5.0 mmPAT: Off Rel. SNR: 1.00 : fl

### Properties

| | |
|---|---|
| Prio recon | On |
| Load images to viewer | Off |
| Inline movie | Off |
| Auto store images | On |
| Load images to stamp segments | On |
| Load images to graphic segments | On |
| Auto open inline display | Off |
| Auto close inline display | Off |
| Start measurement without further preparation | On |
| Wait for user to start | On |
| Start measurements | Single measurement |

### Routine

| | |
|---|---|
| Slice group | 1 |
| Slices | 1 |
| Dist. factor | 20 % |
| Position | L0.0 A20.0 H0.0 mm |
| Orientation | Transversal |
| Phase enc. dir. | A >> P |
| Slice group | 2 |
| Slices | 1 |
| Dist. factor | 20 % |
| Position | L0.0 A20.0 H0.0 mm |
| Orientation | Sagittal |
| Phase enc. dir. | A >> P |
| Slice group | 3 |
| Slices | 1 |
| Dist. factor | 20 % |
| Position | L0.0 A20.0 H0.0 mm |
| Orientation | Coronal |
| Phase enc. dir. | R >> L |
| AutoAlign | --- |
| Phase oversampling | 0 % |
| FoV read | 300 mm |
| FoV phase | 100.0 % |
| Slice thickness | 5.0 mm |
| TR | 40.0 ms |
| TE | 3.00 ms |
| Averages | 1 |
| Concatenations | 1 |
| Filter | Prescan Normalize, Elliptical filter |
| Coil elements | HC1-7;NC1,2 |

### Contrast - Common

| | |
|---|---|
| TR | 40.0 ms |
| TE | 3.00 ms |
| MTC | Off |
| Magn. preparation | None |
| Flip angle | 15 deg |
| Fat suppr. | None |
| Water suppr. | None |

### Contrast - Dynamic

| | |
|---|---|
| Averages | 1 |
| Averaging mode | Short term |
| Reconstruction | Magnitude |
| Measurements | 1 |
| Multiple series | Off |

### Resolution - Common

| | |
|---|---|
| FoV read | 300 mm |
| FoV phase | 100.0 % |
| Slice thickness | 5.0 mm |
| Base resolution | 256 |
| Phase resolution | 75 % |
| Phase partial Fourier | Off |
| Interpolation | Off |

### Resolution - iPAT

| | |
|---|---|
| PAT mode | None |

### Resolution - Filter Image

| | |
|---|---|
| Image Filter | Off |
| Distortion Corr. | Off |
| Prescan Normalize | On |
| Unfiltered images | Off |
| Normalize | Off |
| B1 filter | Off |

### Resolution - Filter Rawdata

| | |
|---|---|
| Raw filter | Off |
| Elliptical filter | On |

### Geometry - Common

| | |
|---|---|
| Slice group | 1 |
| Slices | 1 |
| Dist. factor | 20 % |
| Position | L0.0 A20.0 H0.0 mm |
| Orientation | Transversal |
| Phase enc. dir. | A >> P |
| Slice group | 2 |
| Slices | 1 |
| Dist. factor | 20 % |
| Position | L0.0 A20.0 H0.0 mm |
| Orientation | Sagittal |
| Phase enc. dir. | A >> P |
| Slice group | 3 |
| Slices | 1 |
| Dist. factor | 20 % |
| Position | L0.0 A20.0 H0.0 mm |
| Orientation | Coronal |
| Phase enc. dir. | R >> L |
| FoV read | 300 mm |
| FoV phase | 100.0 % |
| Slice thickness | 5.0 mm |
| TR | 40.0 ms |
| Multi-slice mode | Interleaved |
| Series | Interleaved |
| Concatenations | 1 |

### Geometry - AutoAlign

| | |
|---|---|
| Slice group | 1 |
| Position | L0.0 A20.0 H0.0 mm |
| Orientation | Transversal |
| Phase enc. dir. | A >> P |
| Slice group | 2 |
| Position | L0.0 A20.0 H0.0 mm |
| Orientation | Sagittal |
| Phase enc. dir. | A >> P |
| Slice group | 3 |

### Geometry - AutoAlign

| | |
|---|---|
| Position | L0.0 A20.0 H0.0 mm |
| Orientation | Coronal |
| Phase enc. dir. | R >> L |
| AutoAlign | --- |
| Initial Position | L0.0 A20.0 H0.0 |
| L | 0.0 mm |
| A | 20.0 mm |
| H | 0.0 mm |
| Initial Rotation | 0.00 deg |
| Initial Orientation | Transversal |

### Geometry - Saturation

| | |
|---|---|
| Saturation mode | Standard |
| Fat suppr. | None |
| Water suppr. | None |
| Special sat. | None |

### System - Miscellaneous

| | |
|---|---|
| Positioning mode | REF |
| Table position | H |
| Table position | 0 mm |
| MSMA | S - C - T |
| Sagittal | R >> L |
| Coronal | A >> P |
| Transversal | F >> H |
| Coil Combine Mode | Sum of Squares |
| Save uncombined | Off |
| Matrix Optimization | Off |
| AutoAlign | --- |
| Coil Select Mode | Off - All |

### System - Adjustments

| | |
|---|---|
| B0 Shim mode | Tune up |
| B1 Shim mode | TrueForm |
| Adjust with body coil | Off |
| Confirm freq. adjustment | Off |
| Assume Dominant Fat | Off |
| Assume Silicone | Off |
| Adjustment Tolerance | Auto |

### System - Adjust Volume

| | |
|---|---|
| Position | Isocenter |
| Orientation | Transversal |
| Rotation | 0.00 deg |
| A >> P | 263 mm |
| R >> L | 350 mm |
| F >> H | 350 mm |
| Reset | Off |

### System - pTx Volumes

| | |
|---|---|
| B1 Shim mode | TrueForm |
| Excitation | Slice-sel. |

### System - Tx/Rx

| | |
|---|---|
| Frequency 1H | 123.260801 MHz |
| Correction factor | 1 |
| Gain | High |
| Img. Scale Cor. | 1.000 |
| Reset | Off |
| ? Ref. amplitude 1H | 0.000 V |

### Physio - Signal1

| | |
|---|---|
| 1st Signal/Mode | None |

### Physio - Signal1

| | |
|---|---|
| TR | 40.0 ms |
| Concatenations | 1 |
| Segments | 1 |

### Physio - Cardiac

| | |
|---|---|
| Magn. preparation | None |
| Fat suppr. | None |
| Dark blood | Off |
| FoV read | 300 mm |
| FoV phase | 100.0 % |
| Phase resolution | 75 % |

### Physio - PACE

| | |
|---|---|
| Resp. control | Off |
| Concatenations | 1 |

### Inline - Common

| | |
|---|---|
| Subtract | Off |
| Measurements | 1 |
| StdDev | Off |
| Liver registration | Off |
| Save original images | On |

### Inline - MIP

| | |
|---|---|
| MIP-Sag | Off |
| MIP-Cor | Off |
| MIP-Tra | Off |
| MIP-Time | Off |
| Save original images | On |

### Inline - Soft Tissue

| | |
|---|---|
| Wash - In | Off |
| Wash - Out | Off |
| TTP | Off |
| PEI | Off |
| MIP - time | Off |
| Measurements | 1 |

### Inline - Composing

| | |
|---|---|
| Distortion Corr. | Off |

### Inline - MapIt

| | |
|---|---|
| Save original images | On |
| MapIt | None |
| Flip angle | 15 deg |
| Measurements | 1 |
| Contrasts | 1 |
| TR | 40.0 ms |
| TE | 3.00 ms |

### Sequence - Part 1

| | |
|---|---|
| Introduction | On |
| Dimension | 2D |
| Phase stabilisation | Off |
| Asymmetric echo | Allowed |
| Contrasts | 1 |
| Flow comp. | No |
| Multi-slice mode | Interleaved |
| Bandwidth | 260 Hz/Px |

### Sequence - Part 2

| | |
|---|---|
| Segments | 1 |
| Acoustic noise reduction | None |

### Sequence - Part 2

| | |
|---|---|
| RF pulse type | Normal |
| Gradient mode | Fast |
| Excitation | Slice-sel. |
| RF spoiling | On |

### Sequence - Assistant

| | |
|---|---|
| Mode | Off |
| Allowed delay | 0 s |

**\\USER\Holmes\PCY\CMRR Holmes_Yale_McLean_Jan2022\AAHScout**

TA: 0:14 PM: REF Voxel size: 1.6×1.6×1.6 mmPAT: 3 Rel. SNR: 1.00 : fl

### Properties

| | |
|---|---|
| Prio recon | On |
| Load images to viewer | On |
| Inline movie | Off |
| Auto store images | On |
| Load images to stamp segments | Off |
| Load images to graphic segments | Off |
| Auto open inline display | Off |
| Auto close inline display | Off |
| Start measurement without further preparation | On |
| Wait for user to start | Off |
| Start measurements | Single measurement |

### Routine

| | |
|---|---|
| Slab group | 1 |
| Slabs | 1 |
| Dist. factor | 20 % |
| Position | L0.0 A20.0 H0.0 mm |
| Orientation | Sagittal |
| Phase enc. dir. | A >> P |
| Phase oversampling | 0 % |
| Slice oversampling | 0.0 % |
| Slices per slab | 128 |
| FoV read | 260 mm |
| FoV phase | 100.0 % |
| Slice thickness | 1.6 mm |
| TR | 3.15 ms |
| TE | 1.37 ms |
| Averages | 1 |
| Concatenations | 1 |
| Filter | Prescan Normalize |
| Coil elements | HC1-7;NC1,2 |

### Contrast - Common

| | |
|---|---|
| TR | 3.15 ms |
| TE | 1.37 ms |
| Flip angle | 8 deg |

### Contrast - Dynamic

| | |
|---|---|
| Averages | 1 |
| Averaging mode | Short term |
| Reconstruction | Magnitude |
| Measurements | 1 |

### Resolution - Common

| | |
|---|---|
| FoV read | 260 mm |
| FoV phase | 100.0 % |
| Slice thickness | 1.6 mm |
| Base resolution | 160 |
| Phase resolution | 100 % |
| Slice resolution | 69 % |
| Phase partial Fourier | 6/8 |
| Slice partial Fourier | 6/8 |
| Trajectory | Cartesian |

### Resolution - iPAT

| | |
|---|---|
| PAT mode | GRAPPA |
| Accel. factor PE | 3 |
| Ref. lines PE | 24 |
| Accel. factor 3D | 1 |

### Resolution - iPAT

| | |
|---|---|
| Reference scan mode | Integrated |

### Resolution - Filter Image

| | |
|---|---|
| Image Filter | Off |
| Distortion Corr. | Off |
| Prescan Normalize | On |
| Unfiltered images | Off |
| Normalize | Off |
| B1 filter | Off |

### Resolution - Filter Rawdata

| | |
|---|---|
| Raw filter | Off |
| Elliptical filter | Off |

### Geometry - Common

| | |
|---|---|
| Slab group | 1 |
| Slabs | 1 |
| Dist. factor | 20 % |
| Position | L0.0 A20.0 H0.0 mm |
| Orientation | Sagittal |
| Phase enc. dir. | A >> P |
| Slice oversampling | 0.0 % |
| Slices per slab | 128 |
| FoV read | 260 mm |
| FoV phase | 100.0 % |
| Slice thickness | 1.6 mm |
| TR | 3.15 ms |
| Multi-slice mode | Sequential |
| Series | Ascending |
| Concatenations | 1 |

### Geometry - AutoAlign

| | |
|---|---|
| Slab group | 1 |
| Position | L0.0 A20.0 H0.0 mm |
| Orientation | Sagittal |
| Phase enc. dir. | A >> P |
| Initial Position | Isocenter |
| L | 0.0 mm |
| P | 0.0 mm |
| H | 0.0 mm |
| Initial Rotation | 0.00 deg |
| Initial Orientation | Transversal |

### System - Miscellaneous

| | |
|---|---|
| Positioning mode | REF |
| Table position | H |
| Table position | 0 mm |
| MSMA | S - C - T |
| Sagittal | R >> L |
| Coronal | A >> P |
| Transversal | F >> H |
| Coil Combine Mode | Adaptive Combine |
| Save uncombined | Off |
| Matrix Optimization | Off |
| Coil Select Mode | Off - All |

### System - Adjustments

| | |
|---|---|
| B0 Shim mode | Tune up |
| B1 Shim mode | TrueForm |
| Adjust with body coil | Off |

### System - Adjustments

| | |
|---|---|
| Confirm freq. adjustment | Off |
| Assume Dominant Fat | Off |
| Assume Silicone | Off |
| Adjustment Tolerance | Auto |

### System - Adjust Volume

| | |
|---|---|
| Position | Isocenter |
| Orientation | Transversal |
| Rotation | 0.00 deg |
| A >> P | 263 mm |
| R >> L | 350 mm |
| F >> H | 350 mm |
| Reset | Off |

### System - pTx Volumes

| | |
|---|---|
| B1 Shim mode | TrueForm |
| Excitation | Non-sel. |

### System - Tx/Rx

| | |
|---|---|
| Frequency 1H | 123.260801 MHz |
| Correction factor | 1 |
| Gain | Low |
| Img. Scale Cor. | 1.000 |
| Reset | Off |
| ? Ref. amplitude 1H | 0.000 V |

### Physio - PACE

| | |
|---|---|
| Resp. control | Off |
| Concatenations | 1 |

### Inline - Common

| | |
|---|---|
| Flip angle | 8 deg |
| Measurements | 1 |
| Time to center | 6.2 s |

### Inline - Inline

| | |
|---|---|
| Subtract | Off |
| Measurements | 1 |
| StdDev | Off |
| Save original images | On |

### Inline - MIP

| | |
|---|---|
| MIP-Sag | Off |
| MIP-Cor | Off |
| MIP-Tra | Off |
| MIP-Time | Off |
| Save original images | On |

### Inline - Composing

| | |
|---|---|
| Distortion Corr. | Off |

### Inline - MapIt

| | |
|---|---|
| Save original images | On |
| MapIt | None |
| Flip angle | 8 deg |
| Measurements | 1 |
| Contrasts | 1 |
| TR | 3.15 ms |
| TE | 1.37 ms |

### Sequence - Part 1

| | |
|---|---|
| Introduction | On |
| Dimension | 3D |

### Sequence - Part 1

| | |
|---|---|
| Asymmetric echo | Weak |
| Contrasts | 1 |
| Multi-slice mode | Sequential |
| Bandwidth | 540 Hz/Px |

### Sequence - Part 2

| | |
|---|---|
| RF pulse type | Fast |
| Gradient mode | Normal |
| Excitation | Non-sel. |
| RF spoiling | On |

### Sequence - Assistant

| | |
|---|---|
| Mode | Off |

| \\USER\Holmes\PCY\CMRR Holmes_Yale_McLean_Jan2022\Localizer_aligned |
|---|
| TA: 0:21 PM: REF Voxel size: 1.2×1.2×5.0 mmPAT: Off Rel. SNR: 1.00 : fl |

### Properties

| | |
|---|---|
| Prio recon | On |
| Load images to viewer | On |
| Inline movie | Off |
| Auto store images | On |
| Load images to stamp segments | On |
| Load images to graphic segments | On |
| Auto open inline display | Off |
| Auto close inline display | Off |
| Start measurement without further preparation | On |
| Wait for user to start | Off |
| Start measurements | Single measurement |

### Routine

| | |
|---|---|
| Slice group | 1 |
| Slices | 1 |
| Dist. factor | 20 % |
| Position | Isocenter |
| Orientation | Transversal |
| Phase enc. dir. | A >> P |
| Slice group | 2 |
| Slices | 7 |
| Dist. factor | 200 % |
| Position | Isocenter |
| Orientation | Sagittal |
| Phase enc. dir. | A >> P |
| Slice group | 3 |
| Slices | 1 |
| Dist. factor | 20 % |
| Position | Isocenter |
| Orientation | Coronal |
| Phase enc. dir. | R >> L |
| AutoAlign | Head > Brain |
| Phase oversampling | 0 % |
| FoV read | 300 mm |
| FoV phase | 100.0 % |
| Slice thickness | 5.0 mm |
| TR | 104.0 ms |
| TE | 3.00 ms |
| Averages | 1 |
| Concatenations | 1 |
| Filter | Prescan Normalize, Elliptical filter |
| Coil elements | HC1-7;NC1,2 |

### Contrast - Common

| | |
|---|---|
| TR | 104.0 ms |
| TE | 3.00 ms |
| MTC | Off |
| Magn. preparation | None |
| Flip angle | 15 deg |
| Fat suppr. | None |
| Water suppr. | None |

### Contrast - Dynamic

| | |
|---|---|
| Averages | 1 |
| Averaging mode | Short term |
| Reconstruction | Magnitude |
| Measurements | 1 |
| Multiple series | Off |

### Resolution - Common

| | |
|---|---|
| FoV read | 300 mm |
| FoV phase | 100.0 % |
| Slice thickness | 5.0 mm |
| Base resolution | 256 |
| Phase resolution | 75 % |
| Phase partial Fourier | Off |
| Interpolation | Off |

### Resolution - iPAT

| | |
|---|---|
| PAT mode | None |

### Resolution - Filter Image

| | |
|---|---|
| Image Filter | Off |
| Distortion Corr. | Off |
| Prescan Normalize | On |
| Unfiltered images | Off |
| Normalize | Off |
| B1 filter | Off |

### Resolution - Filter Rawdata

| | |
|---|---|
| Raw filter | Off |
| Elliptical filter | On |

### Geometry - Common

| | |
|---|---|
| Slice group | 1 |
| Slices | 1 |
| Dist. factor | 20 % |
| Position | Isocenter |
| Orientation | Transversal |
| Phase enc. dir. | A >> P |
| Slice group | 2 |
| Slices | 7 |
| Dist. factor | 200 % |
| Position | Isocenter |
| Orientation | Sagittal |
| Phase enc. dir. | A >> P |
| Slice group | 3 |
| Slices | 1 |
| Dist. factor | 20 % |
| Position | Isocenter |
| Orientation | Coronal |
| Phase enc. dir. | R >> L |
| FoV read | 300 mm |
| FoV phase | 100.0 % |
| Slice thickness | 5.0 mm |
| TR | 104.0 ms |
| Multi-slice mode | Interleaved |
| Series | Interleaved |
| Concatenations | 1 |

### Geometry - AutoAlign

| | |
|---|---|
| Slice group | 1 |
| Position | Isocenter |
| Orientation | Transversal |
| Phase enc. dir. | A >> P |
| Slice group | 2 |
| Position | Isocenter |
| Orientation | Sagittal |
| Phase enc. dir. | A >> P |
| Slice group | 3 |

### Geometry - AutoAlign

| | |
|---|---|
| Position | Isocenter |
| Orientation | Coronal |
| Phase enc. dir. | R >> L |
| AutoAlign | Head > Brain |
| Initial Position | Isocenter |
| L | 0.0 mm |
| P | 0.0 mm |
| H | 0.0 mm |
| Initial Rotation | 0.00 deg |
| Initial Orientation | Transversal |

### Geometry - Saturation

| | |
|---|---|
| Saturation mode | Standard |
| Fat suppr. | None |
| Water suppr. | None |
| Special sat. | None |

### System - Miscellaneous

| | |
|---|---|
| Positioning mode | REF |
| Table position | H |
| Table position | 0 mm |
| MSMA | S - C - T |
| Sagittal | R >> L |
| Coronal | A >> P |
| Transversal | F >> H |
| Coil Combine Mode | Sum of Squares |
| Save uncombined | Off |
| Matrix Optimization | Off |
| AutoAlign | Head > Brain |
| Coil Select Mode | Off - All |

### System - Adjustments

| | |
|---|---|
| B0 Shim mode | Tune up |
| B1 Shim mode | TrueForm |
| Adjust with body coil | Off |
| Confirm freq. adjustment | Off |
| Assume Dominant Fat | Off |
| Assume Silicone | Off |
| Adjustment Tolerance | Auto |

### System - Adjust Volume

| | |
|---|---|
| Position | Isocenter |
| Orientation | Transversal |
| Rotation | 0.00 deg |
| A >> P | 263 mm |
| R >> L | 350 mm |
| F >> H | 350 mm |
| Reset | Off |

### System - pTx Volumes

| | |
|---|---|
| B1 Shim mode | TrueForm |
| Excitation | Slice-sel. |

### System - Tx/Rx

| | |
|---|---|
| Frequency 1H | 123.260801 MHz |
| Correction factor | 1 |
| Gain | High |
| Img. Scale Cor. | 1.000 |
| Reset | Off |
| ? Ref. amplitude 1H | 0.000 V |

### Physio - Signal1

| | |
|---|---|
| 1st Signal/Mode | None |

### Physio - Signal1

| | |
|---|---|
| TR | 104.0 ms |
| Concatenations | 1 |
| Segments | 1 |

### Physio - Cardiac

| | |
|---|---|
| Magn. preparation | None |
| Fat suppr. | None |
| Dark blood | Off |
| FoV read | 300 mm |
| FoV phase | 100.0 % |
| Phase resolution | 75 % |

### Physio - PACE

| | |
|---|---|
| Resp. control | Off |
| Concatenations | 1 |

### Inline - Common

| | |
|---|---|
| Subtract | Off |
| Measurements | 1 |
| StdDev | Off |
| Liver registration | Off |
| Save original images | On |

### Inline - MIP

| | |
|---|---|
| MIP-Sag | Off |
| MIP-Cor | Off |
| MIP-Tra | Off |
| MIP-Time | Off |
| Save original images | On |

### Inline - Soft Tissue

| | |
|---|---|
| Wash - In | Off |
| Wash - Out | Off |
| TTP | Off |
| PEI | Off |
| MIP - time | Off |
| Measurements | 1 |

### Inline - Composing

| | |
|---|---|
| Distortion Corr. | Off |

### Inline - MapIt

| | |
|---|---|
| Save original images | On |
| MapIt | None |
| Flip angle | 15 deg |
| Measurements | 1 |
| Contrasts | 1 |
| TR | 104.0 ms |
| TE | 3.00 ms |

### Sequence - Part 1

| | |
|---|---|
| Introduction | On |
| Dimension | 2D |
| Phase stabilisation | Off |
| Asymmetric echo | Allowed |
| Contrasts | 1 |
| Flow comp. | No |
| Multi-slice mode | Interleaved |
| Bandwidth | 260 Hz/Px |

### Sequence - Part 2

| | |
|---|---|
| Segments | 1 |
| Acoustic noise reduction | None |

### Sequence - Part 2

| | |
|---|---|
| RF pulse type | Normal |
| Gradient mode | Fast |
| Excitation | Slice-sel. |
| RF spoiling | On |

### Sequence - Assistant

| | |
|---|---|
| Mode | Off |
| Allowed delay | 0 s |

**\\USER\Holmes\PCY\CMRR Holmes_Yale_McLean_Jan2022\T1_vNav_setter**

TA: 0.3 s PM: FIX Voxel size: 8.0×8.0×8.0 mmRel. SNR: 1.00 : ABCD

### Properties

| | |
|---|---|
| Prio recon | Off |
| Load images to viewer | On |
| Inline movie | Off |
| Auto store images | On |
| Load images to stamp segments | Off |
| Load images to graphic segments | Off |
| Auto open inline display | Off |
| Auto close inline display | Off |
| Start measurement without further preparation | On |
| Wait for user to start | On |
| Start measurements | Single measurement |

### Routine

| | |
|---|---|
| Slab group | 1 |
| Slabs | 1 |
| Dist. factor | 50 % |
| Position | L0.0 P0.0 H50.0 mm |
| Orientation | Sagittal |
| Phase enc. dir. | A >> P |
| AutoAlign | Head > Brain |
| Phase oversampling | 0 % |
| Slice oversampling | 0.0 % |
| Slices per slab | 32 |
| FoV read | 256 mm |
| FoV phase | 100.0 % |
| Slice thickness | 8.00 mm |
| TR | 11.0 ms |
| TE | 4.8 ms |
| Averages | 1 |
| Concatenations | 1 |
| Filter | None |
| Coil elements | BC |

### Contrast - Common

| | |
|---|---|
| TR | 11.0 ms |
| TE | 4.8 ms |
| MTC | Off |
| Flip angle | 2 deg |
| Fat suppr. | None |

### Contrast - Dynamic

| | |
|---|---|
| Averages | 1 |
| Averaging mode | Long term |
| Reconstruction | Magnitude |
| Measurements | 1 |
| Multiple series | Each measurement |

### Resolution - Common

| | |
|---|---|
| FoV read | 256 mm |
| FoV phase | 100.0 % |
| Slice thickness | 8.00 mm |
| Base resolution | 32 |
| Phase resolution | 100 % |
| Slice resolution | 100 % |
| Phase partial Fourier | Off |
| Slice partial Fourier | 6/8 |
| Interpolation | Off |

### Resolution - Filter Image

| | |
|---|---|
| Distortion Corr. | Off |
| Prescan Normalize | Off |

### Resolution - Filter Rawdata

| | |
|---|---|
| Raw filter | Off |
| Elliptical filter | Off |
| Hamming | Off |

### Geometry - Common

| | |
|---|---|
| Slab group | 1 |
| Slabs | 1 |
| Dist. factor | 50 % |
| Position | L0.0 P0.0 H50.0 mm |
| Orientation | Sagittal |
| Phase enc. dir. | A >> P |
| Slice oversampling | 0.0 % |
| Slices per slab | 32 |
| FoV read | 256 mm |
| FoV phase | 100.0 % |
| Slice thickness | 8.00 mm |
| TR | 11.0 ms |
| Multi-slice mode | Interleaved |
| Series | Interleaved |
| Concatenations | 1 |

### Geometry - AutoAlign

| | |
|---|---|
| Slab group | 1 |
| Position | L0.0 P0.0 H50.0 mm |
| Orientation | Sagittal |
| Phase enc. dir. | A >> P |
| AutoAlign | Head > Brain |
| Initial Position | L0.0 P0.0 H50.0 |
| R | 0.0 mm |
| A | 0.0 mm |
| H | 50.0 mm |
| Initial Rotation | 0.00 deg |
| Initial Orientation | Sagittal |

### Geometry - Saturation

| | |
|---|---|
| Fat suppr. | None |
| Special sat. | None |

### System - Miscellaneous

| | |
|---|---|
| Positioning mode | FIX |
| Table position | H |
| Table position | 0 mm |
| MSMA | S - C - T |
| Sagittal | R >> L |
| Coronal | A >> P |
| Transversal | F >> H |
| Coil Combine Mode | Sum of Squares |
| Save uncombined | Off |
| Matrix Optimization | Off |
| AutoAlign | Head > Brain |
| Coil Select Mode | Off - All |

### System - Adjustments

| | |
|---|---|
| B0 Shim mode | Tune up |
| B1 Shim mode | TrueForm |
| Adjust with body coil | Off |

### System - Adjustments

| | |
|---|---|
| Confirm freq. adjustment | Off |
| Assume Dominant Fat | Off |
| Assume Silicone | Off |
| Adjustment Tolerance | Auto |

### System - Adjust Volume

| | |
|---|---|
| Position | Isocenter |
| Orientation | Transversal |
| Rotation | 0.00 deg |
| A >> P | 263 mm |
| R >> L | 350 mm |
| F >> H | 350 mm |
| Reset | Off |

### System - pTx Volumes

| | |
|---|---|
| B1 Shim mode | TrueForm |
| Excitation | Non-sel. |

### System - Tx/Rx

| | |
|---|---|
| Frequency 1H | 123.260801 MHz |
| Correction factor | 1 |
| Gain | Low |
| Img. Scale Cor. | 50.000 |
| Reset | Off |
| ? Ref. amplitude 1H | 0.000 V |

### Physio - Signal1

| | |
|---|---|
| 1st Signal/Mode | None |
| TR | 11.0 ms |
| Concatenations | 1 |

### Sequence - Part 1

| | |
|---|---|
| Introduction | Off |
| Dimension | 3D |
| Contrasts | 1 |
| Multi-slice mode | Interleaved |
| Free echo spacing | Off |
| Echo spacing | 0.27 ms |
| Bandwidth | 4882 Hz/Px |

### Sequence - Part 2

| | |
|---|---|
| EPI factor | 32 |
| RF pulse type | Normal |
| Gradient mode | Fast |
| Excitation | Non-sel. |
| RF spoiling | On |

### Sequence - Special

| | |
|---|---|
| Protocol filename | MPRAGE |

| \\USER\Holmes\PCY\CMRR Holmes_Yale_McLean_Jan2022\T1_MEMPRAGE_GSP_vNavTrk |
|---|
| TA: 2:23 PM: FIX Voxel size: 1.2×1.2×1.2 mmPAT: 4 Rel. SNR: 1.00 : tfl |

### Properties

| | |
|---|---|
| Prio recon | Off |
| Load images to viewer | On |
| Inline movie | Off |
| Auto store images | On |
| Load images to stamp segments | Off |
| Load images to graphic segments | On |
| Auto open inline display | Off |
| Auto close inline display | Off |
| Start measurement without further preparation | On |
| Wait for user to start | Off |
| Start measurements | Single measurement |

### Routine

| | |
|---|---|
| Slab group | 1 |
| Slabs | 1 |
| Dist. factor | 50 % |
| Position | Isocenter |
| Orientation | Sagittal |
| Phase enc. dir. | A >> P |
| AutoAlign | Head > Brain |
| Phase oversampling | 0 % |
| Slice oversampling | 0.0 % |
| Slices per slab | 144 |
| FoV read | 230 mm |
| FoV phase | 100.0 % |
| Slice thickness | 1.20 mm |
| TR | 2200.0 ms |
| TE 1 | 1.57 ms |
| TE 2 | 3.39 ms |
| TE 3 | 5.21 ms |
| TE 4 | 7.03 ms |
| Averages | 1 |
| Concatenations | 1 |
| Filter | Prescan Normalize |
| Coil elements | HC1-7;NC1,2 |

### Contrast - Common

| | |
|---|---|
| TR | 2200.0 ms |
| TE 1 | 1.57 ms |
| TE 2 | 3.39 ms |
| TE 3 | 5.21 ms |
| TE 4 | 7.03 ms |
| Magn. preparation | Non-sel. IR |
| TI | 1100 ms |
| Flip angle | 7.0 deg |
| Fat suppr. | None |
| Water suppr. | None |

### Contrast - Dynamic

| | |
|---|---|
| Averages | 1 |
| Averaging mode | Long term |
| Reconstruction | Magnitude |
| Measurements | 1 |
| Multiple series | Each measurement |

### Resolution - Common

| | |
|---|---|
| FoV read | 230 mm |
| FoV phase | 100.0 % |
| Slice thickness | 1.20 mm |

### Resolution - Common

| | |
|---|---|
| Base resolution | 192 |
| Phase resolution | 100 % |
| Slice resolution | 100 % |
| Phase partial Fourier | 6/8 |
| Slice partial Fourier | Off |
| Interpolation | Off |

### Resolution - iPAT

| | |
|---|---|
| PAT mode | GRAPPA |
| Accel. factor PE | 4 |
| Ref. lines PE | 32 |
| Accel. factor 3D | 1 |
| Reference scan mode | Integrated |

### Resolution - Filter Image

| | |
|---|---|
| Image Filter | Off |
| Distortion Corr. | Off |
| Prescan Normalize | On |
| Unfiltered images | On |
| Normalize | Off |
| B1 filter | Off |

### Resolution - Filter Rawdata

| | |
|---|---|
| Raw filter | Off |
| Elliptical filter | Off |

### Geometry - Common

| | |
|---|---|
| Slab group | 1 |
| Slabs | 1 |
| Dist. factor | 50 % |
| Position | Isocenter |
| Orientation | Sagittal |
| Phase enc. dir. | A >> P |
| Slice oversampling | 0.0 % |
| Slices per slab | 144 |
| FoV read | 230 mm |
| FoV phase | 100.0 % |
| Slice thickness | 1.20 mm |
| TR | 2200.0 ms |
| Multi-slice mode | Single shot |
| Series | Interleaved |
| Concatenations | 1 |

### Geometry - AutoAlign

| | |
|---|---|
| Slab group | 1 |
| Position | Isocenter |
| Orientation | Sagittal |
| Phase enc. dir. | A >> P |
| AutoAlign | Head > Brain |
| Initial Position | Isocenter |
| L | 0.0 mm |
| P | 0.0 mm |
| H | 0.0 mm |
| Initial Rotation | 16.00 deg |
| Initial Orientation | Sagittal |

### System - Miscellaneous

| | |
|---|---|
| Positioning mode | FIX |
| Table position | H |
| Table position | 0 mm |

### System - Miscellaneous

| | |
|---|---|
| MSMA | S - C - T |
| Sagittal | R >> L |
| Coronal | A >> P |
| Transversal | F >> H |
| Coil Combine Mode | Adaptive Combine |
| Save uncombined | Off |
| Matrix Optimization | Off |
| AutoAlign | Head > Brain |
| Coil Select Mode | Off - All |

### System - Adjustments

| | |
|---|---|
| B0 Shim mode | Standard |
| B1 Shim mode | TrueForm |
| Adjust with body coil | Off |
| Confirm freq. adjustment | Off |
| Assume Dominant Fat | Off |
| Assume Silicone | Off |
| Adjustment Tolerance | Auto |

### System - Adjust Volume

| | |
|---|---|
| ! Position | L0.0 P0.0 H6.0 mm |
| ! Orientation | T > C-16.0 |
| ! Rotation | 0.00 deg |
| ! A >> P | 208 mm |
| ! R >> L | 208 mm |
| ! F >> H | 144 mm |
| Reset | Off |

### System - pTx Volumes

| | |
|---|---|
| B1 Shim mode | TrueForm |
| Excitation | Non-sel. |

### System - Tx/Rx

| | |
|---|---|
| Frequency 1H | 123.260801 MHz |
| Correction factor | 1 |
| Gain | Low |
| Img. Scale Cor. | 1.000 |
| Reset | Off |
| ? Ref. amplitude 1H | 0.000 V |

### Physio - Signal1

| | |
|---|---|
| 1st Signal/Mode | None |
| TR | 2200.0 ms |
| Concatenations | 1 |

### Physio - Cardiac

| | |
|---|---|
| Magn. preparation | Non-sel. IR |
| TI | 1100 ms |
| Fat suppr. | None |
| Dark blood | Off |
| FoV read | 230 mm |
| FoV phase | 100.0 % |
| Phase resolution | 100 % |

### Inline - Common

| | |
|---|---|
| Subtract | Off |
| Measurements | 1 |
| StdDev | Off |
| Save original images | On |

### Inline - MIP

| | |
|---|---|
| MIP-Sag | Off |
| MIP-Cor | Off |
| MIP-Tra | Off |
| MIP-Time | Off |
| Save original images | On |

### Inline - Composing

| | |
|---|---|
| Distortion Corr. | Off |

### Inline - MapIt

| | |
|---|---|
| Save original images | On |
| MapIt | None |
| Flip angle | 7.0 deg |
| Measurements | 1 |
| Contrasts | 4 |
| TR | 2200.0 ms |
| TE 1 | 1.57 ms |
| TE 2 | 3.39 ms |
| TE 3 | 5.21 ms |
| TE 4 | 7.03 ms |

### Sequence - Part 1

| | |
|---|---|
| Introduction | Off |
| Dimension | 3D |
| Elliptical scanning | Off |
| Reordering | Linear |
| Asymmetric echo | Off |
| Contrasts | 4 |
| Flow comp. 1 | No |
| Multi-slice mode | Single shot |
| Echo spacing | 9.4 ms |
| Bandwidth 1 | 650 Hz/Px |
| Bandwidth 2 | 650 Hz/Px |
| Bandwidth 3 | 650 Hz/Px |
| Bandwidth 4 | 650 Hz/Px |

### Sequence - Part 2

| | |
|---|---|
| RF pulse type | Fast |
| Gradient mode | Fast |
| Excitation | Non-sel. |
| RF spoiling | On |
| Turbo factor | 144 |

### Sequence - Special

| | |
|---|---|
| Readout polarity | Positive |
| Nav. location | Before |
| Apply moco to | nav only |
| Remeasure | 0 TRs |
| Reacq. threshold | 0.50 |
| Feedback delay | 80 ms |
| Moco ref. image | Use Temp Ref |
| K-space streaming | None |
| ABCD navigator | On |
| Add. grad time | 0.0 ms |
| Apply freq to | parent and nav |
| Averaging | RMS |

### Sequence - Assistant

| | |
|---|---|
| Mode | Off |

**\\USER\Holmes\PCY\CMRR Holmes_Yale_McLean_Jan2022\T2_vNav_setter**

TA: 0.3 s PM: FIX Voxel size: 8.0×8.0×8.0 mmRel. SNR: 1.00 : ABCD

### Properties

| | |
|---|---|
| Prio recon | Off |
| Load images to viewer | On |
| Inline movie | Off |
| Auto store images | On |
| Load images to stamp segments | Off |
| Load images to graphic segments | Off |
| Auto open inline display | Off |
| Auto close inline display | Off |
| Start measurement without further preparation | On |
| Wait for user to start | Off |
| Start measurements | Single measurement |

### Routine

| | |
|---|---|
| Slab group | 1 |
| Slabs | 1 |
| Dist. factor | 50 % |
| Position | L0.0 P0.0 H50.0 mm |
| Orientation | Sagittal |
| Phase enc. dir. | A >> P |
| AutoAlign | Head > Brain |
| Phase oversampling | 0 % |
| Slice oversampling | 0.0 % |
| Slices per slab | 32 |
| FoV read | 256 mm |
| FoV phase | 100.0 % |
| Slice thickness | 8.00 mm |
| TR | 13.0 ms |
| TE | 6.1 ms |
| Averages | 1 |
| Concatenations | 1 |
| Filter | None |
| Coil elements | BC |

### Contrast - Common

| | |
|---|---|
| TR | 13.0 ms |
| TE | 6.1 ms |
| MTC | Off |
| Flip angle | 2 deg |
| Fat suppr. | Water excit. normal |

### Contrast - Dynamic

| | |
|---|---|
| Averages | 1 |
| Averaging mode | Long term |
| Reconstruction | Magnitude |
| Measurements | 1 |
| Multiple series | Each measurement |

### Resolution - Common

| | |
|---|---|
| FoV read | 256 mm |
| FoV phase | 100.0 % |
| Slice thickness | 8.00 mm |
| Base resolution | 32 |
| Phase resolution | 100 % |
| Slice resolution | 100 % |
| Phase partial Fourier | Off |
| Slice partial Fourier | 6/8 |
| Interpolation | Off |

### Resolution - Filter Image

| | |
|---|---|
| Distortion Corr. | Off |
| Prescan Normalize | Off |

### Resolution - Filter Rawdata

| | |
|---|---|
| Raw filter | Off |
| Elliptical filter | Off |
| Hamming | Off |

### Geometry - Common

| | |
|---|---|
| Slab group | 1 |
| Slabs | 1 |
| Dist. factor | 50 % |
| Position | L0.0 P0.0 H50.0 mm |
| Orientation | Sagittal |
| Phase enc. dir. | A >> P |
| Slice oversampling | 0.0 % |
| Slices per slab | 32 |
| FoV read | 256 mm |
| FoV phase | 100.0 % |
| Slice thickness | 8.00 mm |
| TR | 13.0 ms |
| Multi-slice mode | Interleaved |
| Series | Interleaved |
| Concatenations | 1 |

### Geometry - AutoAlign

| | |
|---|---|
| Slab group | 1 |
| Position | L0.0 P0.0 H50.0 mm |
| Orientation | Sagittal |
| Phase enc. dir. | A >> P |
| AutoAlign | Head > Brain |
| Initial Position | L0.0 P0.0 H50.0 |
| R | 0.0 mm |
| A | 0.0 mm |
| H | 50.0 mm |
| Initial Rotation | 0.00 deg |
| Initial Orientation | Sagittal |

### Geometry - Saturation

| | |
|---|---|
| Fat suppr. | Water excit. normal |
| Special sat. | None |

### System - Miscellaneous

| | |
|---|---|
| Positioning mode | FIX |
| Table position | H |
| Table position | 0 mm |
| MSMA | S - C - T |
| Sagittal | R >> L |
| Coronal | A >> P |
| Transversal | F >> H |
| Coil Combine Mode | Sum of Squares |
| Save uncombined | Off |
| Matrix Optimization | Off |
| AutoAlign | Head > Brain |
| Coil Select Mode | Off - All |

### System - Adjustments

| | |
|---|---|
| B0 Shim mode | Tune up |
| B1 Shim mode | TrueForm |
| Adjust with body coil | Off |

### System - Adjustments

| | |
|---|---|
| Confirm freq. adjustment | Off |
| Assume Dominant Fat | Off |
| Assume Silicone | Off |
| Adjustment Tolerance | Auto |

### System - Adjust Volume

| | |
|---|---|
| Position | Isocenter |
| Orientation | Transversal |
| Rotation | 0.00 deg |
| A >> P | 263 mm |
| R >> L | 350 mm |
| F >> H | 350 mm |
| Reset | Off |

### System - pTx Volumes

| | |
|---|---|
| B1 Shim mode | TrueForm |
| Excitation | Non-sel. |

### System - Tx/Rx

| | |
|---|---|
| Frequency 1H | 123.260801 MHz |
| Correction factor | 1 |
| Gain | Low |
| Img. Scale Cor. | 5.000 |
| Reset | Off |
| ? Ref. amplitude 1H | 0.000 V |

### Physio - Signal1

| | |
|---|---|
| 1st Signal/Mode | None |
| TR | 13.0 ms |
| Concatenations | 1 |

### Sequence - Part 1

| | |
|---|---|
| Introduction | Off |
| Dimension | 3D |
| Contrasts | 1 |
| Multi-slice mode | Interleaved |
| Free echo spacing | Off |
| Echo spacing | 0.27 ms |
| Bandwidth | 4882 Hz/Px |

### Sequence - Part 2

| | |
|---|---|
| EPI factor | 32 |
| RF pulse type | Normal |
| Gradient mode | Fast |
| Excitation | Non-sel. |
| RF spoiling | On |

### Sequence - Special

| | |
|---|---|
| Protocol filename | T2-SPACE |

**\\USER\Holmes\PCY\CMRR Holmes_Yale_McLean_Jan2022\T2_SPACE_GSP_vNavTrk**

TA: 2:37 PM: FIX Voxel size: 1.2×1.2×1.2 mmPAT: 2 Rel. SNR: 1.00 : spc

### Properties

| | |
|---|---|
| Prio recon | Off |
| Load images to viewer | On |
| Inline movie | Off |
| Auto store images | On |
| Load images to stamp segments | Off |
| Load images to graphic segments | Off |
| Auto open inline display | Off |
| Auto close inline display | Off |
| Start measurement without further preparation | On |
| Wait for user to start | Off |
| Start measurements | Single measurement |

### Routine

| | |
|---|---|
| Slab group | 1 |
| Slabs | 1 |
| Position | Isocenter |
| Orientation | Sagittal |
| Phase enc. dir. | A >> P |
| AutoAlign | Head > Brain |
| Phase oversampling | 0 % |
| Slice oversampling | 0.0 % |
| Slices per slab | 144 |
| FoV read | 230 mm |
| FoV phase | 100.0 % |
| Slice thickness | 1.20 mm |
| TR | 2800 ms |
| TE | 326 ms |
| Averages | 1.0 |
| Concatenations | 1 |
| Filter | Raw filter, Prescan Normalize |
| Coil elements | HC1-7;NC1,2 |

### Contrast - Common

| | |
|---|---|
| TR | 2800 ms |
| TE | 326 ms |
| MTC | Off |
| Magn. preparation | None |
| Fat suppr. | None |
| Blood suppr. | Off |
| Restore magn. | Off |

### Contrast - Dynamic

| | |
|---|---|
| Averages | 1.0 |
| Reconstruction | Magnitude |
| Measurements | 1 |
| Multiple series | Each measurement |

### Resolution - Common

| | |
|---|---|
| FoV read | 230 mm |
| FoV phase | 100.0 % |
| Slice thickness | 1.20 mm |
| Base resolution | 192 |
| Phase resolution | 100 % |
| Slice resolution | 100 % |
| Phase partial Fourier | Allowed |
| Slice partial Fourier | 6/8 |
| Interpolation | Off |

### Resolution - iPAT

| | |
|---|---|
| PAT mode | GRAPPA |
| Accel. factor PE | 2 |
| Ref. lines PE | 24 |
| Accel. factor 3D | 1 |
| Reference scan mode | Integrated |

### Resolution - Filter Image

| | |
|---|---|
| Image Filter | Off |
| Distortion Corr. | Off |
| Prescan Normalize | On |
| Unfiltered images | On |
| Normalize | Off |
| B1 filter | Off |

### Resolution - Filter Rawdata

| | |
|---|---|
| Raw filter | On |
| Elliptical filter | Off |

### Geometry - Common

| | |
|---|---|
| Slab group | 1 |
| Slabs | 1 |
| Position | Isocenter |
| Orientation | Sagittal |
| Phase enc. dir. | A >> P |
| Slice oversampling | 0.0 % |
| Slices per slab | 144 |
| FoV read | 230 mm |
| FoV phase | 100.0 % |
| Slice thickness | 1.20 mm |
| TR | 2800 ms |
| Concatenations | 1 |

### Geometry - AutoAlign

| | |
|---|---|
| Slab group | 1 |
| Position | Isocenter |
| Orientation | Sagittal |
| Phase enc. dir. | A >> P |
| AutoAlign | Head > Brain |
| Initial Position | Isocenter |
| L | 0.0 mm |
| P | 0.0 mm |
| H | 0.0 mm |
| Initial Rotation | 16.00 deg |
| Initial Orientation | Sagittal |

### Geometry - Saturation

| | |
|---|---|
| Fat suppr. | None |
| Restore magn. | Off |
| Special sat. | None |

### System - Miscellaneous

| | |
|---|---|
| Positioning mode | FIX |
| Table position | H |
| Table position | 0 mm |
| MSMA | S - C - T |
| Sagittal | R >> L |
| Coronal | A >> P |
| Transversal | F >> H |
| Coil Combine Mode | Adaptive Combine |
| Save uncombined | Off |

### System - Miscellaneous

| | |
|---|---|
| Matrix Optimization | Off |
| AutoAlign | Head > Brain |
| Coil Select Mode | Off - All |

### System - Adjustments

| | |
|---|---|
| B0 Shim mode | Standard |
| B1 Shim mode | TrueForm |
| Adjust with body coil | Off |
| Confirm freq. adjustment | Off |
| Assume Dominant Fat | Off |
| Assume Silicone | Off |
| Adjustment Tolerance | Auto |

### System - Adjust Volume

| | |
|---|---|
| ! Position | L0.0 P0.0 H6.0 mm |
| ! Orientation | T > C-16.0 |
| ! Rotation | 0.00 deg |
| ! A >> P | 208 mm |
| ! R >> L | 208 mm |
| ! F >> H | 144 mm |
| Reset | Off |

### System - pTx Volumes

| | |
|---|---|
| B1 Shim mode | TrueForm |
| Excitation | Non-sel. |

### System - Tx/Rx

| | |
|---|---|
| Frequency 1H | 123.260801 MHz |
| Correction factor | 1 |
| Gain | High |
| Img. Scale Cor. | 1.000 |
| Reset | Off |
| ? Ref. amplitude 1H | 0.000 V |

### Physio - Signal1

| | |
|---|---|
| 1st Signal/Mode | None |
| Trigger delay | 0 ms |
| TR | 2800 ms |
| Concatenations | 1 |

### Physio - Cardiac

| | |
|---|---|
| Magn. preparation | None |
| Fat suppr. | None |
| Dark blood | Off |
| FoV read | 230 mm |
| FoV phase | 100.0 % |
| Phase resolution | 100 % |

### Inline - Common

| | |
|---|---|
| Subtract | Off |
| Measurements | 1 |
| StdDev | Off |
| Save original images | On |

### Inline - MIP

| | |
|---|---|
| MIP-Sag | Off |
| MIP-Cor | Off |
| MIP-Tra | Off |
| MIP-Time | Off |
| Save original images | On |

### Inline - Composing

| | |
|---|---|
| Distortion Corr. | Off |

### Sequence - Part 1

| | |
|---|---|
| Introduction | Off |
| Dimension | 3D |
| Elliptical scanning | Off |
| Reordering | Linear |
| Flow comp. | No |
| Echo spacing | 3.5 ms |
| Adiabatic-mode | Off |
| Bandwidth | 651 Hz/Px |

### Sequence - Part 2

| | |
|---|---|
| Echo train duration | 693 ms |
| RF pulse type | Normal |
| Gradient mode | Fast |
| Excitation | Non-sel. |
| Flip angle mode | T2 var |
| Turbo factor | 212 |

### Sequence - Special

| | |
|---|---|
| Include nav. | On |
| Apply moco to | nav only |
| Remeasure | 0 TRs |
| Reacq. threshold | 0.50 |
| Feedback delay | 80 ms |
| Moco ref. image | Use Temp Ref |
| K-space streaming | None |
| ABCD navigator | On |
| Apply freq to | parent and nav |

### Sequence - Assistant

| | |
|---|---|
| Allowed delay | 0 s |

**\\USER\Holmes\PCY\CMRR Holmes_Yale_McLean_Jan2022\SpinEchoFieldMap_AP**

TA: 8.0 s PM: REF Voxel size: 2.0×2.0×2.0 mmPAT: Off Rel. SNR: 1.00 : epse

### Properties

| | |
|---|---|
| Prio recon | Off |
| Load images to viewer | On |
| Inline movie | Off |
| Auto store images | On |
| Load images to stamp segments | Off |
| Load images to graphic segments | Off |
| Auto open inline display | Off |
| Auto close inline display | Off |
| Start measurement without further preparation | On |
| Wait for user to start | On |
| Start measurements | Single measurement |

### Routine

| | |
|---|---|
| Slice group | 1 |
| Slices | 72 |
| Dist. factor | 0 % |
| Position | L0.0 P0.0 H6.0 mm |
| Orientation | T > C-16.0 |
| Phase enc. dir. | A >> P |
| AutoAlign | Head > Brain |
| Phase oversampling | 0 % |
| FoV read | 208 mm |
| FoV phase | 100.0 % |
| Slice thickness | 2.00 mm |
| TR | 8000 ms |
| TE | 66.00 ms |
| Multi-band accel. factor | 1 |
| Filter | None |
| Coil elements | HC1-7;NC1,2 |

### Contrast - Common

| | |
|---|---|
| TR | 8000 ms |
| TE | 66.00 ms |
| MTC | Off |
| Magn. preparation | None |
| Flip angle | 90 deg |
| Refocus flip angle | 180 deg |
| Fat suppr. | Fat sat. |
| Grad. rev. fat suppr. | Disabled |

### Contrast - Dynamic

| | |
|---|---|
| Averaging mode | Long term |
| Reconstruction | Magnitude |
| Measurements | 1 |
| Delay in TR | 0 ms |
| Multiple series | Off |

### Resolution - Common

| | |
|---|---|
| FoV read | 208 mm |
| FoV phase | 100.0 % |
| Slice thickness | 2.00 mm |
| Base resolution | 104 |
| Phase resolution | 100 % |
| Phase partial Fourier | Off |
| Interpolation | Off |

### Resolution - iPAT

| | |
|---|---|
| PAT mode | None |

### Resolution - Filter Image

| | |
|---|---|
| Distortion Corr. | Off |
| Prescan Normalize | Off |

### Resolution - Filter Rawdata

| | |
|---|---|
| Raw filter | Off |
| Elliptical filter | Off |
| Hamming | Off |

### Geometry - Common

| | |
|---|---|
| Slice group | 1 |
| Slices | 72 |
| Dist. factor | 0 % |
| Position | L0.0 P0.0 H6.0 mm |
| Orientation | T > C-16.0 |
| Phase enc. dir. | A >> P |
| FoV read | 208 mm |
| FoV phase | 100.0 % |
| Slice thickness | 2.00 mm |
| TR | 8000 ms |
| Multi-slice mode | Interleaved |
| Series | Interleaved |
| Multi-band accel. factor | 1 |

### Geometry - AutoAlign

| | |
|---|---|
| Slice group | 1 |
| Position | L0.0 P0.0 H6.0 mm |
| Orientation | T > C-16.0 |
| Phase enc. dir. | A >> P |
| AutoAlign | Head > Brain |
| Initial Position | L0.0 P0.0 H6.0 |
| L | 0.0 mm |
| P | 0.0 mm |
| H | 6.0 mm |
| Initial Rotation | 0.00 deg |
| Initial Orientation | T > C |
| T > C | -16.0 |
| > S | 0.0 |

### Geometry - Saturation

| | |
|---|---|
| Fat suppr. | Fat sat. |
| Grad. rev. fat suppr. | Disabled |
| Special sat. | None |

### System - Miscellaneous

| | |
|---|---|
| Positioning mode | REF |
| Table position | H |
| Table position | 0 mm |
| MSMA | S - C - T |
| Sagittal | R >> L |
| Coronal | A >> P |
| Transversal | F >> H |
| Coil Combine Mode | Sum of Squares |
| Matrix Optimization | Off |
| AutoAlign | Head > Brain |
| Coil Select Mode | Off - All |

### System - Adjustments

| | |
|---|---|
| B0 Shim mode | Standard |
| B1 Shim mode | TrueForm |
| Adjust with body coil | Off |

### System - Adjustments

| | |
|---|---|
| Confirm freq. adjustment | Off |
| Assume Dominant Fat | Off |
| Assume Silicone | Off |
| Adjustment Tolerance | Auto |

### System - Adjust Volume

| | |
|---|---|
| Position | L0.0 P0.0 H6.0 mm |
| Orientation | T > C-16.0 |
| Rotation | 0.00 deg |
| A >> P | 208 mm |
| R >> L | 208 mm |
| F >> H | 144 mm |
| Reset | Off |

### System - pTx Volumes

| | |
|---|---|
| B1 Shim mode | TrueForm |
| Excitation | Standard |

### System - Tx/Rx

| | |
|---|---|
| Frequency 1H | 123.260801 MHz |
| Correction factor | 1 |
| Gain | High |
| Img. Scale Cor. | 1.000 |
| Reset | Off |
| ? Ref. amplitude 1H | 0.000 V |

### Physio - Signal1

| | |
|---|---|
| 1st Signal/Mode | None |
| TR | 8000 ms |
| Multi-band accel. factor | 1 |

### BOLD

| | |
|---|---|
| GLM Statistics | Off |
| Dynamic t-maps | Off |
| Ignore meas. at start | 0 |
| Ignore after transition | 0 |
| Model transition states | Off |
| Temp. highpass filter | Off |
| Threshold | 4.00 |
| Paradigm size | 3 |
| Meas[1] | Baseline |
| Meas[2] | Baseline |
| Meas[3] | Active |
| Motion correction | Off |
| Spatial filter | Off |
| Measurements | 1 |
| Delay in TR | 0 ms |
| Multiple series | Off |

### Sequence - Part 1

| | |
|---|---|
| Introduction | Off |
| Contrasts | 1 |
| Multi-slice mode | Interleaved |
| Free echo spacing | On |
| Echo spacing | 0.58 ms |
| Bandwidth | 2290 Hz/Px |

### Sequence - Part 2

| | |
|---|---|
| EPI factor | 104 |
| RF pulse type | Normal |
| Gradient mode | Performance |
| Excitation | Standard |

### Sequence - Special

| | |
|---|---|
| SENSE1 coil combine | Off |
| Invert RO/PE polarity | Off |
| Disable freq. update | Off |
| Force equal slice timing | Off |
| FFT scale factor | 1.00 |
| Physio recording | Off |
| Triggering scheme | Standard |

**\\USER\Holmes\PCY\CMRR Holmes_Yale_McLean_Jan2022\SpinEchoFieldMap_PA**

TA: 8.0 s PM: REF Voxel size: 2.0×2.0×2.0 mmPAT: Off Rel. SNR: 1.00 : epse

### Properties

| | |
|---|---|
| Prio recon | Off |
| Load images to viewer | On |
| Inline movie | Off |
| Auto store images | On |
| Load images to stamp segments | Off |
| Load images to graphic segments | Off |
| Auto open inline display | Off |
| Auto close inline display | Off |
| Start measurement without further preparation | On |
| Wait for user to start | Off |
| Start measurements | Single measurement |

### Routine

| | |
|---|---|
| Slice group | 1 |
| Slices | 72 |
| Dist. factor | 0 % |
| Position | L0.0 P0.0 H6.0 mm |
| Orientation | T > C-16.0 |
| Phase enc. dir. | A >> P |
| AutoAlign | Head > Brain |
| Phase oversampling | 0 % |
| FoV read | 208 mm |
| FoV phase | 100.0 % |
| Slice thickness | 2.00 mm |
| TR | 8000 ms |
| TE | 66.00 ms |
| Multi-band accel. factor | 1 |
| Filter | None |
| Coil elements | HC1-7;NC1,2 |

### Contrast - Common

| | |
|---|---|
| TR | 8000 ms |
| TE | 66.00 ms |
| MTC | Off |
| Magn. preparation | None |
| Flip angle | 90 deg |
| Refocus flip angle | 180 deg |
| Fat suppr. | Fat sat. |
| Grad. rev. fat suppr. | Disabled |

### Contrast - Dynamic

| | |
|---|---|
| Averaging mode | Long term |
| Reconstruction | Magnitude |
| Measurements | 1 |
| Delay in TR | 0 ms |
| Multiple series | Off |

### Resolution - Common

| | |
|---|---|
| FoV read | 208 mm |
| FoV phase | 100.0 % |
| Slice thickness | 2.00 mm |
| Base resolution | 104 |
| Phase resolution | 100 % |
| Phase partial Fourier | Off |
| Interpolation | Off |

### Resolution - iPAT

| | |
|---|---|
| PAT mode | None |

### Resolution - Filter Image

| | |
|---|---|
| Distortion Corr. | Off |
| Prescan Normalize | Off |

### Resolution - Filter Rawdata

| | |
|---|---|
| Raw filter | Off |
| Elliptical filter | Off |
| Hamming | Off |

### Geometry - Common

| | |
|---|---|
| Slice group | 1 |
| Slices | 72 |
| Dist. factor | 0 % |
| Position | L0.0 P0.0 H6.0 mm |
| Orientation | T > C-16.0 |
| Phase enc. dir. | A >> P |
| FoV read | 208 mm |
| FoV phase | 100.0 % |
| Slice thickness | 2.00 mm |
| TR | 8000 ms |
| Multi-slice mode | Interleaved |
| Series | Interleaved |
| Multi-band accel. factor | 1 |

### Geometry - AutoAlign

| | |
|---|---|
| Slice group | 1 |
| Position | L0.0 P0.0 H6.0 mm |
| Orientation | T > C-16.0 |
| Phase enc. dir. | A >> P |
| AutoAlign | Head > Brain |
| Initial Position | L0.0 P0.0 H6.0 |
| L | 0.0 mm |
| P | 0.0 mm |
| H | 6.0 mm |
| Initial Rotation | 0.00 deg |
| Initial Orientation | T > C |
| T > C | -16.0 |
| > S | 0.0 |

### Geometry - Saturation

| | |
|---|---|
| Fat suppr. | Fat sat. |
| Grad. rev. fat suppr. | Disabled |
| Special sat. | None |

### System - Miscellaneous

| | |
|---|---|
| Positioning mode | REF |
| Table position | H |
| Table position | 0 mm |
| MSMA | S - C - T |
| Sagittal | R >> L |
| Coronal | A >> P |
| Transversal | F >> H |
| Coil Combine Mode | Sum of Squares |
| Matrix Optimization | Off |
| AutoAlign | Head > Brain |
| Coil Select Mode | Off - All |

### System - Adjustments

| | |
|---|---|
| B0 Shim mode | Standard |
| B1 Shim mode | TrueForm |
| Adjust with body coil | Off |

#### System - Adjustments

| | |
|---|---|
| Confirm freq. adjustment | Off |
| Assume Dominant Fat | Off |
| Assume Silicone | Off |
| Adjustment Tolerance | Auto |

#### System - Adjust Volume

| | |
|---|---|
| Position | L0.0 P0.0 H6.0 mm |
| Orientation | T > C-16.0 |
| Rotation | 0.00 deg |
| A >> P | 208 mm |
| R >> L | 208 mm |
| F >> H | 144 mm |
| Reset | Off |

#### System - pTx Volumes

| | |
|---|---|
| B1 Shim mode | TrueForm |
| Excitation | Standard |

#### System - Tx/Rx

| | |
|---|---|
| Frequency 1H | 123.260801 MHz |
| Correction factor | 1 |
| Gain | High |
| Img. Scale Cor. | 1.000 |
| Reset | Off |
| ? Ref. amplitude 1H | 0.000 V |

#### Physio - Signal1

| | |
|---|---|
| 1st Signal/Mode | None |
| TR | 8000 ms |
| Multi-band accel. factor | 1 |

#### BOLD

| | |
|---|---|
| GLM Statistics | Off |
| Dynamic t-maps | Off |
| Ignore meas. at start | 0 |
| Ignore after transition | 0 |
| Model transition states | Off |
| Temp. highpass filter | Off |
| Threshold | 4.00 |
| Paradigm size | 3 |
| Meas[1] | Baseline |
| Meas[2] | Baseline |
| Meas[3] | Active |
| Motion correction | Off |
| Spatial filter | Off |
| Measurements | 1 |
| Delay in TR | 0 ms |
| Multiple series | Off |

#### Sequence - Part 1

| | |
|---|---|
| Introduction | Off |
| Contrasts | 1 |
| Multi-slice mode | Interleaved |
| Free echo spacing | On |
| Echo spacing | 0.58 ms |
| Bandwidth | 2290 Hz/Px |

#### Sequence - Part 2

| | |
|---|---|
| EPI factor | 104 |
| RF pulse type | Normal |
| Gradient mode | Performance |
| Excitation | Standard |

#### Sequence - Special

| | |
|---|---|
| SENSE1 coil combine | Off |
| Invert RO/PE polarity | On |
| Disable freq. update | Off |
| Force equal slice timing | Off |
| FFT scale factor | 1.00 |
| Physio recording | Off |
| Triggering scheme | Standard |

**\\USER\Holmes\PCY\CMRR Holmes_Yale_McLean_Jan2022\rfMRI_REST_AP**

TA: 6:40 PM: REF Voxel size: 2.0×2.0×2.0 mmPAT: Off Rel. SNR: 1.00 : epfid

### Properties

| | |
|---|---|
| Prio recon | Off |
| Load images to viewer | On |
| Inline movie | Off |
| Auto store images | On |
| Load images to stamp segments | Off |
| Load images to graphic segments | Off |
| Auto open inline display | On |
| Auto close inline display | Off |
| Start measurement without further preparation | Off |
| Wait for user to start | On |
| Start measurements | Single measurement |

### Routine

| | |
|---|---|
| Slice group | 1 |
| Slices | 72 |
| Dist. factor | 0 % |
| Position | L0.0 P0.0 H6.0 mm |
| Orientation | T > C-16.0 |
| Phase enc. dir. | A >> P |
| AutoAlign | Head > Brain |
| Phase oversampling | 0 % |
| FoV read | 208 mm |
| FoV phase | 100.0 % |
| Slice thickness | 2.00 mm |
| TR | 800 ms |
| TE | 37.00 ms |
| Multi-band accel. factor | 8 |
| Filter | None |
| Coil elements | HC1-7;NC1,2 |

### Contrast - Common

| | |
|---|---|
| TR | 800 ms |
| TE | 37.00 ms |
| MTC | Off |
| Magn. preparation | None |
| Flip angle | 52 deg |
| Fat suppr. | Fat sat. |

### Contrast - Dynamic

| | |
|---|---|
| Averaging mode | Long term |
| Reconstruction | Magnitude |
| Measurements | 488 |
| Delay in TR | 0 ms |
| Multiple series | Off |

### Resolution - Common

| | |
|---|---|
| FoV read | 208 mm |
| FoV phase | 100.0 % |
| Slice thickness | 2.00 mm |
| Base resolution | 104 |
| Phase resolution | 100 % |
| Phase partial Fourier | Off |
| Interpolation | Off |

### Resolution - iPAT

| | |
|---|---|
| PAT mode | None |

### Resolution - Filter Image

| | |
|---|---|
| Distortion Corr. | Off |

### Resolution - Filter Image

| | |
|---|---|
| Prescan Normalize | Off |

### Resolution - Filter Rawdata

| | |
|---|---|
| Raw filter | Off |
| Elliptical filter | Off |
| Hamming | Off |

### Geometry - Common

| | |
|---|---|
| Slice group | 1 |
| Slices | 72 |
| Dist. factor | 0 % |
| Position | L0.0 P0.0 H6.0 mm |
| Orientation | T > C-16.0 |
| Phase enc. dir. | A >> P |
| FoV read | 208 mm |
| FoV phase | 100.0 % |
| Slice thickness | 2.00 mm |
| TR | 800 ms |
| Multi-slice mode | Interleaved |
| Series | Interleaved |
| Multi-band accel. factor | 8 |

### Geometry - AutoAlign

| | |
|---|---|
| Slice group | 1 |
| Position | L0.0 P0.0 H6.0 mm |
| Orientation | T > C-16.0 |
| Phase enc. dir. | A >> P |
| AutoAlign | Head > Brain |
| Initial Position | L0.0 P0.0 H6.0 |
| L | 0.0 mm |
| P | 0.0 mm |
| H | 6.0 mm |
| Initial Rotation | 0.00 deg |
| Initial Orientation | T > C |
| T > C | -16.0 |
| > S | 0.0 |

### Geometry - Saturation

| | |
|---|---|
| Fat suppr. | Fat sat. |
| Special sat. | None |

### System - Miscellaneous

| | |
|---|---|
| Positioning mode | REF |
| Table position | H |
| Table position | 0 mm |
| MSMA | S - C - T |
| Sagittal | R >> L |
| Coronal | A >> P |
| Transversal | F >> H |
| Coil Combine Mode | Sum of Squares |
| Matrix Optimization | Performance |
| AutoAlign | Head > Brain |
| Coil Select Mode | Off - All |

### System - Adjustments

| | |
|---|---|
| B0 Shim mode | Standard |
| B1 Shim mode | TrueForm |
| Adjust with body coil | Off |
| Confirm freq. adjustment | Off |
| Assume Dominant Fat | Off |

### System - Adjustments

| | |
|---|---|
| Assume Silicone | Off |
| Adjustment Tolerance | Auto |

### System - Adjust Volume

| | |
|---|---|
| Position | L0.0 P0.0 H6.0 mm |
| Orientation | T > C-16.0 |
| Rotation | 0.00 deg |
| A >> P | 208 mm |
| R >> L | 208 mm |
| F >> H | 144 mm |
| Reset | Off |

### System - pTx Volumes

| | |
|---|---|
| B1 Shim mode | TrueForm |
| Excitation | Standard |

### System - Tx/Rx

| | |
|---|---|
| Frequency 1H | 123.260801 MHz |
| Correction factor | 1 |
| Gain | High |
| Img. Scale Cor. | 1.000 |
| Reset | Off |
| ? Ref. amplitude 1H | 0.000 V |

### Physio - Signal1

| | |
|---|---|
| 1st Signal/Mode | None |
| TR | 800 ms |
| Multi-band accel. factor | 8 |

### BOLD

| | |
|---|---|
| GLM Statistics | Off |
| Dynamic t-maps | Off |
| Ignore meas. at start | 0 |
| Ignore after transition | 0 |
| Model transition states | On |
| Temp. highpass filter | On |
| Threshold | 4.00 |
| Paradigm size | 3 |
| Meas[1] | Baseline |
| Meas[2] | Baseline |
| Meas[3] | Active |
| Motion correction | Off |
| Spatial filter | Off |
| Measurements | 488 |
| Delay in TR | 0 ms |
| Multiple series | Off |

### Sequence - Part 1

| | |
|---|---|
| Introduction | Off |
| Contrasts | 1 |
| Flow comp. | No |
| Multi-slice mode | Interleaved |
| Free echo spacing | Off |
| Echo spacing | 0.58 ms |
| Bandwidth | 2290 Hz/Px |

### Sequence - Part 2

| | |
|---|---|
| EPI factor | 104 |
| Gradient mode | Performance |
| Excitation | Standard |
| RF spoiling | Off |

### Sequence - Special

| | |
|---|---|
| Excite pulse duration | 6600 us |
| Single-band images | On |
| MB LeakBlock kernel | On |
| MB dual kernel | Off |
| MB RF phase scramble | Off |
| SENSE1 coil combine | Off |
| Invert RO/PE polarity | Off |
| Disable freq. update | Off |
| Force equal slice timing | Off |
| Online multi-band recon. | Online |
| FFT scale factor | 1.00 |
| Physio recording | DICOM |
| Triggering scheme | Standard |

**\\USER\Holmes\PCY\CMRR Holmes_Yale_McLean_Jan2022\rfMRI_REST_PA**

TA: 6:40 PM: REF Voxel size: 2.0×2.0×2.0 mmPAT: Off Rel. SNR: 1.00 : epfid

### Properties

| | |
|---|---|
| Prio recon | Off |
| Load images to viewer | On |
| Inline movie | Off |
| Auto store images | On |
| Load images to stamp segments | Off |
| Load images to graphic segments | Off |
| Auto open inline display | Off |
| Auto close inline display | Off |
| Start measurement without further preparation | On |
| Wait for user to start | On |
| Start measurements | Single measurement |

### Routine

| | |
|---|---|
| Slice group | 1 |
| Slices | 72 |
| Dist. factor | 0 % |
| Position | L0.0 P0.0 H6.0 mm |
| Orientation | T > C-16.0 |
| Phase enc. dir. | A >> P |
| AutoAlign | Head > Brain |
| Phase oversampling | 0 % |
| FoV read | 208 mm |
| FoV phase | 100.0 % |
| Slice thickness | 2.00 mm |
| TR | 800 ms |
| TE | 37.00 ms |
| Multi-band accel. factor | 8 |
| Filter | None |
| Coil elements | HC1-7;NC1,2 |

### Contrast - Common

| | |
|---|---|
| TR | 800 ms |
| TE | 37.00 ms |
| MTC | Off |
| Magn. preparation | None |
| Flip angle | 52 deg |
| Fat suppr. | Fat sat. |

### Contrast - Dynamic

| | |
|---|---|
| Averaging mode | Long term |
| Reconstruction | Magnitude |
| Measurements | 488 |
| Delay in TR | 0 ms |
| Multiple series | Off |

### Resolution - Common

| | |
|---|---|
| FoV read | 208 mm |
| FoV phase | 100.0 % |
| Slice thickness | 2.00 mm |
| Base resolution | 104 |
| Phase resolution | 100 % |
| Phase partial Fourier | Off |
| Interpolation | Off |

### Resolution - iPAT

| | |
|---|---|
| PAT mode | None |

### Resolution - Filter Image

| | |
|---|---|
| Distortion Corr. | Off |

### Resolution - Filter Image

| | |
|---|---|
| Prescan Normalize | Off |

### Resolution - Filter Rawdata

| | |
|---|---|
| Raw filter | Off |
| Elliptical filter | Off |
| Hamming | Off |

### Geometry - Common

| | |
|---|---|
| Slice group | 1 |
| Slices | 72 |
| Dist. factor | 0 % |
| Position | L0.0 P0.0 H6.0 mm |
| Orientation | T > C-16.0 |
| Phase enc. dir. | A >> P |
| FoV read | 208 mm |
| FoV phase | 100.0 % |
| Slice thickness | 2.00 mm |
| TR | 800 ms |
| Multi-slice mode | Interleaved |
| Series | Interleaved |
| Multi-band accel. factor | 8 |

### Geometry - AutoAlign

| | |
|---|---|
| Slice group | 1 |
| Position | L0.0 P0.0 H6.0 mm |
| Orientation | T > C-16.0 |
| Phase enc. dir. | A >> P |
| AutoAlign | Head > Brain |
| Initial Position | L0.0 P0.0 H6.0 |
| L | 0.0 mm |
| P | 0.0 mm |
| H | 6.0 mm |
| Initial Rotation | 0.00 deg |
| Initial Orientation | T > C |
| T > C | -16.0 |
| > S | 0.0 |

### Geometry - Saturation

| | |
|---|---|
| Fat suppr. | Fat sat. |
| Special sat. | None |

### System - Miscellaneous

| | |
|---|---|
| Positioning mode | REF |
| Table position | H |
| Table position | 0 mm |
| MSMA | S - C - T |
| Sagittal | R >> L |
| Coronal | A >> P |
| Transversal | F >> H |
| Coil Combine Mode | Sum of Squares |
| Matrix Optimization | Performance |
| AutoAlign | Head > Brain |
| Coil Select Mode | Off - All |

### System - Adjustments

| | |
|---|---|
| B0 Shim mode | Standard |
| B1 Shim mode | TrueForm |
| Adjust with body coil | Off |
| Confirm freq. adjustment | Off |
| Assume Dominant Fat | Off |

#### System - Adjustments

| | |
|---|---|
| Assume Silicone | Off |
| Adjustment Tolerance | Auto |

#### System - Adjust Volume

| | |
|---|---|
| Position | L0.0 P0.0 H6.0 mm |
| Orientation | T > C-16.0 |
| Rotation | 0.00 deg |
| A >> P | 208 mm |
| R >> L | 208 mm |
| F >> H | 144 mm |
| Reset | Off |

#### System - pTx Volumes

| | |
|---|---|
| B1 Shim mode | TrueForm |
| Excitation | Standard |

#### System - Tx/Rx

| | |
|---|---|
| Frequency 1H | 123.260801 MHz |
| Correction factor | 1 |
| Gain | High |
| Img. Scale Cor. | 1.000 |
| Reset | Off |
| ? Ref. amplitude 1H | 0.000 V |

#### Physio - Signal1

| | |
|---|---|
| 1st Signal/Mode | None |
| TR | 800 ms |
| Multi-band accel. factor | 8 |

#### BOLD

| | |
|---|---|
| GLM Statistics | Off |
| Dynamic t-maps | Off |
| Ignore meas. at start | 0 |
| Ignore after transition | 0 |
| Model transition states | On |
| Temp. highpass filter | On |
| Threshold | 4.00 |
| Paradigm size | 3 |
| Meas[1] | Baseline |
| Meas[2] | Baseline |
| Meas[3] | Active |
| Motion correction | Off |
| Spatial filter | Off |
| Measurements | 488 |
| Delay in TR | 0 ms |
| Multiple series | Off |

#### Sequence - Part 1

| | |
|---|---|
| Introduction | Off |
| Contrasts | 1 |
| Flow comp. | No |
| Multi-slice mode | Interleaved |
| Free echo spacing | Off |
| Echo spacing | 0.58 ms |
| Bandwidth | 2290 Hz/Px |

#### Sequence - Part 2

| | |
|---|---|
| EPI factor | 104 |
| Gradient mode | Performance |
| Excitation | Standard |
| RF spoiling | Off |

#### Sequence - Special

| | |
|---|---|
| Excite pulse duration | 6600 us |
| Single-band images | On |
| MB LeakBlock kernel | On |
| MB dual kernel | Off |
| MB RF phase scramble | Off |
| SENSE1 coil combine | Off |
| Invert RO/PE polarity | On |
| Disable freq. update | Off |
| Force equal slice timing | Off |
| Online multi-band recon. | Online |
| FFT scale factor | 1.00 |
| Physio recording | DICOM |
| Triggering scheme | Standard |

| \\USER\Holmes\PCY\CMRR Holmes_Yale_McLean_Jan2022\Stroop_BI1_AP |
|---|
| TA: 6:58 PM: REF Voxel size: 2.0×2.0×2.0 mmPAT: Off Rel. SNR: 1.00 : epfid |

### Properties

| | |
|---|---|
| Prio recon | Off |
| Load images to viewer | On |
| Inline movie | Off |
| Auto store images | On |
| Load images to stamp segments | Off |
| Load images to graphic segments | Off |
| Auto open inline display | On |
| Auto close inline display | Off |
| Start measurement without further preparation | Off |
| Wait for user to start | On |
| Start measurements | Single measurement |

### Routine

| | |
|---|---|
| Slice group | 1 |
| Slices | 72 |
| Dist. factor | 0 % |
| Position | L0.0 P0.0 H6.0 mm |
| Orientation | T > C-16.0 |
| Phase enc. dir. | A >> P |
| AutoAlign | Head > Brain |
| Phase oversampling | 0 % |
| FoV read | 208 mm |
| FoV phase | 100.0 % |
| Slice thickness | 2.00 mm |
| TR | 800 ms |
| TE | 37.00 ms |
| Multi-band accel. factor | 8 |
| Filter | None |
| Coil elements | HC1-7;NC1,2 |

### Contrast - Common

| | |
|---|---|
| TR | 800 ms |
| TE | 37.00 ms |
| MTC | Off |
| Magn. preparation | None |
| Flip angle | 52 deg |
| Fat suppr. | Fat sat. |

### Contrast - Dynamic

| | |
|---|---|
| Averaging mode | Long term |
| Reconstruction | Magnitude |
| Measurements | 510 |
| Delay in TR | 0 ms |
| Multiple series | Off |

### Resolution - Common

| | |
|---|---|
| FoV read | 208 mm |
| FoV phase | 100.0 % |
| Slice thickness | 2.00 mm |
| Base resolution | 104 |
| Phase resolution | 100 % |
| Phase partial Fourier | Off |
| Interpolation | Off |

### Resolution - iPAT

| | |
|---|---|
| PAT mode | None |

### Resolution - Filter Image

| | |
|---|---|
| Distortion Corr. | Off |

### Resolution - Filter Image

| | |
|---|---|
| Prescan Normalize | Off |

### Resolution - Filter Rawdata

| | |
|---|---|
| Raw filter | Off |
| Elliptical filter | Off |
| Hamming | Off |

### Geometry - Common

| | |
|---|---|
| Slice group | 1 |
| Slices | 72 |
| Dist. factor | 0 % |
| Position | L0.0 P0.0 H6.0 mm |
| Orientation | T > C-16.0 |
| Phase enc. dir. | A >> P |
| FoV read | 208 mm |
| FoV phase | 100.0 % |
| Slice thickness | 2.00 mm |
| TR | 800 ms |
| Multi-slice mode | Interleaved |
| Series | Interleaved |
| Multi-band accel. factor | 8 |

### Geometry - AutoAlign

| | |
|---|---|
| Slice group | 1 |
| Position | L0.0 P0.0 H6.0 mm |
| Orientation | T > C-16.0 |
| Phase enc. dir. | A >> P |
| AutoAlign | Head > Brain |
| Initial Position | L0.0 P0.0 H6.0 |
| L | 0.0 mm |
| P | 0.0 mm |
| H | 6.0 mm |
| Initial Rotation | 0.00 deg |
| Initial Orientation | T > C |
| T > C | -16.0 |
| > S | 0.0 |

### Geometry - Saturation

| | |
|---|---|
| Fat suppr. | Fat sat. |
| Special sat. | None |

### System - Miscellaneous

| | |
|---|---|
| Positioning mode | REF |
| Table position | H |
| Table position | 0 mm |
| MSMA | S - C - T |
| Sagittal | R >> L |
| Coronal | A >> P |
| Transversal | F >> H |
| Coil Combine Mode | Sum of Squares |
| Matrix Optimization | Performance |
| AutoAlign | Head > Brain |
| Coil Select Mode | Off - All |

### System - Adjustments

| | |
|---|---|
| B0 Shim mode | Standard |
| B1 Shim mode | TrueForm |
| Adjust with body coil | Off |
| Confirm freq. adjustment | Off |
| Assume Dominant Fat | Off |

#### System - Adjustments

| | |
|---|---|
| Assume Silicone | Off |
| Adjustment Tolerance | Auto |

#### System - Adjust Volume

| | |
|---|---|
| Position | L0.0 P0.0 H6.0 mm |
| Orientation | T > C-16.0 |
| Rotation | 0.00 deg |
| A >> P | 208 mm |
| R >> L | 208 mm |
| F >> H | 144 mm |
| Reset | Off |

#### System - pTx Volumes

| | |
|---|---|
| B1 Shim mode | TrueForm |
| Excitation | Standard |

#### System - Tx/Rx

| | |
|---|---|
| Frequency 1H | 123.260801 MHz |
| Correction factor | 1 |
| Gain | High |
| Img. Scale Cor. | 1.000 |
| Reset | Off |
| ? Ref. amplitude 1H | 0.000 V |

#### Physio - Signal1

| | |
|---|---|
| 1st Signal/Mode | None |
| TR | 800 ms |
| Multi-band accel. factor | 8 |

#### BOLD

| | |
|---|---|
| GLM Statistics | Off |
| Dynamic t-maps | Off |
| Ignore meas. at start | 0 |
| Ignore after transition | 0 |
| Model transition states | On |
| Temp. highpass filter | On |
| Threshold | 4.00 |
| Paradigm size | 3 |
| Meas[1] | Baseline |
| Meas[2] | Baseline |
| Meas[3] | Active |
| Motion correction | Off |
| Spatial filter | Off |
| Measurements | 510 |
| Delay in TR | 0 ms |
| Multiple series | Off |

#### Sequence - Part 1

| | |
|---|---|
| Introduction | Off |
| Contrasts | 1 |
| Flow comp. | No |
| Multi-slice mode | Interleaved |
| Free echo spacing | Off |
| Echo spacing | 0.58 ms |
| Bandwidth | 2290 Hz/Px |

#### Sequence - Part 2

| | |
|---|---|
| EPI factor | 104 |
| Gradient mode | Performance |
| Excitation | Standard |
| RF spoiling | Off |

#### Sequence - Special

| | |
|---|---|
| Excite pulse duration | 6600 us |
| Single-band images | On |
| MB LeakBlock kernel | On |
| MB dual kernel | Off |
| MB RF phase scramble | Off |
| SENSE1 coil combine | Off |
| Invert RO/PE polarity | Off |
| Disable freq. update | Off |
| Force equal slice timing | Off |
| Online multi-band recon. | Online |
| FFT scale factor | 1.00 |
| Physio recording | DICOM |
| Triggering scheme | Standard |

**\\USER\Holmes\PCY\CMRR Holmes_Yale_McLean_Jan2022\Stroop_BI2_PA**

TA: 6:58 PM: REF Voxel size: 2.0×2.0×2.0 mmPAT: Off Rel. SNR: 1.00 : epfid

### Properties

| | |
|---|---|
| Prio recon | Off |
| Load images to viewer | On |
| Inline movie | Off |
| Auto store images | On |
| Load images to stamp segments | Off |
| Load images to graphic segments | Off |
| Auto open inline display | Off |
| Auto close inline display | Off |
| Start measurement without further preparation | On |
| Wait for user to start | On |
| Start measurements | Single measurement |

### Routine

| | |
|---|---|
| Slice group | 1 |
| Slices | 72 |
| Dist. factor | 0 % |
| Position | L0.0 P0.0 H6.0 mm |
| Orientation | T > C-16.0 |
| Phase enc. dir. | A >> P |
| AutoAlign | Head > Brain |
| Phase oversampling | 0 % |
| FoV read | 208 mm |
| FoV phase | 100.0 % |
| Slice thickness | 2.00 mm |
| TR | 800 ms |
| TE | 37.00 ms |
| Multi-band accel. factor | 8 |
| Filter | None |
| Coil elements | HC1-7;NC1,2 |

### Contrast - Common

| | |
|---|---|
| TR | 800 ms |
| TE | 37.00 ms |
| MTC | Off |
| Magn. preparation | None |
| Flip angle | 52 deg |
| Fat suppr. | Fat sat. |

### Contrast - Dynamic

| | |
|---|---|
| Averaging mode | Long term |
| Reconstruction | Magnitude |
| Measurements | 510 |
| Delay in TR | 0 ms |
| Multiple series | Off |

### Resolution - Common

| | |
|---|---|
| FoV read | 208 mm |
| FoV phase | 100.0 % |
| Slice thickness | 2.00 mm |
| Base resolution | 104 |
| Phase resolution | 100 % |
| Phase partial Fourier | Off |
| Interpolation | Off |

### Resolution - iPAT

| | |
|---|---|
| PAT mode | None |

### Resolution - Filter Image

| | |
|---|---|
| Distortion Corr. | Off |

### Resolution - Filter Image

| | |
|---|---|
| Prescan Normalize | Off |

### Resolution - Filter Rawdata

| | |
|---|---|
| Raw filter | Off |
| Elliptical filter | Off |
| Hamming | Off |

### Geometry - Common

| | |
|---|---|
| Slice group | 1 |
| Slices | 72 |
| Dist. factor | 0 % |
| Position | L0.0 P0.0 H6.0 mm |
| Orientation | T > C-16.0 |
| Phase enc. dir. | A >> P |
| FoV read | 208 mm |
| FoV phase | 100.0 % |
| Slice thickness | 2.00 mm |
| TR | 800 ms |
| Multi-slice mode | Interleaved |
| Series | Interleaved |
| Multi-band accel. factor | 8 |

### Geometry - AutoAlign

| | |
|---|---|
| Slice group | 1 |
| Position | L0.0 P0.0 H6.0 mm |
| Orientation | T > C-16.0 |
| Phase enc. dir. | A >> P |
| AutoAlign | Head > Brain |
| Initial Position | L0.0 P0.0 H6.0 |
| L | 0.0 mm |
| P | 0.0 mm |
| H | 6.0 mm |
| Initial Rotation | 0.00 deg |
| Initial Orientation | T > C |
| T > C | -16.0 |
| > S | 0.0 |

### Geometry - Saturation

| | |
|---|---|
| Fat suppr. | Fat sat. |
| Special sat. | None |

### System - Miscellaneous

| | |
|---|---|
| Positioning mode | REF |
| Table position | H |
| Table position | 0 mm |
| MSMA | S - C - T |
| Sagittal | R >> L |
| Coronal | A >> P |
| Transversal | F >> H |
| Coil Combine Mode | Sum of Squares |
| Matrix Optimization | Performance |
| AutoAlign | Head > Brain |
| Coil Select Mode | Off - All |

### System - Adjustments

| | |
|---|---|
| B0 Shim mode | Standard |
| B1 Shim mode | TrueForm |
| Adjust with body coil | Off |
| Confirm freq. adjustment | Off |
| Assume Dominant Fat | Off |

### System - Adjustments

| | |
|---|---|
| Assume Silicone | Off |
| Adjustment Tolerance | Auto |

### System - Adjust Volume

| | |
|---|---|
| Position | L0.0 P0.0 H6.0 mm |
| Orientation | T > C-16.0 |
| Rotation | 0.00 deg |
| A >> P | 208 mm |
| R >> L | 208 mm |
| F >> H | 144 mm |
| Reset | Off |

### System - pTx Volumes

| | |
|---|---|
| B1 Shim mode | TrueForm |
| Excitation | Standard |

### System - Tx/Rx

| | |
|---|---|
| Frequency 1H | 123.260801 MHz |
| Correction factor | 1 |
| Gain | High |
| Img. Scale Cor. | 1.000 |
| Reset | Off |
| ? Ref. amplitude 1H | 0.000 V |

### Physio - Signal1

| | |
|---|---|
| 1st Signal/Mode | None |
| TR | 800 ms |
| Multi-band accel. factor | 8 |

### BOLD

| | |
|---|---|
| GLM Statistics | Off |
| Dynamic t-maps | Off |
| Ignore meas. at start | 0 |
| Ignore after transition | 0 |
| Model transition states | On |
| Temp. highpass filter | On |
| Threshold | 4.00 |
| Paradigm size | 3 |
| Meas[1] | Baseline |
| Meas[2] | Baseline |
| Meas[3] | Active |
| Motion correction | Off |
| Spatial filter | Off |
| Measurements | 510 |
| Delay in TR | 0 ms |
| Multiple series | Off |

### Sequence - Part 1

| | |
|---|---|
| Introduction | Off |
| Contrasts | 1 |
| Flow comp. | No |
| Multi-slice mode | Interleaved |
| Free echo spacing | Off |
| Echo spacing | 0.58 ms |
| Bandwidth | 2290 Hz/Px |

### Sequence - Part 2

| | |
|---|---|
| EPI factor | 104 |
| Gradient mode | Performance |
| Excitation | Standard |
| RF spoiling | Off |

### Sequence - Special

| | |
|---|---|
| Excite pulse duration | 6600 us |
| Single-band images | On |
| MB LeakBlock kernel | On |
| MB dual kernel | Off |
| MB RF phase scramble | Off |
| SENSE1 coil combine | Off |
| Invert RO/PE polarity | On |
| Disable freq. update | Off |
| Force equal slice timing | Off |
| Online multi-band recon. | Online |
| FFT scale factor | 1.00 |
| Physio recording | DICOM |
| Triggering scheme | Standard |

| \\USER\Holmes\PCY\CMRR Holmes_Yale_McLean_Jan2022\Hammer_BI1_AP |
| --- |
| TA: 6:44 PM: REF Voxel size: 2.0×2.0×2.0 mmPAT: Off Rel. SNR: 1.00 : epfid |

### Properties

| | |
| --- | --- |
| Prio recon | Off |
| Load images to viewer | On |
| Inline movie | Off |
| Auto store images | On |
| Load images to stamp segments | Off |
| Load images to graphic segments | Off |
| Auto open inline display | On |
| Auto close inline display | Off |
| Start measurement without further preparation | Off |
| Wait for user to start | On |
| Start measurements | Single measurement |

### Routine

| | |
| --- | --- |
| Slice group | 1 |
| Slices | 72 |
| Dist. factor | 0 % |
| Position | L0.0 P0.0 H6.0 mm |
| Orientation | T > C-16.0 |
| Phase enc. dir. | A >> P |
| AutoAlign | Head > Brain |
| Phase oversampling | 0 % |
| FoV read | 208 mm |
| FoV phase | 100.0 % |
| Slice thickness | 2.00 mm |
| TR | 800 ms |
| TE | 37.00 ms |
| Multi-band accel. factor | 8 |
| Filter | None |
| Coil elements | HC1-7;NC1,2 |

### Contrast - Common

| | |
| --- | --- |
| TR | 800 ms |
| TE | 37.00 ms |
| MTC | Off |
| Magn. preparation | None |
| Flip angle | 52 deg |
| Fat suppr. | Fat sat. |

### Contrast - Dynamic

| | |
| --- | --- |
| Averaging mode | Long term |
| Reconstruction | Magnitude |
| Measurements | 493 |
| Delay in TR | 0 ms |
| Multiple series | Off |

### Resolution - Common

| | |
| --- | --- |
| FoV read | 208 mm |
| FoV phase | 100.0 % |
| Slice thickness | 2.00 mm |
| Base resolution | 104 |
| Phase resolution | 100 % |
| Phase partial Fourier | Off |
| Interpolation | Off |

### Resolution - iPAT

| | |
| --- | --- |
| PAT mode | None |

### Resolution - Filter Image

| | |
| --- | --- |
| Distortion Corr. | Off |

### Resolution - Filter Image

| | |
| --- | --- |
| Prescan Normalize | Off |

### Resolution - Filter Rawdata

| | |
| --- | --- |
| Raw filter | Off |
| Elliptical filter | Off |
| Hamming | Off |

### Geometry - Common

| | |
| --- | --- |
| Slice group | 1 |
| Slices | 72 |
| Dist. factor | 0 % |
| Position | L0.0 P0.0 H6.0 mm |
| Orientation | T > C-16.0 |
| Phase enc. dir. | A >> P |
| FoV read | 208 mm |
| FoV phase | 100.0 % |
| Slice thickness | 2.00 mm |
| TR | 800 ms |
| Multi-slice mode | Interleaved |
| Series | Interleaved |
| Multi-band accel. factor | 8 |

### Geometry - AutoAlign

| | |
| --- | --- |
| Slice group | 1 |
| Position | L0.0 P0.0 H6.0 mm |
| Orientation | T > C-16.0 |
| Phase enc. dir. | A >> P |
| AutoAlign | Head > Brain |
| Initial Position | L0.0 P0.0 H6.0 |
| L | 0.0 mm |
| P | 0.0 mm |
| H | 6.0 mm |
| Initial Rotation | 0.00 deg |
| Initial Orientation | T > C |
| T > C | -16.0 |
| > S | 0.0 |

### Geometry - Saturation

| | |
| --- | --- |
| Fat suppr. | Fat sat. |
| Special sat. | None |

### System - Miscellaneous

| | |
| --- | --- |
| Positioning mode | REF |
| Table position | H |
| Table position | 0 mm |
| MSMA | S - C - T |
| Sagittal | R >> L |
| Coronal | A >> P |
| Transversal | F >> H |
| Coil Combine Mode | Sum of Squares |
| Matrix Optimization | Performance |
| AutoAlign | Head > Brain |
| Coil Select Mode | Off - All |

### System - Adjustments

| | |
| --- | --- |
| B0 Shim mode | Standard |
| B1 Shim mode | TrueForm |
| Adjust with body coil | Off |
| Confirm freq. adjustment | Off |
| Assume Dominant Fat | Off |

### System - Adjustments

| | |
|---|---|
| Assume Silicone | Off |
| Adjustment Tolerance | Auto |

### System - Adjust Volume

| | |
|---|---|
| Position | L0.0 P0.0 H6.0 mm |
| Orientation | T > C-16.0 |
| Rotation | 0.00 deg |
| A >> P | 208 mm |
| R >> L | 208 mm |
| F >> H | 144 mm |
| Reset | Off |

### System - pTx Volumes

| | |
|---|---|
| B1 Shim mode | TrueForm |
| Excitation | Standard |

### System - Tx/Rx

| | |
|---|---|
| Frequency 1H | 123.260801 MHz |
| Correction factor | 1 |
| Gain | High |
| Img. Scale Cor. | 1.000 |
| Reset | Off |
| ? Ref. amplitude 1H | 0.000 V |

### Physio - Signal1

| | |
|---|---|
| 1st Signal/Mode | None |
| TR | 800 ms |
| Multi-band accel. factor | 8 |

### BOLD

| | |
|---|---|
| GLM Statistics | Off |
| Dynamic t-maps | Off |
| Ignore meas. at start | 0 |
| Ignore after transition | 0 |
| Model transition states | On |
| Temp. highpass filter | On |
| Threshold | 4.00 |
| Paradigm size | 3 |
| Meas[1] | Baseline |
| Meas[2] | Baseline |
| Meas[3] | Active |
| Motion correction | Off |
| Spatial filter | Off |
| Measurements | 493 |
| Delay in TR | 0 ms |
| Multiple series | Off |

### Sequence - Part 1

| | |
|---|---|
| Introduction | Off |
| Contrasts | 1 |
| Flow comp. | No |
| Multi-slice mode | Interleaved |
| Free echo spacing | Off |
| Echo spacing | 0.58 ms |
| Bandwidth | 2290 Hz/Px |

### Sequence - Part 2

| | |
|---|---|
| EPI factor | 104 |
| Gradient mode | Performance |
| Excitation | Standard |
| RF spoiling | Off |

### Sequence - Special

| | |
|---|---|
| Excite pulse duration | 6600 us |
| Single-band images | On |
| MB LeakBlock kernel | On |
| MB dual kernel | Off |
| MB RF phase scramble | Off |
| SENSE1 coil combine | Off |
| Invert RO/PE polarity | Off |
| Disable freq. update | Off |
| Force equal slice timing | Off |
| Online multi-band recon. | Online |
| FFT scale factor | 1.00 |
| Physio recording | DICOM |
| Triggering scheme | Standard |

| \\USER\Holmes\PCY\CMRR Holmes_Yale_McLean_Jan2022\rfMRI_REST_AP |
|---|
| TA: 6:40 PM: REF Voxel size: 2.0×2.0×2.0 mmPAT: Off Rel. SNR: 1.00 : epfid |

### Properties

| | |
|---|---|
| Prio recon | Off |
| Load images to viewer | On |
| Inline movie | Off |
| Auto store images | On |
| Load images to stamp segments | Off |
| Load images to graphic segments | Off |
| Auto open inline display | On |
| Auto close inline display | Off |
| Start measurement without further preparation | Off |
| Wait for user to start | On |
| Start measurements | Single measurement |

### Routine

| | |
|---|---|
| Slice group | 1 |
| Slices | 72 |
| Dist. factor | 0 % |
| Position | L0.0 P0.0 H6.0 mm |
| Orientation | T > C-16.0 |
| Phase enc. dir. | A >> P |
| AutoAlign | Head > Brain |
| Phase oversampling | 0 % |
| FoV read | 208 mm |
| FoV phase | 100.0 % |
| Slice thickness | 2.00 mm |
| TR | 800 ms |
| TE | 37.00 ms |
| Multi-band accel. factor | 8 |
| Filter | None |
| Coil elements | HC1-7;NC1,2 |

### Contrast - Common

| | |
|---|---|
| TR | 800 ms |
| TE | 37.00 ms |
| MTC | Off |
| Magn. preparation | None |
| Flip angle | 52 deg |
| Fat suppr. | Fat sat. |

### Contrast - Dynamic

| | |
|---|---|
| Averaging mode | Long term |
| Reconstruction | Magnitude |
| Measurements | 488 |
| Delay in TR | 0 ms |
| Multiple series | Off |

### Resolution - Common

| | |
|---|---|
| FoV read | 208 mm |
| FoV phase | 100.0 % |
| Slice thickness | 2.00 mm |
| Base resolution | 104 |
| Phase resolution | 100 % |
| Phase partial Fourier | Off |
| Interpolation | Off |

### Resolution - iPAT

| | |
|---|---|
| PAT mode | None |

### Resolution - Filter Image

| | |
|---|---|
| Distortion Corr. | Off |

### Resolution - Filter Image

| | |
|---|---|
| Prescan Normalize | Off |

### Resolution - Filter Rawdata

| | |
|---|---|
| Raw filter | Off |
| Elliptical filter | Off |
| Hamming | Off |

### Geometry - Common

| | |
|---|---|
| Slice group | 1 |
| Slices | 72 |
| Dist. factor | 0 % |
| Position | L0.0 P0.0 H6.0 mm |
| Orientation | T > C-16.0 |
| Phase enc. dir. | A >> P |
| FoV read | 208 mm |
| FoV phase | 100.0 % |
| Slice thickness | 2.00 mm |
| TR | 800 ms |
| Multi-slice mode | Interleaved |
| Series | Interleaved |
| Multi-band accel. factor | 8 |

### Geometry - AutoAlign

| | |
|---|---|
| Slice group | 1 |
| Position | L0.0 P0.0 H6.0 mm |
| Orientation | T > C-16.0 |
| Phase enc. dir. | A >> P |
| AutoAlign | Head > Brain |
| Initial Position | L0.0 P0.0 H6.0 |
| L | 0.0 mm |
| P | 0.0 mm |
| H | 6.0 mm |
| Initial Rotation | 0.00 deg |
| Initial Orientation | T > C |
| T > C | -16.0 |
| > S | 0.0 |

### Geometry - Saturation

| | |
|---|---|
| Fat suppr. | Fat sat. |
| Special sat. | None |

### System - Miscellaneous

| | |
|---|---|
| Positioning mode | REF |
| Table position | H |
| Table position | 0 mm |
| MSMA | S - C - T |
| Sagittal | R >> L |
| Coronal | A >> P |
| Transversal | F >> H |
| Coil Combine Mode | Sum of Squares |
| Matrix Optimization | Performance |
| AutoAlign | Head > Brain |
| Coil Select Mode | Off - All |

### System - Adjustments

| | |
|---|---|
| B0 Shim mode | Standard |
| B1 Shim mode | TrueForm |
| Adjust with body coil | Off |
| Confirm freq. adjustment | Off |
| Assume Dominant Fat | Off |

### System - Adjustments

| | |
|---|---|
| Assume Silicone | Off |
| Adjustment Tolerance | Auto |

### System - Adjust Volume

| | |
|---|---|
| Position | L0.0 P0.0 H6.0 mm |
| Orientation | T > C-16.0 |
| Rotation | 0.00 deg |
| A >> P | 208 mm |
| R >> L | 208 mm |
| F >> H | 144 mm |
| Reset | Off |

### System - pTx Volumes

| | |
|---|---|
| B1 Shim mode | TrueForm |
| Excitation | Standard |

### System - Tx/Rx

| | |
|---|---|
| Frequency 1H | 123.260801 MHz |
| Correction factor | 1 |
| Gain | High |
| Img. Scale Cor. | 1.000 |
| Reset | Off |
| ? Ref. amplitude 1H | 0.000 V |

### Physio - Signal1

| | |
|---|---|
| 1st Signal/Mode | None |
| TR | 800 ms |
| Multi-band accel. factor | 8 |

### BOLD

| | |
|---|---|
| GLM Statistics | Off |
| Dynamic t-maps | Off |
| Ignore meas. at start | 0 |
| Ignore after transition | 0 |
| Model transition states | On |
| Temp. highpass filter | On |
| Threshold | 4.00 |
| Paradigm size | 3 |
| Meas[1] | Baseline |
| Meas[2] | Baseline |
| Meas[3] | Active |
| Motion correction | Off |
| Spatial filter | Off |
| Measurements | 488 |
| Delay in TR | 0 ms |
| Multiple series | Off |

### Sequence - Part 1

| | |
|---|---|
| Introduction | Off |
| Contrasts | 1 |
| Flow comp. | No |
| Multi-slice mode | Interleaved |
| Free echo spacing | Off |
| Echo spacing | 0.58 ms |
| Bandwidth | 2290 Hz/Px |

### Sequence - Part 2

| | |
|---|---|
| EPI factor | 104 |
| Gradient mode | Performance |
| Excitation | Standard |
| RF spoiling | Off |

### Sequence - Special

| | |
|---|---|
| Excite pulse duration | 6600 us |
| Single-band images | On |
| MB LeakBlock kernel | On |
| MB dual kernel | Off |
| MB RF phase scramble | Off |
| SENSE1 coil combine | Off |
| Invert RO/PE polarity | Off |
| Disable freq. update | Off |
| Force equal slice timing | Off |
| Online multi-band recon. | Online |
| FFT scale factor | 1.00 |
| Physio recording | DICOM |
| Triggering scheme | Standard |

**\\USER\Holmes\PCY\CMRR Holmes_Yale_McLean_Jan2022\rfMRI_REST_PA**

TA: 6:40 PM: REF Voxel size: 2.0×2.0×2.0 mmPAT: Off Rel. SNR: 1.00 : epfid

### Properties

| | |
|---|---|
| Prio recon | Off |
| Load images to viewer | On |
| Inline movie | Off |
| Auto store images | On |
| Load images to stamp segments | Off |
| Load images to graphic segments | Off |
| Auto open inline display | Off |
| Auto close inline display | Off |
| Start measurement without further preparation | On |
| Wait for user to start | On |
| Start measurements | Single measurement |

### Routine

| | |
|---|---|
| Slice group | 1 |
| Slices | 72 |
| Dist. factor | 0 % |
| Position | L0.0 P0.0 H6.0 mm |
| Orientation | T > C-16.0 |
| Phase enc. dir. | A >> P |
| AutoAlign | Head > Brain |
| Phase oversampling | 0 % |
| FoV read | 208 mm |
| FoV phase | 100.0 % |
| Slice thickness | 2.00 mm |
| TR | 800 ms |
| TE | 37.00 ms |
| Multi-band accel. factor | 8 |
| Filter | None |
| Coil elements | HC1-7;NC1,2 |

### Contrast - Common

| | |
|---|---|
| TR | 800 ms |
| TE | 37.00 ms |
| MTC | Off |
| Magn. preparation | None |
| Flip angle | 52 deg |
| Fat suppr. | Fat sat. |

### Contrast - Dynamic

| | |
|---|---|
| Averaging mode | Long term |
| Reconstruction | Magnitude |
| Measurements | 488 |
| Delay in TR | 0 ms |
| Multiple series | Off |

### Resolution - Common

| | |
|---|---|
| FoV read | 208 mm |
| FoV phase | 100.0 % |
| Slice thickness | 2.00 mm |
| Base resolution | 104 |
| Phase resolution | 100 % |
| Phase partial Fourier | Off |
| Interpolation | Off |

### Resolution - iPAT

| | |
|---|---|
| PAT mode | None |

### Resolution - Filter Image

| | |
|---|---|
| Distortion Corr. | Off |

### Resolution - Filter Image

| | |
|---|---|
| Prescan Normalize | Off |

### Resolution - Filter Rawdata

| | |
|---|---|
| Raw filter | Off |
| Elliptical filter | Off |
| Hamming | Off |

### Geometry - Common

| | |
|---|---|
| Slice group | 1 |
| Slices | 72 |
| Dist. factor | 0 % |
| Position | L0.0 P0.0 H6.0 mm |
| Orientation | T > C-16.0 |
| Phase enc. dir. | A >> P |
| FoV read | 208 mm |
| FoV phase | 100.0 % |
| Slice thickness | 2.00 mm |
| TR | 800 ms |
| Multi-slice mode | Interleaved |
| Series | Interleaved |
| Multi-band accel. factor | 8 |

### Geometry - AutoAlign

| | |
|---|---|
| Slice group | 1 |
| Position | L0.0 P0.0 H6.0 mm |
| Orientation | T > C-16.0 |
| Phase enc. dir. | A >> P |
| AutoAlign | Head > Brain |
| Initial Position | L0.0 P0.0 H6.0 |
| L | 0.0 mm |
| P | 0.0 mm |
| H | 6.0 mm |
| Initial Rotation | 0.00 deg |
| Initial Orientation | T > C |
| T > C | -16.0 |
| > S | 0.0 |

### Geometry - Saturation

| | |
|---|---|
| Fat suppr. | Fat sat. |
| Special sat. | None |

### System - Miscellaneous

| | |
|---|---|
| Positioning mode | REF |
| Table position | H |
| Table position | 0 mm |
| MSMA | S - C - T |
| Sagittal | R >> L |
| Coronal | A >> P |
| Transversal | F >> H |
| Coil Combine Mode | Sum of Squares |
| Matrix Optimization | Performance |
| AutoAlign | Head > Brain |
| Coil Select Mode | Off - All |

### System - Adjustments

| | |
|---|---|
| B0 Shim mode | Standard |
| B1 Shim mode | TrueForm |
| Adjust with body coil | Off |
| Confirm freq. adjustment | Off |
| Assume Dominant Fat | Off |

### System - Adjustments

| | |
|---|---|
| Assume Silicone | Off |
| Adjustment Tolerance | Auto |

### System - Adjust Volume

| | |
|---|---|
| Position | L0.0 P0.0 H6.0 mm |
| Orientation | T > C-16.0 |
| Rotation | 0.00 deg |
| A >> P | 208 mm |
| R >> L | 208 mm |
| F >> H | 144 mm |
| Reset | Off |

### System - pTx Volumes

| | |
|---|---|
| B1 Shim mode | TrueForm |
| Excitation | Standard |

### System - Tx/Rx

| | |
|---|---|
| Frequency 1H | 123.260801 MHz |
| Correction factor | 1 |
| Gain | High |
| Img. Scale Cor. | 1.000 |
| Reset | Off |
| ? Ref. amplitude 1H | 0.000 V |

### Physio - Signal1

| | |
|---|---|
| 1st Signal/Mode | None |
| TR | 800 ms |
| Multi-band accel. factor | 8 |

### BOLD

| | |
|---|---|
| GLM Statistics | Off |
| Dynamic t-maps | Off |
| Ignore meas. at start | 0 |
| Ignore after transition | 0 |
| Model transition states | On |
| Temp. highpass filter | On |
| Threshold | 4.00 |
| Paradigm size | 3 |
| Meas[1] | Baseline |
| Meas[2] | Baseline |
| Meas[3] | Active |
| Motion correction | Off |
| Spatial filter | Off |
| Measurements | 488 |
| Delay in TR | 0 ms |
| Multiple series | Off |

### Sequence - Part 1

| | |
|---|---|
| Introduction | Off |
| Contrasts | 1 |
| Flow comp. | No |
| Multi-slice mode | Interleaved |
| Free echo spacing | Off |
| Echo spacing | 0.58 ms |
| Bandwidth | 2290 Hz/Px |

### Sequence - Part 2

| | |
|---|---|
| EPI factor | 104 |
| Gradient mode | Performance |
| Excitation | Standard |
| RF spoiling | Off |

### Sequence - Special

| | |
|---|---|
| Excite pulse duration | 6600 us |
| Single-band images | On |
| MB LeakBlock kernel | On |
| MB dual kernel | Off |
| MB RF phase scramble | Off |
| SENSE1 coil combine | Off |
| Invert RO/PE polarity | On |
| Disable freq. update | Off |
| Force equal slice timing | Off |
| Online multi-band recon. | Online |
| FFT scale factor | 1.00 |
| Physio recording | DICOM |
| Triggering scheme | Standard |
