## Appendix for "The Transdiagnostic Connectome Project: a richly phenotyped open dataset for advancing the study of brain-behavior relationships in psychiatry": Holmes_Yale_McLean_Dec2019_NOEXPIRATION.pdf

### Table of contents

\\Research
Investigators
Baker
Holmes_Yale_McLean_Dec2019_NOEXPIRATION
Localizer
AAHScout
Localizer_aligned
T1_vNav_setter
T1_MEMPRAGE_GSP_vNavTrk
T2_vNav_setter
T2_SPACE_GSP_vNavTrk
SpinEchoFieldMap_AP
SpinEchoFieldMap_PA
rfMRI_REST_AP
rfMRI_REST_PA
Stroop_Bl1_AP
Stroop_Bl2_PA
Hammer_Bl1_AP
rfMRI_REST_AP
rfMRI_REST_PA
T1_MEMPRAGE_64ch
t2_tse_dark-fluid_tra
pd+t2_tse_tra
resolve_4scan_trace_tra_p2_192

### Sequence - Part 2

| | |
|---|---|
| Segments | 1 |
| Acoustic noise reduction | None |
| RF pulse type | Normal |
| Gradient mode | Fast |
| Excitation | Slice-sel. |
| RF spoiling | On |

### Sequence - Assistant

| | |
|---|---|
| Mode | Off |
| Allowed delay | 0 s |

**\\Research\Investigators\Baker\Holmes_Yale_McLean_Dec2019_NOEXPIRATION\AAHScout**

### Sequence - Part 2

| | |
|---|---|
| RF spoiling | On |

### Sequence - Assistant

| | |
|---|---|
| Mode | Off |

| \\Research\Investigators\Baker\Holmes_Yale_McLean_Dec2019_NOEXPIRATION\Localizer_aligned |
|---|
| TA: 0:21 PM: REF Voxel size: 1.2×1.2×5.0 mmPAT: Off Rel. SNR: 1.00 : fl |

### Sequence - Part 2

| | |
|---|---|
| Segments | 1 |
| Acoustic noise reduction | None |
| RF pulse type | Normal |
| Gradient mode | Fast |
| Excitation | Slice-sel. |
| RF spoiling | On |

### Sequence - Assistant

| | |
|---|---|
| Mode | Off |
| Allowed delay | 0 s |

| \\Research\Investigators\Baker\Holmes_Yale_McLean_Dec2019_NOEXPIRATION\T1_vNav_setter |
|---|
| TA: 0.3 s PM: FIX Voxel size: 8.0×8.0×8.0 mmRel. SNR: 1.00 : ABCD |

### Sequence - Part 2

| | |
|---|---|
| EPI factor | 32 |
| RF pulse type | Normal |
| Gradient mode | Fast |
| Excitation | Non-sel. |
| RF spoiling | On |

### Sequence - Special

| | |
|---|---|
| Protocol filename | MPRAGE |

**\\Research\Investigators\Baker\Holmes_Yale_McLean_Dec2019_NOEXPIRATION\T1_MEMPRAGE_GSP_vNavTrk**

TA: 2:23 PM: FIX Voxel size: 1.2×1.2×1.2 mmPAT: 4 Rel. SNR: 1.00 : tfl

### Properties

| | |
|---|---|
| Prio recon | Off |
| Load images to viewer | On |
| Inline movie | Off |
| Auto store images | On |
| Load images to stamp segments | Off |
| Load images to graphic segments | On |
| Auto open inline display | Off |
| Auto close inline display | Off |
| Start measurement without further preparation | On |
| Wait for user to start | Off |
| Start measurements | Single measurement |

### Routine

| | |
|---|---|
| Slab group | 1 |
| Slabs | 1 |
| Dist. factor | 50 % |
| Position | Isocenter |
| Orientation | Sagittal |
| Phase enc. dir. | A >> P |
| AutoAlign | Head > Brain |
| Phase oversampling | 0 % |
| Slice oversampling | 0.0 % |
| Slices per slab | 144 |
| FoV read | 230 mm |
| FoV phase | 100.0 % |
| Slice thickness | 1.20 mm |
| TR | 2200.0 ms |
| TE 1 | 1.57 ms |
| TE 2 | 3.39 ms |
| TE 3 | 5.21 ms |
| TE 4 | 7.03 ms |
| Averages | 1 |
| Concatenations | 1 |
| Filter | Prescan Normalize |
| Coil elements | HC1-7;NC1,2 |

### Contrast - Common

| | |
|---|---|
| TR | 2200.0 ms |
| TE 1 | 1.57 ms |
| TE 2 | 3.39 ms |
| TE 3 | 5.21 ms |
| TE 4 | 7.03 ms |
| Magn. preparation | Non-sel. IR |
| TI | 1100 ms |
| Flip angle | 7.0 deg |
| Fat suppr. | None |
| Water suppr. | None |

### Contrast - Dynamic

| | |
|---|---|
| Averages | 1 |
| Averaging mode | Long term |
| Reconstruction | Magnitude |
| Measurements | 1 |
| Multiple series | Each measurement |

### Resolution - Common

| | |
|---|---|
| FoV read | 230 mm |
| FoV phase | 100.0 % |

### Resolution - Common

| | |
|---|---|
| Slice thickness | 1.20 mm |
| Base resolution | 192 |
| Phase resolution | 100 % |
| Slice resolution | 100 % |
| Phase partial Fourier | 6/8 |
| Slice partial Fourier | Off |
| Interpolation | Off |

### Sequence - Assistant

| | |
|---|---|
| Mode | Off |

**\\Research\Investigators\Baker\Holmes_Yale_McLean_Dec2019_NOEXPIRATION\T2_vNav_setter**

TA: 0.3 s PM: FIX Voxel size: 8.0×8.0×8.0 mmRel. SNR: 1.00 : ABCD

### Properties

| | |
|---|---|
| Prio recon | Off |
| Load images to viewer | On |
| Inline movie | Off |
| Auto store images | On |
| Load images to stamp segments | Off |
| Load images to graphic segments | Off |
| Auto open inline display | Off |
| Auto close inline display | Off |
| Start measurement without further preparation | On |
| Wait for user to start | Off |
| Start measurements | Single measurement |

#### Sequence - Part 2

| | |
|---|---|
| EPI factor | 32 |
| RF pulse type | Normal |
| Gradient mode | Fast |
| Excitation | Non-sel. |
| RF spoiling | On |

#### Sequence - Special

| | |
|---|---|
| Protocol filename | T2-SPACE |

**\\Research\Investigators\Baker\Holmes_Yale_McLean_Dec2019_NOEXPIRATION\T2_SPACE_GSP_vNavTrk**

### System - Adjustments

| | |
|---|---|
| B0 Shim mode | Standard |
| B1 Shim mode | TrueForm |

### System - Adjustments

| | |
|---|---|
| Adjust with body coil | On |
| Confirm freq. adjustment | Off |
| Assume Dominant Fat | Off |
| Assume Silicone | Off |
| Adjustment Tolerance | Auto |

### System - Adjust Volume

| | |
|---|---|
| Position | L0.0 P0.0 H6.0 mm |
| Orientation | T > C-16.0 |
| Rotation | 0.00 deg |
| A >> P | 208 mm |
| R >> L | 208 mm |
| F >> H | 144 mm |
| Reset | Off |

### System - pTx Volumes

| | |
|---|---|
| B1 Shim mode | TrueForm |
| Excitation | Standard |

### System - Tx/Rx

| | |
|---|---|
| Frequency 1H | 123.261582 MHz |
| Correction factor | 1 |
| Gain | High |
| Img. Scale Cor. | 1.000 |
| Reset | Off |
| ? Ref. amplitude 1H | 0.000 V |

### Physio - Signal1

| | |
|---|---|
| 1st Signal/Mode | None |
| TR | 8000 ms |
| Multi-band accel. factor | 1 |

### BOLD

| | |
|---|---|
| GLM Statistics | Off |
| Dynamic t-maps | Off |
| Ignore meas. at start | 0 |
| Ignore after transition | 0 |
| Model transition states | Off |
| Temp. highpass filter | Off |
| Threshold | 4.00 |
| Paradigm size | 3 |
| Meas[1] | Baseline |
| Meas[2] | Baseline |
| Meas[3] | Active |
| Motion correction | Off |
| Spatial filter | Off |
| Measurements | 1 |
| Delay in TR | 0 ms |
| Multiple series | Off |

### Sequence - Part 1

| | |
|---|---|
| Introduction | Off |
| Contrasts | 1 |
| Multi-slice mode | Interleaved |
| Free echo spacing | On |
| Echo spacing | 0.58 ms |
| Bandwidth | 2290 Hz/Px |

### Sequence - Part 2

| | |
|---|---|
| EPI factor | 104 |
| RF pulse type | Normal |
| Gradient mode | Performance |
| Excitation | Standard |

### Sequence - Special

| | |
|---|---|
| SENSE1 coil combine | Off |
| Invert RO/PE polarity | Off |
| Disable freq. update | Off |
| Force equal slice timing | Off |
| FFT scale factor | 1.00 |
| Physio recording | Off |
| Triggering scheme | Standard |

**\\Research\Investigators\Baker\Holmes_Yale_McLean_Dec2019_NOEXPIRATION\SpinEchoFieldMap_PA**

### System - Adjustments

| | |
|---|---|
| B0 Shim mode | Standard |
| B1 Shim mode | TrueForm |

### System - Adjustments

| | |
|---|---|
| Adjust with body coil | On |
| Confirm freq. adjustment | Off |
| Assume Dominant Fat | Off |
| Assume Silicone | Off |
| Adjustment Tolerance | Auto |

### System - Adjust Volume

| | |
|---|---|
| Position | L0.0 P0.0 H6.0 mm |
| Orientation | T > C-16.0 |
| Rotation | 0.00 deg |
| A >> P | 208 mm |
| R >> L | 208 mm |
| F >> H | 144 mm |
| Reset | Off |

### System - pTx Volumes

| | |
|---|---|
| B1 Shim mode | TrueForm |
| Excitation | Standard |

### System - Tx/Rx

| | |
|---|---|
| Frequency 1H | 123.261582 MHz |
| Correction factor | 1 |
| Gain | High |
| Img. Scale Cor. | 1.000 |
| Reset | Off |
| ? Ref. amplitude 1H | 0.000 V |

### Physio - Signal1

| | |
|---|---|
| 1st Signal/Mode | None |
| TR | 8000 ms |
| Multi-band accel. factor | 1 |

### BOLD

| | |
|---|---|
| GLM Statistics | Off |
| Dynamic t-maps | Off |
| Ignore meas. at start | 0 |
| Ignore after transition | 0 |
| Model transition states | Off |
| Temp. highpass filter | Off |
| Threshold | 4.00 |
| Paradigm size | 3 |
| Meas[1] | Baseline |
| Meas[2] | Baseline |
| Meas[3] | Active |
| Motion correction | Off |
| Spatial filter | Off |
| Measurements | 1 |
| Delay in TR | 0 ms |
| Multiple series | Off |

### Sequence - Part 1

| | |
|---|---|
| Introduction | Off |
| Contrasts | 1 |
| Multi-slice mode | Interleaved |
| Free echo spacing | On |
| Echo spacing | 0.58 ms |
| Bandwidth | 2290 Hz/Px |

### Sequence - Part 2

| | |
|---|---|
| EPI factor | 104 |
| RF pulse type | Normal |
| Gradient mode | Performance |
| Excitation | Standard |

### Sequence - Special

| | |
|---|---|
| SENSE1 coil combine | Off |
| Invert RO/PE polarity | On |
| Disable freq. update | Off |
| Force equal slice timing | Off |
| FFT scale factor | 1.00 |
| Physio recording | Off |
| Triggering scheme | Standard |

**\\Research\Investigators\Baker\Holmes_Yale_McLean_Dec2019_NOEXPIRATION\rfMRI_REST_AP**

**\\Research\Investigators\Baker\Holmes_Yale_McLean_Dec2019_NOEXPIRATION\T1_MEMPRAGE_64ch**

TA: 6:02 PM: REF Voxel size: 1.0×1.0×1.0 mmPAT: 2 Rel. SNR: 1.00 : tfl_me

### Properties

| | |
|---|---|
| Prio recon | Off |
| Load images to viewer | On |
| Inline movie | Off |
| Auto store images | On |
| Load images to stamp segments | Off |
| Load images to graphic segments | Off |
| Auto open inline display | Off |
| Auto close inline display | Off |
| Start measurement without further preparation | Off |
| Wait for user to start | On |
| Start measurements | Single measurement |

### Routine

| | |
|---|---|
| Slab group | 1 |
| Slabs | 1 |
| Dist. factor | 50 % |
| Position | R2.2 P1.7 F31.8 mm |
| Orientation | Sagittal |
| Phase enc. dir. | A >> P |
| AutoAlign | Head > Brain |
| Phase oversampling | 0 % |
| Slice oversampling | 0.0 % |
| Slices per slab | 176 |
| FoV read | 256 mm |
| FoV phase | 100.0 % |
| Slice thickness | 1.00 mm |
| TR | 2530.0 ms |
| TE 1 | 1.69 ms |
| TE 2 | 3.55 ms |
| TE 3 | 5.41 ms |
| TE 4 | 7.27 ms |
| Averages | 1 |
| Concatenations | 1 |
| Filter | Raw filter, Prescan Normalize |
| Coil elements | HC1-7 |

### Contrast - Common

| | |
|---|---|
| TR | 2530.0 ms |
| TE 1 | 1.69 ms |
| TE 2 | 3.55 ms |
| TE 3 | 5.41 ms |
| TE 4 | 7.27 ms |
| Magn. preparation | Non-sel. IR |
| TI | 1100 ms |
| Flip angle | 7.0 deg |
| Fat suppr. | None |
| Water suppr. | None |

### Contrast - Dynamic

| | |
|---|---|
| Averages | 1 |
| Averaging mode | Long term |
| Reconstruction | Magnitude |
| Measurements | 1 |
| Multiple series | Each measurement |

### Resolution - Common

| | |
|---|---|
| FoV read | 256 mm |
| FoV phase | 100.0 % |
| Slice thickness | 1.00 mm |
| Base resolution | 256 |
| Phase resolution | 100 % |
| Slice resolution | 100 % |
| Phase partial Fourier | Off |
| Slice partial Fourier | Off |
| Interpolation | Off |

### Resolution - iPAT

| | |
|---|---|
| PAT mode | GRAPPA |
| Accel. factor PE | 2 |
| Ref. lines PE | 32 |
| Accel. factor 3D | 1 |
| Reference scan mode | Integrated |

### Resolution - Filter Image

| | |
|---|---|
| Image Filter | Off |
| Distortion Corr. | Off |
| Prescan Normalize | On |
| Unfiltered images | Off |
| Normalize | Off |
| B1 filter | Off |

### Resolution - Filter Rawdata

| | |
|---|---|
| Raw filter | On |
| Elliptical filter | Off |

### Geometry - Common

| | |
|---|---|
| Slab group | 1 |
| Slabs | 1 |
| Dist. factor | 50 % |
| Position | R2.2 P1.7 F31.8 mm |
| Orientation | Sagittal |
| Phase enc. dir. | A >> P |
| Slice oversampling | 0.0 % |
| Slices per slab | 176 |
| FoV read | 256 mm |
| FoV phase | 100.0 % |
| Slice thickness | 1.00 mm |
| TR | 2530.0 ms |
| Multi-slice mode | Single shot |
| Series | Interleaved |
| Concatenations | 1 |

### Geometry - AutoAlign

| | |
|---|---|
| Slab group | 1 |
| Position | R2.2 P1.7 F31.8 mm |
| Orientation | Sagittal |
| Phase enc. dir. | A >> P |
| AutoAlign | Head > Brain |
| Initial Position | R2.2 P1.7 F31.8 |
| R | 2.2 mm |
| P | 1.7 mm |
| F | 31.8 mm |
| Initial Rotation | 0.00 deg |
| Initial Orientation | Sagittal |

### Geometry - Navigator

### System - Miscellaneous

| | |
|---|---|
| Positioning mode | REF |
| Table position | H |
| Table position | 0 mm |
| MSMA | S - C - T |
| Sagittal | R >> L |
| Coronal | A >> P |
| Transversal | F >> H |
| Coil Combine Mode | Adaptive Combine |
| Save uncombined | Off |
| Matrix Optimization | Off |
| AutoAlign | Head > Brain |
| Coil Select Mode | Default |

### System - Adjustments

| | |
|---|---|
| B0 Shim mode | Standard |
| B1 Shim mode | TrueForm |
| Adjust with body coil | On |
| Confirm freq. adjustment | Off |
| Assume Dominant Fat | Off |
| Assume Silicone | Off |
| Adjustment Tolerance | Auto |

### System - Adjust Volume

| | |
|---|---|
| Position | R2.2 P1.7 F31.8 mm |
| Orientation | Sagittal |
| Rotation | 0.00 deg |
| A >> P | 256 mm |
| F >> H | 256 mm |
| R >> L | 176 mm |
| Reset | Off |

### System - pTx Volumes

| | |
|---|---|
| B1 Shim mode | TrueForm |
| Excitation | Non-sel. |

### System - Tx/Rx

| | |
|---|---|
| Frequency 1H | 123.261582 MHz |
| Correction factor | 1 |
| Gain | Low |
| Img. Scale Cor. | 1.000 |
| Reset | Off |
| ? Ref. amplitude 1H | 0.000 V |

### Physio - Signal1

| | |
|---|---|
| 1st Signal/Mode | None |
| TR | 2530.0 ms |
| Concatenations | 1 |

### Physio - Cardiac

| | |
|---|---|
| Magn. preparation | Non-sel. IR |
| TI | 1100 ms |
| Fat suppr. | None |
| Dark blood | Off |
| FoV read | 256 mm |
| FoV phase | 100.0 % |
| Phase resolution | 100 % |

### Physio - PACE

| | |
|---|---|
| Resp. control | Off |
| Concatenations | 1 |

### Inline - Common

| | |
|---|---|
| Subtract | Off |

### Inline - Common

| | |
|---|---|
| Measurements | 1 |
| StdDev | Off |
| Save original images | On |

### Inline - MIP

| | |
|---|---|
| MIP-Sag | Off |
| MIP-Cor | Off |
| MIP-Tra | Off |
| MIP-Time | Off |
| Save original images | On |

### Inline - Composing

| | |
|---|---|
| Distortion Corr. | Off |

### Sequence - Part 1

| | |
|---|---|
| Introduction | Off |
| Dimension | 3D |
| Elliptical scanning | Off |
| Reordering | Linear |
| Asymmetric echo | Off |
| Contrasts | 4 |
| Flow comp. 1 | No |
| Multi-slice mode | Single shot |
| Echo spacing | 9.8 ms |
| Bandwidth 1 | 650 Hz/Px |
| Bandwidth 2 | 650 Hz/Px |
| Bandwidth 3 | 650 Hz/Px |
| Bandwidth 4 | 650 Hz/Px |

### Sequence - Part 2

| | |
|---|---|
| RF pulse type | Fast |
| Gradient mode | Fast |
| Excitation | Non-sel. |
| RF spoiling | On |
| Incr. Gradient spoiling | Off |
| Turbo factor | 176 |

### Sequence - Special

| | |
|---|---|
| Readout polarity | Positive |
| Readout trajectory | Bipolar |
| Gradient spoiling | Siemens |
| Gradient moment factor | 1 |
| Averaging | RMS |

### Sequence - Assistant

| | |
|---|---|
| Mode | Off |

**\\Research\Investigators\Baker\Holmes_Yale_McLean_Dec2019_NOEXPIRATION\t2_tse_dark-fluid_tra**

TA: 4:14 PM: REF Voxel size: 0.7×0.7×4.0 mmPAT: Off Rel. SNR: 1.00 : tir

### Properties

| | |
|---|---|
| Prio recon | Off |
| Load images to viewer | On |
| Inline movie | Off |
| Auto store images | On |
| Load images to stamp segments | On |
| Load images to graphic segments | Off |
| Auto open inline display | Off |
| Auto close inline display | Off |
| Start measurement without further preparation | Off |
| Wait for user to start | Off |
| Start measurements | Single measurement |

### Routine

| | |
|---|---|
| Slice group | 1 |
| Slices | 30 |
| Dist. factor | 25 % |
| Position | L0.0 P0.0 H0.0 mm |
| Orientation | Transversal |
| Phase enc. dir. | R >> L |
| AutoAlign | Head > Brain |
| Phase oversampling | 0 % |
| FoV read | 220 mm |
| FoV phase | 96.9 % |
| Slice thickness | 4.0 mm |
| TR | 9000.0 ms |
| TE | 83.0 ms |
| Averages | 1 |
| Concatenations | 2 |
| Filter | Prescan Normalize, Elliptical filter |
| Coil elements | HC1-7 |

### Contrast - Common

| | |
|---|---|
| TR | 9000.0 ms |
| TE | 83.0 ms |
| TD | 0.0 ms |
| MTC | Off |
| Magn. preparation | Slice-sel. IR |
| TI | 2500 ms |
| Flip angle | 150 deg |
| Fat suppr. | Fat sat. |
| Fat sat. mode | Strong |
| Water suppr. | None |
| Restore magn. | Off |
| Freeze suppressed tissue | On |

### Contrast - Dynamic

| | |
|---|---|
| Averages | 1 |
| Averaging mode | Long term |
| Reconstruction | Magnitude |
| Measurements | 1 |
| Multiple series | Each measurement |

### Resolution - Common

| | |
|---|---|
| FoV read | 220 mm |
| FoV phase | 96.9 % |
| Slice thickness | 4.0 mm |
| Base resolution | 320 |

### Resolution - Common

| | |
|---|---|
| Phase resolution | 70 % |
| Phase partial Fourier | Off |
| Trajectory | Cartesian |
| Interpolation | Off |

### Resolution - iPAT

| | |
|---|---|
| PAT mode | None |

### Resolution - Filter Image

| | |
|---|---|
| Image Filter | Off |
| Distortion Corr. | Off |
| Prescan Normalize | On |
| Unfiltered images | Off |
| Normalize | Off |
| B1 filter | Off |

### Resolution - Filter Rawdata

| | |
|---|---|
| Raw filter | Off |
| Elliptical filter | On |

### Geometry - Common

| | |
|---|---|
| Slice group | 1 |
| Slices | 30 |
| Dist. factor | 25 % |
| Position | L0.0 P0.0 H0.0 mm |
| Orientation | Transversal |
| Phase enc. dir. | R >> L |
| FoV read | 220 mm |
| FoV phase | 96.9 % |
| Slice thickness | 4.0 mm |
| TR | 9000.0 ms |
| Multi-slice mode | Interleaved |
| Series | Interleaved |
| Concatenations | 2 |

### Geometry - AutoAlign

| | |
|---|---|
| Slice group | 1 |
| Position | L0.0 P0.0 H0.0 mm |
| Orientation | Transversal |
| Phase enc. dir. | R >> L |
| AutoAlign | Head > Brain |
| Initial Position | L0.0 P0.0 F37.0 |
| L | 0.0 mm |
| P | 0.0 mm |
| H | 37.0 mm |
| Initial Rotation | 90.00 deg |
| Initial Orientation | Transversal |

### Geometry - Saturation

| | |
|---|---|
| Fat suppr. | Fat sat. |
| Fat sat. mode | Strong |
| Water suppr. | None |
| Restore magn. | Off |
| Special sat. | None |

### Geometry - Navigator

### System - Miscellaneous

| | |
|---|---|
| Positioning mode | REF |
| Table position | H |
| Table position | 37 mm |
| MSMA | S - C - T |
| Sagittal | R >> L |
| Coronal | A >> P |
| Transversal | F >> H |
| Coil Combine Mode | Adaptive Combine |
| Save uncombined | Off |
| Matrix Optimization | Off |
| AutoAlign | Head > Brain |
| Coil Select Mode | Off - AutoCoilSelect |

### System - Adjustments

| | |
|---|---|
| B0 Shim mode | Standard |
| B1 Shim mode | TrueForm |
| Adjust with body coil | On |
| Confirm freq. adjustment | Off |
| Assume Dominant Fat | Off |
| Assume Silicone | Off |
| Adjustment Tolerance | Auto |

### System - Adjust Volume

| | |
|---|---|
| Position | L0.0 P0.0 H0.0 mm |
| Orientation | Transversal |
| Rotation | 90.00 deg |
| R >> L | 214 mm |
| A >> P | 220 mm |
| F >> H | 149 mm |
| Reset | Off |

### System - pTx Volumes

| | |
|---|---|
| B1 Shim mode | TrueForm |

### System - Tx/Rx

| | |
|---|---|
| Frequency 1H | 123.261582 MHz |
| Correction factor | 1 |
| Gain | High |
| Img. Scale Cor. | 1.000 |
| Reset | Off |
| ? Ref. amplitude 1H | 0.000 V |

### Physio - Signal1

| | |
|---|---|
| 1st Signal/Mode | None |
| TR | 9000.0 ms |
| Concatenations | 2 |

### Physio - Cardiac

| | |
|---|---|
| Magn. preparation | Slice-sel. IR |
| TI | 2500 ms |
| Fat suppr. | Fat sat. |
| Dark blood | Off |
| FoV read | 220 mm |
| FoV phase | 96.9 % |
| Phase resolution | 70 % |
| Trajectory | Cartesian |

### Physio - PACE

| | |
|---|---|
| Resp. control | Off |
| Concatenations | 2 |

### Inline - Common

| | |
|---|---|
| Subtract | Off |

### Inline - Common

| | |
|---|---|
| Measurements | 1 |
| StdDev | Off |
| Save original images | On |

### Inline - MIP

| | |
|---|---|
| MIP-Sag | Off |
| MIP-Cor | Off |
| MIP-Tra | Off |
| MIP-Time | Off |
| Save original images | On |

### Inline - Composing

| | |
|---|---|
| Distortion Corr. | Off |

### Sequence - Part 1

| | |
|---|---|
| Introduction | On |
| Dimension | 2D |
| Compensate T2 decay | Off |
| Reduce Motion Sens. | On |
| Contrasts | 1 |
| Flow comp. | No |
| Multi-slice mode | Interleaved |
| Free echo spacing | Off |
| Echo spacing | 8.28 ms |
| Bandwidth | 289 Hz/Px |

### Sequence - Part 2

| | |
|---|---|
| Define | Turbo factor |
| Echo trains per slice | 13 |
| Phase correction | Automatic |
| Acoustic noise reduction | None |
| RF pulse type | Normal |
| Gradient mode | Performance |
| Hyperecho | Off |
| WARP | Off |
| Red. EC sensitivity | Off |
| Turbo factor | 17 |

### Sequence - Assistant

| | |
|---|---|
| Mode | Off |
| Allowed delay | 60 s |

| \\Research\Investigators\Baker\Holmes_Yale_McLean_Dec2019_NOEXPIRATION\pd+t2_tse_tra |
|---|
| TA: 2:11 PM: FIX Voxel size: 0.3×0.3×4.0 mmPAT: 2 Rel. SNR: 1.00 : tse |

### Properties

| | |
|---|---|
| Prio recon | Off |
| Load images to viewer | On |
| Inline movie | Off |
| Auto store images | On |
| Load images to stamp segments | Off |
| Load images to graphic segments | Off |
| Auto open inline display | Off |
| Auto close inline display | Off |
| Start measurement without further preparation | Off |
| Wait for user to start | Off |
| Start measurements | Single measurement |

### Routine

| | |
|---|---|
| Slice group | 1 |
| Slices | 30 |
| Dist. factor | 25 % |
| Position | L0.0 P0.0 H0.0 mm |
| Orientation | Transversal |
| Phase enc. dir. | R >> L |
| AutoAlign | Head > Brain |
| Phase oversampling | 0 % |
| FoV read | 220 mm |
| FoV phase | 100.0 % |
| Slice thickness | 4.0 mm |
| TR | 3600.0 ms |
| TE 1 | 9.4 ms |
| TE 2 | 94 ms |
| Averages | 1 |
| Concatenations | 1 |
| Filter | Prescan Normalize, Elliptical filter |
| Coil elements | HC1-7 |

### Contrast - Common

| | |
|---|---|
| TR | 3600.0 ms |
| TE 1 | 9.4 ms |
| TE 2 | 94 ms |
| MTC | Off |
| Magn. preparation | None |
| Flip angle | 160 deg |
| Fat suppr. | None |
| Water suppr. | None |
| Restore magn. | Off |

### Contrast - Dynamic

| | |
|---|---|
| Averages | 1 |
| Averaging mode | Long term |
| Reconstruction | Magnitude |
| Measurements | 1 |
| Multiple series | Each measurement |

### Resolution - Common

| | |
|---|---|
| FoV read | 220 mm |
| FoV phase | 100.0 % |
| Slice thickness | 4.0 mm |
| Base resolution | 320 |
| Phase resolution | 100 % |
| Phase partial Fourier | Off |
| Trajectory | Cartesian |

### Resolution - Common

| | |
|---|---|
| Interpolation | On |

### Resolution - iPAT

| | |
|---|---|
| PAT mode | GRAPPA |
| Accel. factor PE | 2 |
| Ref. lines PE | 30 |
| Reference scan mode | Integrated |

### Resolution - Filter Image

| | |
|---|---|
| Image Filter | Off |
| Distortion Corr. | Off |
| Prescan Normalize | On |
| Unfiltered images | Off |
| Normalize | Off |
| B1 filter | Off |

### Resolution - Filter Rawdata

| | |
|---|---|
| Raw filter | Off |
| Elliptical filter | On |

### Geometry - Common

| | |
|---|---|
| Slice group | 1 |
| Slices | 30 |
| Dist. factor | 25 % |
| Position | L0.0 P0.0 H0.0 mm |
| Orientation | Transversal |
| Phase enc. dir. | R >> L |
| FoV read | 220 mm |
| FoV phase | 100.0 % |
| Slice thickness | 4.0 mm |
| TR | 3600.0 ms |
| Multi-slice mode | Interleaved |
| Series | Interleaved |
| Concatenations | 1 |

### Geometry - AutoAlign

| | |
|---|---|
| Slice group | 1 |
| Position | L0.0 P0.0 H0.0 mm |
| Orientation | Transversal |
| Phase enc. dir. | R >> L |
| AutoAlign | Head > Brain |
| Initial Position | L0.0 P0.0 F37.0 |
| L | 0.0 mm |
| P | 0.0 mm |
| H | 37.0 mm |
| Initial Rotation | 90.00 deg |
| Initial Orientation | Transversal |

### Geometry - Saturation

| | |
|---|---|
| Fat suppr. | None |
| Water suppr. | None |
| Restore magn. | Off |
| Special sat. | None |

### Geometry - Navigator

### System - Miscellaneous

| | |
|---|---|
| Positioning mode | FIX |
| Table position | H |

### System - Miscellaneous

| | |
|---|---|
| Table position | 37 mm |
| MSMA | S - C - T |
| Sagittal | R >> L |
| Coronal | A >> P |
| Transversal | F >> H |
| Coil Combine Mode | Adaptive Combine |
| Save uncombined | Off |
| Matrix Optimization | Off |
| AutoAlign | Head > Brain |
| Coil Select Mode | Off - AutoCoilSelect |

### System - Adjustments

| | |
|---|---|
| B0 Shim mode | Tune up |
| B1 Shim mode | TrueForm |
| Adjust with body coil | On |
| Confirm freq. adjustment | Off |
| Assume Dominant Fat | Off |
| Assume Silicone | Off |
| Adjustment Tolerance | Auto |

### System - Adjust Volume

| | |
|---|---|
| Position | Isocenter |
| Orientation | Transversal |
| Rotation | 0.00 deg |
| A >> P | 263 mm |
| R >> L | 350 mm |
| F >> H | 350 mm |
| Reset | Off |

### System - pTx Volumes

| | |
|---|---|
| B1 Shim mode | TrueForm |

### System - Tx/Rx

| | |
|---|---|
| Frequency 1H | 123.261582 MHz |
| Correction factor | 1 |
| Gain | High |
| Img. Scale Cor. | 1.000 |
| Reset | Off |
| ? Ref. amplitude 1H | 0.000 V |

### Physio - Signal1

| | |
|---|---|
| 1st Signal/Mode | None |
| TR | 3600.0 ms |
| Concatenations | 1 |

### Physio - Cardiac

| | |
|---|---|
| Magn. preparation | None |
| Fat suppr. | None |
| Dark blood | Off |
| FoV read | 220 mm |
| FoV phase | 100.0 % |
| Phase resolution | 100 % |
| Trajectory | Cartesian |

### Physio - PACE

| | |
|---|---|
| Resp. control | Off |
| Concatenations | 1 |

### Inline - Common

| | |
|---|---|
| Subtract | Off |
| Measurements | 1 |
| StdDev | Off |
| Save original images | On |

### Inline - MIP

| | |
|---|---|
| MIP-Sag | Off |
| MIP-Cor | Off |
| MIP-Tra | Off |
| MIP-Time | Off |
| Save original images | On |

### Inline - Composing

| | |
|---|---|
| Distortion Corr. | Off |

### Sequence - Part 1

| | |
|---|---|
| Introduction | On |
| Dimension | 2D |
| Compensate T2 decay | Off |
| Reduce Motion Sens. | On |
| Contrasts | 2 |
| Flow comp. | No |
| Multi-slice mode | Interleaved |
| Free echo spacing | Off |
| Echo spacing | 9.36 ms |
| Bandwidth | 252 Hz/Px |

### Sequence - Part 2

| | |
|---|---|
| Define | Turbo factor |
| Echo trains per slice | 35 |
| Phase correction | Automatic |
| Acoustic noise reduction | None |
| RF pulse type | Normal |
| Gradient mode | Normal |
| Hyperecho | Off |
| WARP | Off |
| Red. EC sensitivity | Off |
| Turbo factor | 5 |

### Sequence - Assistant

| | |
|---|---|
| Mode | Min flip angle |
| Min flip angle | 130 deg |
| Allowed delay | 60 s |

**\\Research\Investigators\Baker\Holmes_Yale_McLean_Dec2019_NOEXPIRATION\resolve_4scan_trace_tra_p2_192**

TA: 1:51 PM: REF Voxel size: 1.1×1.1×4.0 mmPAT: 2 Rel. SNR: 1.00 : resolve

### Properties

| | |
|---|---|
| Prio recon | Off |
| Load images to viewer | On |
| Inline movie | Off |
| Auto store images | On |
| Load images to stamp segments | Off |
| Load images to graphic segments | Off |
| Auto open inline display | Off |
| Auto close inline display | Off |
| Start measurement without further preparation | Off |
| Wait for user to start | Off |
| Start measurements | Single measurement |

### Routine

| | |
|---|---|
| Slice group | 1 |
| Slices | 30 |
| Dist. factor | 25 % |
| Position | L0.0 P0.0 H0.0 mm |
| Orientation | Transversal |
| Phase enc. dir. | A >> P |
| AutoAlign | Head > Brain |
| Phase oversampling | 0 % |
| FoV read | 220 mm |
| FoV phase | 100.0 % |
| Slice thickness | 4.0 mm |
| TR | 4040 ms |
| TE 1 | 55 ms |
| TE 2 | 93 ms |
| Concatenations | 1 |
| Filter | Raw filter, Prescan Normalize |
| Coil elements | HC1-7;NC1,2 |

### Contrast - Common

| | |
|---|---|
| TR | 4040 ms |
| TE 1 | 55 ms |
| TE 2 | 93 ms |
| MTC | Off |
| Magn. preparation | None |
| Flip angle | 180 deg |
| Fat suppr. | Fat sat. |
| Fat sat. mode | Strong |

### Contrast - Dynamic

| | |
|---|---|
| Averaging mode | Short term |
| Reconstruction | Magnitude |
| Measurements | 1 |

### Resolution - Common

| | |
|---|---|
| FoV read | 220 mm |
| FoV phase | 100.0 % |
| Slice thickness | 4.0 mm |
| Base resolution | 192 |
| Phase resolution | 100 % |
| Phase partial Fourier | Off |
| Readout partial Fourier | 5/8 |
| Readout segments | 7 |
| Interpolation | Off |

### Resolution - iPAT

| | |
|---|---|
| PAT mode | GRAPPA |
| Accel. factor PE | 2 |
| Ref. lines PE | 96 |
| Reference scan mode | EPI/separate |

### Resolution - Filter Image

| | |
|---|---|
| Distortion Corr. | Off |
| Prescan Normalize | On |
| Unfiltered images | Off |

### Resolution - Filter Rawdata

| | |
|---|---|
| Raw filter | On |

### Geometry - Common

| | |
|---|---|
| Slice group | 1 |
| Slices | 30 |
| Dist. factor | 25 % |
| Position | L0.0 P0.0 H0.0 mm |
| Orientation | Transversal |
| Phase enc. dir. | A >> P |
| FoV read | 220 mm |
| FoV phase | 100.0 % |
| Slice thickness | 4.0 mm |
| TR | 4040 ms |
| Multi-slice mode | Interleaved |
| Series | Interleaved |
| Concatenations | 1 |

### Geometry - AutoAlign

| | |
|---|---|
| Slice group | 1 |
| Position | L0.0 P0.0 H0.0 mm |
| Orientation | Transversal |
| Phase enc. dir. | A >> P |
| AutoAlign | Head > Brain |
| Initial Position | L0.0 P0.0 F37.0 |
| L | 0.0 mm |
| P | 0.0 mm |
| H | 37.0 mm |
| Initial Rotation | 0.00 deg |
| Initial Orientation | Transversal |

### Geometry - Saturation

| | |
|---|---|
| Fat suppr. | Fat sat. |
| Fat sat. mode | Strong |
| Special sat. | None |

### System - Miscellaneous

| | |
|---|---|
| Positioning mode | REF |
| Table position | H |
| Table position | 37 mm |
| MSMA | S - C - T |
| Sagittal | R >> L |
| Coronal | A >> P |
| Transversal | F >> H |
| Coil Combine Mode | Adaptive Combine |
| Save uncombined | Off |
| Matrix Optimization | Off |
| AutoAlign | Head > Brain |
| Coil Select Mode | Off - AutoCoilSelect |

### System - Adjustments

| | |
|---|---|
| B0 Shim mode | Standard |
| B1 Shim mode | TrueForm |
| Adjust with body coil | On |
| Confirm freq. adjustment | Off |
| Assume Dominant Fat | Off |
| Assume Silicone | Off |
| Adjustment Tolerance | Auto |

### System - Adjust Volume

| | |
|---|---|
| Position | L0.0 P0.0 H0.0 mm |
| Orientation | Transversal |
| Rotation | 0.00 deg |
| A >> P | 220 mm |
| R >> L | 220 mm |
| F >> H | 149 mm |
| Reset | Off |

### System - pTx Volumes

| | |
|---|---|
| B1 Shim mode | TrueForm |

### System - Tx/Rx

| | |
|---|---|
| Frequency 1H | 123.261582 MHz |
| Correction factor | 1 |
| Gain | High |
| Img. Scale Cor. | 1.000 |
| Reset | Off |
| ? Ref. amplitude 1H | 0.000 V |

### Physio - Signal1

| | |
|---|---|
| 1st Signal/Mode | None |
| TR | 4040 ms |
| Concatenations | 1 |

### Diff - Neuro

| | |
|---|---|
| Diffusion mode | 4-Scan Trace |
| Diff. directions | 4 |
| Diffusion Scheme | Monopolar |
| Diff. weightings | 2 |
| b-value 1 | 0 s/mm² |
| b-value 2 | 1000 s/mm² |
| b-value 1 | 1 |
| b-value 2 | 1 |
| Diff. weighted images | Off |
| Trace weighted images | On |
| ADC maps | On |
| FA maps | Off |
| Mosaic | Off |
| Tensor | Off |
| Noise level | 100 |

### Diff - Body

| | |
|---|---|
| Diffusion mode | 4-Scan Trace |
| Diff. directions | 4 |
| Diffusion Scheme | Monopolar |
| Diff. weightings | 2 |
| b-value 1 | 0 s/mm² |
| b-value 2 | 1000 s/mm² |
| b-value 1 | 1 |
| b-value 2 | 1 |
| Diff. weighted images | Off |
| Trace weighted images | On |
| ADC maps | On |
| Exponential ADC Maps | Off |
| FA maps | Off |

### Diff - Body

| | |
|---|---|
| Invert Gray Scale | Off |
| Calculated Image | Off |
| b-Value >= | 0 s/mm² |
| Noise level | 100 |

### Diff - Composing

| | |
|---|---|
| Distortion Corr. | Off |

### Sequence - Part 1

| | |
|---|---|
| Introduction | On |
| Dimension | 2D |
| Contrasts | 2 |
| Optimization | Min. TE |
| Multi-slice mode | Interleaved |
| Echo spacing | 0.32 ms |
| Bandwidth | 766 Hz/Px |

### Sequence - Part 2

| | |
|---|---|
| EPI factor | 96 |
| RF pulse type | Normal |
| Gradient mode | Fast |
| Reacquisition mode | On |

### Sequence - Assistant

| | |
|---|---|
| Mode | Off |
