## Appendix for "The Transdiagnostic Connectome Project: a richly phenotyped open dataset for advancing the study of brain-behavior relationships in psychiatry": McLean_Consent_Form_032322_ CLEAN.pdf

Subject Identification

---

Protocol Title: Neurogenetic analysis in unipolar and bipolar depression

Principal Investigator: Justin Baker, MD, PhD

Site Principal Investigator: Justin Baker, MD, PhD

Description of Subject Population: Adults with unipolar depression, bipolar depression and schizophrenia as well as other anxiety and affective disorders and adults without psychiatric diagnoses

#### About this consent form

Please read this form carefully. It tells you important information about a research study. A member of our research team will also talk to you about taking part in this research study. People who agree to take part in research studies are called "subjects." This term will be used throughout this consent form.

Partners HealthCare System is made up of Partners hospitals, health care providers, and researchers. In the rest of this consent form, we refer to the Partners system simply as "Partners."

If you decide to take part in this research study, you must sign this form to show that you want to take part. We will give you a signed copy of this form to keep.

### Key Information

Taking part in this research study is up to you. You can decide not to take part. If you decide to take part now, you can change your mind and drop out later. Your decision won't change the medical care you get within Partners now or in the future.

The following key information is to help you decide whether or not to take part in this research study. We have included more details about the research in the Detailed Information section that follows the key information.

- We are asking you to join a research study.
- The purpose of the research study is to find out how genes play a role in unipolar and bipolar depression and related mental health conditions, and whether these genes make an

impact on brain structure and function.

- Study activities will include a diagnostic interview, questionnaires, and an MRI scan session. You will complete a diagnostic interview with a member of the research staff. Following the diagnostic interview if you qualify to participate in the study you will complete a battery of demographic questionnaires. Then, we will then ask you to make several different kinds of decisions in a series of separate tests or "tasks" inside the MRI scanner. A sample of blood (about 30ml) may also be drawn.
- Your participation will require up to 4 hours for each study session. The total study may be split over multiple days.
- Risks of participating in this study are minimal. However, some questionnaire items may be considered sensitive and there is a possible risk of loss of confidentiality; every effort will be made to keep your information confidential, but this cannot be guaranteed. MRI scans are *completely non-invasive* and use radio waves in combination with a magnetic field to record images of your body. We want you to carefully read and answer the questions on the MR Safety Questionnaire related to your personal safety.
- There are no direct benefits to participating in this research. The information obtained in this study may provide new therapies for patients diagnosed with affective and related disorders. It is anticipated that your health will in no way be directly affected by this research. Results of the study are purely scientific and will not influence your treatment.
- Taking part in this study is your choice: you can choose to take part, or you can choose not to take part in this study. Whatever choice you make will not harm your relationship with your own doctors, research staff or the McLean Hospital.
- If you are interested in learning more about the study, please continue reading, or have someone read to you, the rest of this document. Ask the study staff questions about anything you do not understand. Once you understand the study, we will ask you if you wish to participate; if so, you will have to sign this form. We will give you a signed copy of this form to keep.

#### Why is this research study being done?

In this research study we want to learn more about how genes a role in unipolar depression, bipolar depression, schizophrenia and related mental health conditions, and whether these genes make an impact on brain structure and function. Genes are made of material called DNA and carry instructions for making the body's proteins. This study is designed to see if and how genes are involved in how the brain works.

#### How long will you take part in this research study?

If you decide to join this research study, it will take you about 4 hours for each study session and may be split into multiple days.

#### What will happen if you take part in this research study?

If you decide to join this research study, the following things will happen:

**Study Procedures**
If you agree to be in this study, the following will happen:

1) You will complete a diagnostic interview with a member of the research staff. This may be the person who initially approached you about this study or it may be a psychiatrist or a clinical psychologist. The interview will take 1-2 hours on average, but in some cases it may take up to four hours divided over two or three days.
2) Following the diagnostic interview if you qualify to participate in the study you will complete a battery of demographic questionnaires. This will take approximately 1-2 hours on average. The study visit may be conducted in person at or virtually using Zoom Video Conferencing.
3) A sample of blood (about 30ml) may be drawn. This procedure takes approximately 2 minutes.
4) You will be escorted by research staff to the McLean Hospital Brain Imaging Center to undergo an MRI scan for up to 60 minutes (more details below). During the MRI session you will complete a battery of demographic questionnaires. This will take approximately an hour on average
5) You may be given the option to download a mobile application for your smartphone. If you agree to do so, the application will collect behavioral data as well as present you with prompts to answer a small number of questions or to provide a voice recording once or twice per day. The behavioral data will be collected continuously over a one-week period, beginning on the day that you agree to participate in the study and sign this consent form. Examples of behavioral data include: accelerometer data, which may be collected to estimate physical activity and sleep patterns, GPS data, which may collected to estimate travel, and text message metadata / logs, which may be collected to estimate frequency of social interaction (text message content and identity of individual texted will never be recorded; no data will ever be recorded without the full consent of the participant, and the data will be encrypted on the phone such that, even if the phone were stolen, no information from our application could be recovered). The survey questions and voice recordings should take you about two minutes to complete and will be presented during the same one-week period. *Note- Some of the surveys involve questions related to*

*sensitive topics that may cause distress. More specifically, question 15 deals with suicidal ideation. All data collected via the application (e.g., survey responses and passive behavioral data) is NOT monitored by research staff in real time and therefore does substitute the application's data collecting features for contact with a medical team. Should a response for this question be higher than a 0, you will be sent an automated inapp message with a button to call the National Suicide Hotline.*

6) You may be given the option to complete supplemental questionnaires. This portion of the study will include various assessments that can be completed online. If you choose to complete this portion of the study, you can either use a computer at the McLean Hospital Brain Imaging Center, directly after your MRI scan, or you can complete these measures after you leave the imaging center within 7 days of your study visit.

You can withdraw from any portion of the study at any time, including during the scan.
If you are enrolled in this study, we will be sharing your data with another McLean study protocol (#2015P002189 "Circuit dynamics underlying longitudinal fluctuations in mood and cognition in bipolar patients"). By signing this form, you give us permission to share your data between this study and the above-mentioned study.

#### Why might you choose to take part in this study?

There are no direct benefits to participating in this research. The information obtained in this study may provide new therapies for patients diagnosed with affective and related disorders. It is anticipated that your health will in no way be directly affected by this research. Results of the study are purely scientific and will not influence your treatment. One potential benefit of participating in this research is that MRI scans obtained for this research will be analyzed by a radiologist and significant findings will be reported to the participants. follow-up for these participants.

#### Why might you choose NOT to take part in this study?

Taking part in this research study has some risks and requirements that you should consider carefully.

Important risks and possible discomforts to know about include:

Blood draw may cause a small arm bruise and, in rare cases, clot or infection at the site the blood was drawn. Some people become light-headed during or immediately after a blood draw. Proper sterile techniques are used to ensure safety for study participants. Some questions during the diagnostic interview may make you uncomfortable. You do not have to answer any questions you do not feel comfortable answering. In addition, there is the potential for a breach of your confidentiality. This could lead your employer, insurance company, or others to find out that you participated in a research study, or to discover genetic information about you. Steps we take to prevent this are described below in the Confidentiality section.

A detailed description of side effects, risks, and possible discomforts can be found later in this consent form in the section called “What are the risks and possible discomforts from being in this research study?”

For virtual encounters, both the participant and study staff must find a private, secure area to conduct the interview. The study staff will use Zoom videoconferencing under the Partners license, on an encrypted device that is compliant with Partners security policies. Recordings of encounters will be saved locally on Partners Dropbox.

TestMyBrain: There are no significant risks associated with the administration of the cognitive tasks, besides mild boredom, fatigue, or frustration. The TestMyBrain system is implemented as a virtual Linux host, so the primary danger from malware is malicious SQL injection attacks. To guard against this, all entered data are sanitized before being included in any SQL updates.

#### If you have questions or concerns about this research study, whom can you call?

You can call us with your questions or concerns. Our telephone numbers are listed below. Ask questions as often as you want. Justin Baker, MD, PhD is the person in charge of this research study. You can call him at 617384-8214 on Mondays-Fridays from 9AM to 5PM. You can also call Bryce Gillis 671 855 2018 with questions about this research study.
If you have questions about the scheduling of appointments or study visits, call Bryce Gillis 671 855 2018.

If you want to speak with someone **not** directly involved in thi44s research study, please contact the Partners Human Research Committee office. You can call them at 857-282-1900.
You can talk to them about:

- Your rights as a research subject
- Your concerns about the research
- A complaint about the research
- Any pressure to take part in, or to continue in the research study

### Detailed Information

A description of this clinical trial will be available on *http://www.ClinicalTrials.gov,* as required by U.S. Law. This Web site will not include information that can identify you. At most, the Web site will include a summary of the results. You can search this Web site at any time.

#### Why is this research study being done?

In this research study we want to learn more about how genes a role in unipolar depression, bipolar depression, schizophrenia and related mental health conditions, and whether these genes make an impact on brain structure and function. Genes are made of material called DNA and carry instructions for making the body's proteins. This study is designed to see if and how genes are involved in how the brain works.

#### Who will take part in this research?

Study population: individuals diagnosed with unipolar depression, bipolar disorder, and schizophrenia as well as healthy comparison participants.

About 525 people will take part in this research study. About 225 subjects will take part at McLean (300 at Yale). Sponsor/funding information: National Institute of Mental Health (NIMH), Yale University.

#### What will happen in this research study?

##### Study Procedures

If you agree to be in this study, the following will happen:

1) You will complete a diagnostic interview with a member of the research staff. This may be the person who initially approached you about this study or it may be a psychiatrist or a clinical psychologist. The interview will take 1-2 hours on average, but in some cases it may take up to four hours divided over two or three days.

2) Following the diagnostic interview if you qualify to participate in the study you will complete a battery of demographic questionnaires. This will take approximately 1-2 hours on average. The study visit may be conducted in person at McLean Hospital, or virtually using Zoom Video Conferencing.

3) A sample of blood (about 30ml) may be drawn. This procedure takes approximately 2 minutes.

4) You will be escorted by research staff to the McLean Hospital Brain Imaging Center to undergo an MRI scan for up to 60 minutes (more details below). During the MRI session you will complete a battery of demographic questionnaires. This will take approximately an hour on average.

5) You may be given the option to download a mobile application for your smartphone. If you agree to do so, the application will collect behavioral data as well as present you with prompts to answer a small number of questions or to provide a voice recording once or twice per day. The behavioral data will be collected continuously over a one-week period, beginning on the day that you agree to participate in the study and sign this consent form. Examples of behavioral data include: accelerometer data, which may be collected to estimate physical activity and sleep patterns, GPS data, which may collected to estimate travel, and text message metadata / logs, which may be collected to estimate frequency of social interaction (text message content and identity of individual texted will never be recorded; no data will ever be recorded without the full consent of the participant, and the data will be encrypted on the phone such that, even if the phone were stolen, no information from our application could be recovered). The survey questions and voice recordings should take you about two minutes to complete and will be presented during the same one-week period. *Note- Some of the surveys involve questions related to sensitive topics that may cause distress. More specifically, question 15 deals with suicidal ideation. All data collected via the application (e.g., survey responses and passive behavioral data) is NOT monitored by research staff in real time and therefore does substitute the application's data collecting features for contact with a medical team. Should a response for this question be higher than a 0, you will be sent an automated inapp message with a button to call the National Suicide Hotline.*

6) You may be given the option to complete supplemental questionnaires. This portion of the study will include various assessments that can be completed online. If you choose to complete this portion of the study, you can either use a computer at the McLean Hospital Brain Imaging Center, directly after your MRI scan, or you can complete these measures after you leave the imaging center within 7 days of your study visit.

You can withdraw from any portion of the study at any time, including during the scan.

#### Use and Storage of Data

Genomic and phenotypic data, and any other data relevant for the study (such as exposure or disease status) will be generated and may be shared broadly and used for future research in a manner consistent with your informed consent and all applicable federal and state laws and regulations.

Prior to submitting the data to an NIH-designated data repository, data will be stripped of identifiers such as name, address, account and other identification numbers and will be deidentified by standards consistent with the Common Rule and HIPAA. Safeguards to protect the data according to Federal standards for information protection will be implemented.

Access to de-identified, individual-level participant data will be controlled, unless participants explicitly consent to allow unrestricted access to and use of their data for any purpose.

Aggregate study information (including genomic summary results) and study analyses may be shared in the scientific literature or through other public scientific resources, such as data repositories or other data sharing resources that provide broad or unrestricted access to the information. The privacy protections, and limitations of those protections, afforded by a Certificate of Confidentiality to individual-level data do not apply to summary results.

Once your blood sample has been obtained, it will be stored in a refrigerator until it can be shipped to our collaborators at Massachusetts General Hospital (MGH). After it is shipped, DNA will be taken from your blood sample and stored. We will keep your DNA sample we have extracted all possible DNA. We will also keep MRI images of your brain stored in a computer.

**How long will my DNA samples and information be kept?**

There is no scheduled date on which your samples and information in the bank will be destroyed. Your samples may be stored for research until they are "used up."

**Can I stop allowing my DNA samples and information to be stored and used for research?**

Yes. You have a right to withdraw your permission, at any time and if you do, your samples and your information will be destroyed. However, it will not be possible to destroy samples and information already given to researchers within Partners or at other academic institutions.
If you decide to take away your permission, contact Dr. Justin Baker at 617-384-8214 or Dr. Avram Holmes at 203-436-9240, Monday-Friday 9-5.

#### How may we use and share your samples and health information for other research?

Your sample and data may also be used in additional research to be conducted by McLean Hospital personnel or researchers outside MacLean Hospital, including our collaborators at Yale University, for the research of unipolar and bipolar depression and related conditions. The information we collect in this study may help advance other research. If you join this study, we

may remove all information that identifies you (for example your name, medical record number, and date of birth) and use these de-identified data in other research.
Information may be shared with investigators at our hospitals, at other academic institutions or at for-profit, commercial entities. You will not be asked to provide additional informed consent for these uses.

As part of your participation in the study, a unique subject number will be assigned to you that will allow researchers to see if you have been involved in more than one research study or database. If you have participated in more than one study or database, this unique subject number will help connect information across studies. This subject number will also allow your de-identified data to be combined with data from other research studies to increase the likelihood of meaningful analysis. Only this subject number and not your personal identifiable information will be accessible to other investigators. This unique subject number may make it possible for a study doctor who used this unique subject number in another study that you took part in to identify you.

Should your sample be shared with researchers outside McLean Hospital, your rights to confidentiality will be strictly observed. Under such circumstances the sample will be coded with a number and will not be labeled with your name or other identifying information. Computer files containing your information will be protected with a password. Your sample and its derivatives may have significant therapeutic or commercial value. You will not receive any financial compensation from your sample or any of its derivatives.

**May we share your MRI, phenotypic, whole genome analysis, and genetic information with the NIH and other central repositories?**

☐ **Yes** ☐ **No** **Subject Initials ___________**

#### Will you get the results of this research study?

No. This research is only a stepping-stone in understanding the genetic effects on brain function. The investigators cannot share genetic or imaging information with you. This information will not be placed in your medical records and will not be useful in directing your medical treatment. The testing done in this study is to look at how genes pay a role in unipolar and bipolar depression and related mental health conditions, and whether these genes make an impact on brain structure and function. It is not intended to be diagnostic, and the investigators are not looking at the same areas a doctor would in a medical scan. The testing cannot substitute for a scan recommended by your doctor. Sometimes researchers will find an irregular result. If this happens, the PI will consult with a radiologist. If the radiologist suggests follow-up, someone will contact you. If this occurs, we suggest that you follow-up with your doctor. At your request, McLean will provide a copy of the research scan results for your doctor, and we will ask you to sign a release form before we send them to your doctor. You will be responsible for the costs of your doctor's care.

##### Description of MRI and Related Procedures

MRI studies will be conducted at the McLean Hospital Brain Imaging Center. If you agree to participate, you will be studied in a scanner with a 3 Tesla field strength. MRI is an imaging technique that uses a strong magnet to take images and measure activity patterns of the brain.

We will ask you to make several different kinds of decisions in a series of separate tests or "tasks." The experimenter will provide you with instructions about each task right before you are asked to perform that task. Although the instructions for performing each task are different, in general, we will have you view words, pictures, or shapes, and ask you to make decisions about those words, pictures, or shapes. You will make these decisions by pressing a button on a handheld box in the scanner.

Towards the end of the scan, you may be asked to watch movie clips inside the scanner while keeping your eyes open. These clips show neutral situations of people or things and there will be no sound.

If you participate in the MRI scan, the duration may be up to 60 minutes and your pulse and respiratory rate information will be collected with MR-safe monitoring devices. You may be asked to participate in the same scanning protocol at a later point in time to ensure data reliability and to help answer another research question.

If you are a control and have completed this portion of the study previously, you may be asked to complete a second, optional scan which will help us answer a related research question, and ensure data reliability, for which you will be provided additional compensation.

The magnetic resonance (MR) scanner looks like a large cylinder with a tube running down the center. You will be asked to lie down on your back on a foam-padded table and place your head into a special holder. The table will slide you inside the "hole" of the scanner. Soft foam rubber sponges may be placed on both sides of your head for comfort and to help keep your head from moving. Because the scanner contains a strong magnet, you will be asked to remove all metal objects from your person including, but not limited to: watches, rings, necklaces, bracelets, earrings and other body piercings, belts, loose change, wallet (with credit cards), items of clothing containing magnetic materials (for example, underwire bras, certain types of zippers), and shoes.

These items will be secured in a safe place until your scan is completed. You will be able to remain in your street clothes.

This scan session should last no more than 60 minutes. During the scanning procedure itself you will hear a number of different sounds, which are part of the normal operation of the scanner. You are free to talk during the preparation time and during breaks, but you should not talk during the actual scanning process. During scans, you should try to remain as still as possible. When your session is over the technician will move you out of the scanner and assist you from the table. The technologist will be able to see and hear you at all times and you are free to end the procedure at any time.

**Description of Mobile Application Procedures**

Your activity levels, daily mood, social interactions, and sleep habits may be continuously monitored using a smartphone. You will use your existing smartphone by loading software onto it. We will show you how to load the software during the first orientation session. The software must be turned on at all times to participate in this study. The content of your e-mails and text messages, and the identity of the people you are communicating with will never be recorded. Information about your social activities will be recorded such as when you text, how long the message was, and whether it was the same person as on another occasion (even though the phone number and identify of that person will not be captured). The goal of these recordings is to measure your social activity levels, but never to capture the contents of your discussions.

Once or twice a day we may send a message to your smartphone that prompts you to answer a small number of questions as soon as possible. When you receive a prompt, you may be asked to answer questions about how you are feeling, what you are doing, and recent activities including whether you slept well and whether you had alcohol, or tobacco. These questions should take you about two minutes to complete and never more than five minutes. Once a day you will also be asked to make a voice recording where you will talk about how you presently feel and any feelings that you have experienced over the past day in relation to routine or significant events. You are free to provide as much or as little information, so long as the information avoids names and description that could identify individual people. The goal is to allow you to talk about yourself but avoid identifying information about others.

As the app only uploads data to the server when connected to a Wi-Fi network, it is not using the smartphone data plan to upload data. The data use involved in completing surveys on the application will be extremely minimal, and overall not add to the burden of data cost for participants using the application.

**Please check this box if you DO NOT wish to take part in this portion of the study:** ☐

#### What are the risks and possible discomforts from being in this research study?

**Blood draw and Interview:**

Blood draw may cause a small arm bruise and, in rare cases, clot or infection at the site the blood was drawn. Some people become light-headed during or immediately after a blood draw. Proper sterile techniques are used to ensure safety for study participants. Some questions during the diagnostic interview may make you uncomfortable. You do not have to answer any questions you do not feel comfortable answering. The interview and some of the questionnaires may inquire about psychological symptoms such as depression, anxiety, and suicidality. Should you report having suicidal thoughts, steps will be taken to ensure that you are safe, and a list of available mental health resources and/or clinical referrals will be provided to you. In addition, there is the potential for a breach of your confidentiality. This could lead your employer, insurance company, or others to find out that you participated in a research study, or to discover genetic information about you. Steps we take to prevent this are described below in the Confidentiality section. The interview portion of this session may be recorded on a digital, video and/or audio recorder. The purpose of this recording is to check the quality of the work the interviewers are doing. You will not be asked for any identifying information during the taped interview, and no identifying information will be associated with the recording. Associated data files will be stored on a secure sever that only qualified study staff will access. After recordings are checked for interviewer quality, they will be destroyed.

**MRI:**

Unlike X-rays or CAT scans, magnetic resonance (MR) technology does not use ionizing radiation. Instead, it uses strong magnetic fields and radio waves to collect the images and data. There are no known hazards or risks associated with MR techniques. Significant risks may exist for people with:

-Cardiac pacemakers
-Metal clips on blood vessels (also called stents)
-Artificial heart valves
-Artificial arms, hands, legs, etc.
-Brain stimulator devices
-Implanted drug pumps
-Ear implants
-Eye implants or known metal fragments in eyes
-Exposure to shrapnel or metal filings (wounded in military combat, sheetmetal workers, welders, and others)
-Other metallic surgical hardware in vital areas

-Certain tattoos with metallic ink (please tell us if you have a tattoo)
-Certain transdermal (skin) patches such as NicoDerm (nicotine for tobacco dependence), Transderm Scop (scopolamine for motion sickness), or Ortho Evra (birth control)
-Certain IUDs (intrauterine devices) made of certain kinds of metal

If you are unsure whether you have any of these items in your body, you should know that most would have been implanted as part of a surgical procedure. So, trying to remember any past operations may help you remember. You will be asked whether you have any implanted devices or history of exposure to shrapnel or metal filings, and if so, you will not be able to participate in this study.

Significant risks also can arise if certain materials (many types of metal objects) are brought into the scanning area, as they can be pulled into the magnet at great speed. Such items can cause serious injury if they hit you. Therefore, these types of items are not permitted in the scanning area. You will not be allowed to bring anything with you into the scanning room.

The MR exams are painless, and except for pulsating sounds, you will not be aware that scanning is taking place.

The fMRI scan takes place in a 3T scanner. These scanners are approved by the FDA for routine clinical studies in children or adults. Although there are no known risks from these scans, there could be adverse effects that are delayed or very mild, such that they have not yet been recognized. Most people experience no ill effects from these scans, but some people do report claustrophobia (fear of being in enclosed small spaces), dizziness, mild nausea, headaches, a metallic taste in their mouth, double vision, or sensation of flashing lights. These symptoms, if present, disappear shortly after leaving the scanner.

In rare cases, a very slight, uncomfortable tingling of the back due is induced in some people undergoing certain types of scans. If you experience this sensation, you are asked to report this immediately so the scan can be stopped. Although these precautions will avoid all known risks associated with MRI, this procedure may involve risks to you that are currently unforeseeable.
The sounds that you hear inside the scanner are the normal operating sounds the scanner makes while it takes pictures of your brain. The noises vary with the type of scan being performed and include: sounds like a hammer hitting a piece of wood, repetitive buzzing noises, and long series of loud beeps. These sounds, or combinations of them, may be repeated several times. While they may be annoying, their intensity is not harmful to your hearing. However, you will be given a pair of earplugs to wear to muffle the sounds. You also may be asked to wear a set of headphones, which further reduces the noise level and permits the technician to speak to you.

*Women of childbearing age:*

While there are also no known risks for fetuses, the safety of MRI for pregnant women, women of childbearing potential, and nursing mothers has not been established. If you are a woman of childbearing potential, you must be using an IUD, oral contraceptive, or barrier methods or must be abstinent prior to each MRI scan. In addition, you must have a negative pregnancy test prior to the MRI scan. Nursing mothers may not participate in the MRI portion of the study.

#### What are the possible benefits from being in this research study?

There are no direct benefits to participating in this research. The information obtained in this study may provide new therapies for patients diagnosed with affective and related disorders. It is anticipated that your health will in no way be directly affected by this research. Results of the study are purely scientific and will not influence your treatment. One potential benefit of participating in this research is that MRI scans obtained for this research will be analyzed by a radiologist and significant findings will be reported to the participants. We will provide follow-up for these participants

#### What other treatments or procedures are available for your condition?

Not applicable.

#### Can you still get medical care within Partners if you don't take part in this research study, or if you stop taking part?

Yes. Your decision won't change the medical care you get within Partners now or in the future. There will be no penalty, and you won't lose any benefits you receive now or have a right to receive.

We will tell you if we learn new information that could make you change your mind about taking part in this research study.

#### What should you do if you want to stop taking part in the study?

If you take part in this research study, and want to drop out, you should tell us. We will make sure that you stop the study safely. We will also talk to you about follow-up care, if needed.

Also, it is possible that we will have to ask you to drop out of the study before you finish it. If this happens, we will tell you why. We will also help arrange other care for you, if needed.

#### Will you be paid to take part in this research study?

There is no cost for you to participate in this procedure. As an identifier for internal auditing purposes, your social security number is needed because you are receiving payment for participation in this study. McLean Hospital is required to inform the IRS of any payments to you as a participant in research studies in a given calendar year totaling $600 or more. If that occurs, you will receive a 1099 form at the end of the year. No information identifying why you received payment is communicated to either the Hospital's accounting department or the government. This information is kept strictly confidential.

You will be paid for your participation. Participants will be compensated up to $130 for completing the entire study. This includes $25 supplement for completing the optional online battery of cognitive measures. In addition, they will be compensated $10/hr if session 1 runs beyond 3 hours and if fMRI session runs beyond 2.5 hours. Participants who elect to withdraw or fail to meet study criteria will be paid $30/hour for their participation.

If you travel to McLean as an outpatient or as a control to participate in research and are somehow unable to complete any part of the study, you will be compensated up to $25 for the time and travel expenses incurred during your trip here. If you complete a portion of the study, you will be compensated for that portion appropriately.

We may use your samples and information to develop a new product or medical test to be sold. The Sponsor, hospital, and researchers may benefit if this happens. There are no plans to pay you if your samples or information are used for this purpose.

#### What happens if you are injured as a result of taking part in this research study?

We will offer you the care needed to treat any injury that directly results from taking part in this research study. We reserve the right to bill your insurance company or other third parties, if appropriate, for the care you get for the injury. We will try to have these costs paid for, but you may be responsible for some of them. For example, if the care is billed to your insurer, you will be responsible for payment of any deductibles and co-payments required by your insurer.

Injuries sometimes happen in research even when no one is at fault. There are no plans to pay you or give you other compensation for an injury, should one occur. However, you are not giving up any of your legal rights by signing this form.

If you think you have been injured or have experienced a medical problem as a result of taking part in this research study, tell the person in charge of this study as soon as possible. The researcher's name and phone number are listed in the next section of this consent form.

Injuries sometimes happen in research even when no one is at fault. There are no plans to pay you or give you other compensation for an injury, should one occur. However, you are not giving up any of your legal rights by signing this form.

If you think you have been injured or have experienced a medical problem as a result of taking part in this research study, tell the person in charge of this study as soon as possible. The researcher's name and phone number are listed in the beginning of this consent form.

#### If you take part in this research study, how will we protect your privacy?

Federal law requires Partners to protect the privacy of health information and related information that identifies you. We refer to this information as "identifiable information."

**In this study, we may collect identifiable information about you from:**

- Past, present, and future medical records
- Research procedures, including research office visits, tests, interviews, and questionnaires

**Who may see, use, and share your identifiable information and why:**

- Partners researchers and staff involved in this study
- The sponsor(s) of the study, and people or groups it hires to help perform this research or to audit the research
- Other researchers and medical centers that are part of this study
- The Partners ethics board or an ethics board outside Partners that oversees the research
- A group that oversees the data (study information) and safety of this study
- Non-research staff within Partners who need identifiable information to do their jobs, such as for treatment, payment (billing), or hospital operations (such as assessing the quality of care or research)
- People or groups that we hire to do certain work for us, such as data storage companies, accreditors, insurers, and lawyers
- Federal agencies (such as the U.S. Department of Health and Human Services (DHHS) and agencies within DHHS like the Food and Drug Administration, the National Institutes of Health, and the Office for Human Research Protections), state agencies, and

foreign government bodies that oversee, evaluate, and audit research, which may include inspection of your records

- Public health and safety authorities, if we learn information that could mean harm to you or others (such as to make required reports about communicable diseases or about child or elder abuse)
- Other researchers within or outside Partners, for use in other research as allowed by law.

#### Certificate of Confidentiality

A federal Certificate of Confidentiality (Certificate) has been issued for this research to add special protection for information and specimens that may identify you. With a Certificate, unless you give permission (such as in this form) and except as described above, the researchers are not allowed to share your identifiable information or identifiable specimens, including for a court order or subpoena.

Certain information from the research will be put into your medical record and will not be covered by the Certificate. This includes records of medical tests or procedures done at the hospitals and clinics, and information that treating health care providers may need to care for you. Please ask your study doctor if you have any questions about what information will be included in your medical record. Other researchers receiving your identifiable information or specimens are expected to comply with the privacy protections of the Certificate. The Certificate does not stop you from voluntarily releasing information about yourself or your participation in this study.

Even with these measures to protect your privacy, once your identifiable information is shared outside Partners, we cannot control all the ways that others use or share it and cannot promise that it will remain completely private.

Because research is an ongoing process, we cannot give you an exact date when we will either destroy or stop using or sharing your identifiable information. Your permission to use and share your identifiable information does not expire.

The results of this research may be published in a medical book or journal, or used to teach others. However, your name or other identifiable information **will not** be used for these purposes without your specific permission.

#### Your Privacy Rights

You have the right **not** to sign this form that allows us to use and share your identifiable information for research; however, if you don't sign it, you can't take part in this research study.

You have the right to withdraw your permission for us to use or share your identifiable information for this research study. If you want to withdraw your permission, you must notify the person in charge of this research study in writing. Once permission is withdrawn, you cannot continue to take part in the study.

If you withdraw your permission, we will not be able to take back information that has already been used or shared with others, and such information may continue to be used for certain purposes, such as to comply with the law or maintain the reliability of the study. You have the right to see and get a copy of your identifiable information that is used or shared for treatment or for payment. To ask for this information, please contact the person in charge of this research study. You may only get such information after the research is finished.

### Informed Consent and Authorization

##### Statement of Person Giving Informed Consent and Authorization

- I have read this consent form.
- This research study has been explained to me, including risks and possible benefits (if any), other possible treatments or procedures, and other important things about the study.
- I have had the opportunity to ask questions.
- I understand the information given to me.

#### Signature of Subject:

I give my consent to take part in this research study and agree to allow my identifiable information to be used and shared as described above.

_______________________________________ _______________ _______________

Subject Date Time (optional)

---

#### Signature of Study Doctor or Person Obtaining Consent:

**Statement of Study Doctor or Person Obtaining Consent**

- I have explained the research to the study subject.
- I have answered all questions about this research study to the best of my ability.

____________________________________ ____________ ____________

Study Doctor or Person Obtaining Consent    Date    Time (optional)

In order to obtain the best information available, we will contact a first (parent, sibling, or child) or second-degree (aunts, uncles, nieces, nephews, and grandparents) family member, your outpatient doctor, or outpatient case worker. Additionally, in order to facilitate any follow-up scheduling, we may contact them to help in locating you if we cannot reach you through the contact information you provided. We may want to call only one of these people, or all of them to collect reliable information. If we do call, we can only ask questions related to your past history and symptoms for the purposes of our research. We cannot talk to them about anything else, and we cannot provide them information about you. If you want to allow us to contact these people, please check "Yes", if not, check "No". Checking "Yes" gives us permission to call one or all of them, but they can still refuse to talk with us. You may also withdraw permission for us to call them at any time.

☐ Yes: Contact Information________________________________________

☐ No

________________________________________________________

Participant Signature    Date

We may wish to contact you up to five (5) years in the future to see if you are interested in participating in another research study. If you want to allow us to contact you please check "Yes", if not, check "No". Checking "Yes" gives us permission to call you in the future, but you can still refuse to be in future studies. You may also withdraw permission to be contacted at any time by contacting us. By checking this box, you are also agreeing to have information and/or brochures for other research studies in the area for which you are eligible sent to your house. You are free to decline participating in these studies.

☐ Yes

☐ No

Participant Signature Date

Consent Form Version Date: 23 March, 2022
