## Appendix for "The Transdiagnostic Connectome Project: a richly phenotyped open dataset for advancing the study of brain-behavior relationships in psychiatry": Yale_Consent_Form_032322_clean.pdf

#### How long will you take part in this research study?

If you decide to join this research study, it will take you up to 4 hours for each study session and may be split into multiple days. The study visit may be conducted in person at or virtually using Zoom Video Conferencing.

#### What will happen if you take part in this research study?

If you decide to join this research study, the following things will happen:

**Study Procedures**

If you agree to be in this study, the following will happen:

1) You will complete a diagnostic interview with a member of the research staff. This may be the person who initially approached you about this study or it may be a psychiatrist or a clinical psychologist. The interview will take 1-2 hours on average, but in some cases it may take up to four hours divided over two or three days. The study visit may be conducted in person at or virtually using Zoom Video Conferencing.

2) Following the diagnostic interview if you qualify to participate in the study you will complete a battery of demographic questionnaires. This will take approximately 1-2 hours on average

3) A sample of blood (about 30ml) may be drawn. This procedure takes approximately 2 minutes.

4) You will be escorted by research staff to the Yale FAS Brain Imaging Center to undergo an MRI scan for up to 60 minutes (more details below). During the MRI session you will complete a battery of demographic questionnaires. This battery of questionnaires may be conducted in person at or virtually using a HIPAA complaint Zoom conferencing account and will take approximately an hour on average.

#### If you have questions or concerns about this research study, whom can you call?

You can call us with your questions or concerns. Our telephone numbers are listed below. Ask questions as often as you want.

Avram Holmes, PhD, is the person on site in charge of this research study. You can call him at 203-436-9240 on Mondays-Fridays from 9AM to 5AM.

If you have questions about the scheduling of appointments or study visits, call Connor Lawhead or Jocelyn Ricard at 203-436-9449.

If you want to speak with someone **not** directly involved in this research study, please contact the Partners Human Research Committee office. You can call them at 857-282-1900.

The testing done in this study is to look at how genes pay a role in unipolar and bipolar depression and related mental health conditions, and whether these genes make an impact on brain structure and function. It is not intended to be diagnostic, and the investigators are not looking at the same areas a doctor would in a medical scan. The testing cannot substitute for a scan recommended by your doctor. Sometimes researchers will find an irregular result. If this happens, the PI will consult with a radiologist. If a potential structure anomaly is detected, a radiologist or another physician may be asked to review the relevant images according to standard Yale Brain Imaging Center policies. Based on their recommendation (if any), the principal investigator or consulting physician will contact the participant, inform them of the finding, and recommend that they seek medical advice as a precautionary measure. The decision for additional examination or treatment would lie only with the participant and their physician. The investigators, the consulting physician, the Yale Brain Imaging Center, and Yale University are not responsible for any examination or treatment that the participant receives based on these findings. The images collected in this study are not a health care MR exam and for that reason, they will not be made available for health care purposes. You will be responsible for the costs of your doctor's care.

### What happens if you are injured as a result of taking part in this research study?

We will offer you the care needed to treat any injury that directly results from taking part in this research study. You or your insurance carrier will be expected to pay the costs of this treatment. You may be responsible for some of them. For example, if the care is billed to your insurer, you will be responsible for payment of any deductibles and co-payments required by your insurer.

#### Signature of Subject:

I give my consent to take part in this research study and agree to allow my identifiable information to be used and shared as described above.

| ______________________________ | ____________ | ____________ |
|---|---|---|
| Subject | Date | Time (optional) |

Subject Identification
